# A Sham Controlled Randomised Trial evaluating the Safety, Acceptability and Efficacy of Autonomic neuromodulation using Trans-Cutaneous vagal sensory stimulation in uncontrolled Hypertensive patients: Rationale and study design of the SCRATCH-HTN Study

**DOI:** 10.64898/2025.12.12.25342144

**Authors:** Ajay Gupta, David Collier, James Steckelmacher, Jane Field, Olivier Zongo, Mital Patel, George Collett, Everard Mascarenhas, Andrey Gourine, Annastazia Learoyd, Alexander V Gourine, Peter Sever, SCRATCH-HTN Trial Investigators

## Abstract

**Background:** Despite the broad availability of antihypertensive drugs, approximately 40% of hypertensive patients fail to achieve the recommended blood pressure (BP) levels and may require alternative treatment(s). Currently, renal denervation is the only proven non-pharmacological device-based alternative treatment available but is a costly hospital-based invasive procedure that is unlikely to be widely available. Transcutaneous autonomic neuromodulation (tAN), if safe, acceptable and efficacious, can offer a non-invasive, inexpensive, self-administered device-based innovative adjunct or alternative to pharmacological therapy.

**Methods:** SCRATCH-HTN is a double-blind, sham-controlled trial, with 63 participants randomised on a 2:1 basis to receive either tAN or sham-tAN treatment. Hypertensive patients on medications were included if they had elevated systo-diastolic BPs on daytime ambulatory BP monitoring (ABPM) (systolic BP (SBP) of ≥135 and <170 mmHg and mean daytime diastolic BP (DBP) of ≥85 and < 115 mmHg). Participants were trained to self-administer tAN therapy for 30 minutes every day for first 14 days, and then once a week for 10 weeks. The primary endpoint was the change in daytime ambulatory SBP from baseline to 3 months. Secondary endpoints included the change in 24-hour ambulatory and office SBP and DBP, BP variability, heart rate variability, quality of life, and sleep quality from baseline to end-of-treatment. Other exploratory outcomes included evaluation of impact on functional exercise (6-minute walk test), structural and functional changes in the heart, cognitive function and central blood pressures. In a subgroup of patients detailed autonomic functional assessment was conducted at the start and end of the study.

**Conclusion:** The SCRATCH-HTN trial is a phase 2a study testing the safety, acceptability, and potential efficacy of tAN treatment for improving blood pressure control in patients with elevated BP despite medication. It also explores tAN’s effects on sleep, exercise tolerance, heart rate variability, central BP, cardiac structure, and autonomic function. If effective, it could offer a transformative approach to hypertension management.

**Registration:** clinicaltrials.gov (NCT05179343).

## Introduction

Hypertension or high blood pressure (BP) affects more than 1.3 billion people worldwide ^1^. It is the leading risk factor for premature death and disability, and a key contributor to global disease burden ^2–4^. This is driven by its role as the leading global cause of cardiovascular and cerebrovascular disease mortality and morbidity and its significant contribution to the global burden of chronic kidney disease ^5–7^. In England, 1 in 3 adults have hypertension and this rises to over 60% of adults aged ≥ 65 years ^8,9^. As such, the cost to healthcare systems is significant: hypertension is estimated to cost the UK National Health Service (NHS) more than £2.1bn annually^10^.

Despite the widespread availability of antihypertensive drugs, 40% of all patients with hypertension fail to achieve the National Institute for Health and Care Excellence (NICE) recommended BP levels (<140 mmHg systolic and <90 mmHg diastolic) ^9,10^. These patients are classified as having uncontrolled hypertension. Non-adherence to prescribed medications is a major challenge, partly due to drug intolerance/adverse side-effects. Global prevalence of antihypertensive medication non-adherence in diagnosed hypertensive patients is estimated to be around 27-40%^11^ and is present in approximately 84% of those with uncontrolled hypertension^12^ and 37% of those with drug-resistant hypertension^11,13^. As such, there is an urgent, unmet clinical need for an effective therapy for uncontrolled hypertensive patients, including drug-resistant patients.

One possible therapy is device-based autonomic neuromodulation. During the development of hypertension, parasympathetic (vagal) activity declines, concomitantly with sympathetic activation, which can, if left untreated, lead to chronically altered homeostatic autonomic balance. Redressing autonomic imbalance via device-based solutions has already been achieved using catheter-based renal denervation ^14,15^ which appears to be a safe treatment for reducing arterial blood pressure ^14,16^. A recent meta-analysis found modest but significant reductions in 24hr ambulatory systolic BP (SBP, mean reduction = −2.23 mmHg) and 24h ambulatory diastolic BP (DBP, mean reduction = −1.16 mmHg) following renal denervation amongst uncontrolled hypertensives on antihypertensive medication^17^. However, renal denervation is an invasive procedure, requires hospitalisation and tertiary care delivered by experts ^15^. This limits universal adoption, while the duration of the blood pressure lowering effect and the requirement for repeat procedures remains uncertain.

Autonomic balance can potentially be re-instated non-invasively using a method called transcutaneous autonomic neuromodulation (tAN), otherwise known as transcutaneous vagus nerve stimulation (tVNS) ^18^. In this method, electrical stimulation is applied to the regions of the outer ear that are innervated by sensory (afferent) fibres of several cranial and spinal nerves, including the auricular branch of the vagus nerve ^18,19^. These afferent nerve fibres, that constitute 80% of the vagus nerve, transmit sensory information, predominantly from visceral organs, to the nucleus tractus solitarius (NTS) where 95% of vagal afferent fibres terminate. Efferent vagal nerve fibres leaving the NTS via the dorsal vagal nucleus and nucleus ambiguous have various visceral organ targets and influence an array of homeostatic physiological functions including BP. The firing of efferent vagal nerve fibres reduces BP through a range of targets and mechanisms including via cardiovagal innervation of the heart which produces negative inotropic and chronotropic effects, and attenuation of sympathetic activity (via inhibition of the rostroventrolateral medulla) that reduces vasoconstriction and activity of the renin-angiotensin-aldosterone system ^20–22^. Recruitment of these afferent projections through transcutaneous electrical stimulation of the auricular nerve modulates the autonomic control circuits in the brainstem and acutely shifts the autonomic balance towards a net vagal dominance^23–26^, as assessed by baroreflex sensitivity ^27^ and heart rate variability measures ^19,23,28^. These effects are consistent with vagal recruitment and sympathetic inhibition as demonstrated by beneficial effects of tAN in patients with epilepsy^29,30^, coronary artery disease ^31^ and atrial fibrillation^32,33^.

There is recent evidence that autonomic neuromodulation via unilateral tAN applied daily for 3 months reduced BP in untreated young individuals with grade-1 hypertension ^34^. The study was open-labelled, unblinded and BP was measured by patients at home with significant self-reported reductions reported after one month. These data, however, are at odds with the results of the study by Stavrakis et al. 2022^35^ who used the identical device as the above study and treatment protocol involving unilateral daily tAN and observed no change in office BP in patients with heart failure with preserved ejection fraction; 96% of these patients were hypertensive. We noted that most published studies, including these two studies, involving longer periods of tAN used unilateral auricular stimulations; were open-labelled and with inconsistent methods of BP monitoring potentially leading to ascertainment bias.

The SCRATCH-HTN study (s <u>S</u>ham <u>C</u>ontrolled <u>R</u>andomized Control Trial Evaluating the Safety, Acceptability and Efficacy of <u>A</u>utonomic Neuromodulation using <u>T</u>rans<u>c</u>utaneous Vagal Sensory Stimulation in Uncontrolled <u>H</u>ypertensive Patients) tests the hypothesis that tAN treatment is safe and acceptable to the patient, improves the control of BP in hypertension and improves well-being amongst those receiving the active treatment compared to those on sham treatment. In this study, tAN is applied to both ears (bilaterally) using a modified and improved protocol. This stimulation protocol was developed based on significant evidence indicating that applying sensory stimuli bilaterally is more effective than unilateral stimulations in inducing brain plasticity and neuromodulation^36,37^. Using this protocol in our unpublished proof-of-concept study of 10 patients with drug-resistant hypertension, bilateral tAN was found to reduce 24-h SBP by an average of 14 mmHg (Supplementary Figure 1). This set the premise for the current trial protocol, although the findings need critical appraisal given the observational, single-arm nature of the proof-of-concept study conducted in a small number of self-selected individuals.

The data obtained will be used to develop a larger efficacy and cost-effectiveness follow-up study. The SCRATCH-HTN trial also includes a sub-study of participants to evaluate autonomic function at baseline and at the end of the treatment period. This will provide the mechanistic insights on the effects of treatment on components of the autonomic nervous system responses. The present manuscript outlines the protocol for this trial.

## Methods and Analysis

### Patient and Public Involvement

At the time of study development and prior to the start of the trial, a group of hypertensive patients were invited to give their opinion. There were also patient and public involvement representatives in our trial steering committee who helped develop the study design, device logbooks, and questionnaires, and oversaw the conduct of the study.

### Trial design

The SCRATCH-HTN trial is a double-blind, sham-controlled study, with a block randomised on a 2:1 basis where participants are receiving either tAN or sham-tAN treatment. tAN is applied using an electronic device AffeX-CT developed by Afferent Medical Solutions, Ltd (UK).

### Study population

Study participants were recruited from 6 participants-identifying centres (PIC) in and around London (Supplementary Table 1), and all participants were processed for consistency, in one single site at the William Harvey Clinical Research Centre at Queen Mary University of London. Hypertensive patients, who were receiving between one and four antihypertensive medications, were eligible for recruitment if they had elevated systo-diastolic BPs on daytime ambulatory BP monitoring (ABPM): daytime average SBP of ≥135 and <170 mmHg, and daytime average DBP of ≥85 and < 115 mmHg), and presence of one or more of the following conditions: obesity, type 2 diabetes, heart rate ≥ 70 beats per minute, metabolic syndrome, dyslipidaemia or polycystic ovarian syndrome (see Table 1 for summary of key inclusion criteria and further details in Supplementary Table 2). Exclusion criteria included presence of anyone with the following: atrial fibrillation, eGFR < 45 mL/min, type 1 diabetes mellitus, poorly controlled type 2 diabetes (HbA1c > 69 mmol/mol) and/or or receiving insulin therapy, orthostatic hypotension (defined as a fall >20 mmHg in SBP on moving from sitting to standing). The full list of exclusion criteria can be found in Supplementary Table 3.

**Table 1.** Summary of participant inclusion criteria. Full inclusion and exclusion criteria detailed in supplementary tables 1 and 2.

| Summary of participant inclusion criteria |
| --- |
| • Aged $\geq 18$ years and $<80$ years<br>• Taking between 1 to 4 antihypertensive medications (inclusive)<br>• Confirmed diagnosis of hypertension<br>• 24-hour ABPM mean daytime<sup>1</sup> SBP $\geq 135$ mmHg and $<170$ mmHg and mean daytime DBP $\geq 85$ mmHg and $<115$ mmHg |
| Additionally, participant must have **one or more** of the following associated conditions |
| • Obesity (Body Mass Index (BMI) $>30$ kg/m<sup>2</sup> or waist circumference $>94$ cm (men) or $>80$ cm (women) (NB. For participants of South-East Asian/Chinese/Japanese origin these cut-offs are $>90$ cm (men) or $>80$ cm (women))<br>• Type 2 diabetes – controlled or sub-optimally controlled (HbA1c $\leq 8.5\%$ or $\leq 69$ mmol/mol) on diet and/or medications except insulin<br>• Heart rate (any one of the three recordings) $\geq 70$ bpm (measurement taken after 5 minutes of rest in a seated position) or heart rate $\geq 60$ bpm if taking beta-blocker medication<br>• HbA1c $\geq 42$ mmol/mol or fasting glucose (if available) $\geq 5.6$ mmol/L **AND** either low HDL cholesterol ($\leq 1.03$ mmol/L for men and $\leq 1.29$ mmol/L for women) or high triglycerides ($\geq 1.7$ mmol/L)<br>• Both low HDL cholesterol ($\leq 1.03$ mmol/L for men and $\leq 1.29$ mmol/L for women) **AND** high triglycerides ($\geq 1.7$ mmol/L)<br>• Diagnosed or known polycystic ovarian syndrome |

<sup>1</sup>Daytime defined as hours from 0700-2300

### Randomisation

Eligible participants were randomised in a 2:1 ratio to tAN and sham-tAN intervention arms, respectively, using the online randomisation tool ‘<u>Sealed Envelope’</u>. This tool employed block randomisation and a dynamic minimisation approach to ensure balanced allocation between treatment groups^38^. The minimisation algorithm incorporated the following baseline factors: age (<65 years or ≥65 years), sex, body mass index (BMI; <30 or ≥30 kg/m²), and mean daytime average SBP (<160 mmHg or ≥160 mmHg).

### Description of intervention

All randomised participants were provided with identical devices, with allocation to either tAN or sham-tAN arm concealed. On the day of randomisation, all participants received standardised training using a dedicated training device before being assigned their personal study device. Each participant was provided with an individually set stimulation threshold, as well as access to further training resources via user guide (see Supplementary Material 2 – User Guide), demonstration video, and telephone support.

During device training, participants were instructed to place the electrode clips (with electrode surfaces made of electrically-conductive rubber) on the left and right tragi (Figure 1). The current amplitude was then gradually increased by the trainer, starting from 0.1 mA, until the participant felt a tingling sensation, after which it was reduced to set the level of stimulation at ∼1.5 mA below this threshold (Figure 1). Once the threshold current was determined, participants were issued their individual units, which were identical to the training device but with concealed controls. Stimulation current was set to 0 mA (sham-tAN) or ∼1.5 mA below the individual perception threshold (tAN), with 200 μs pulses generated at a frequency of 30 Hz.

**Figure 1.**
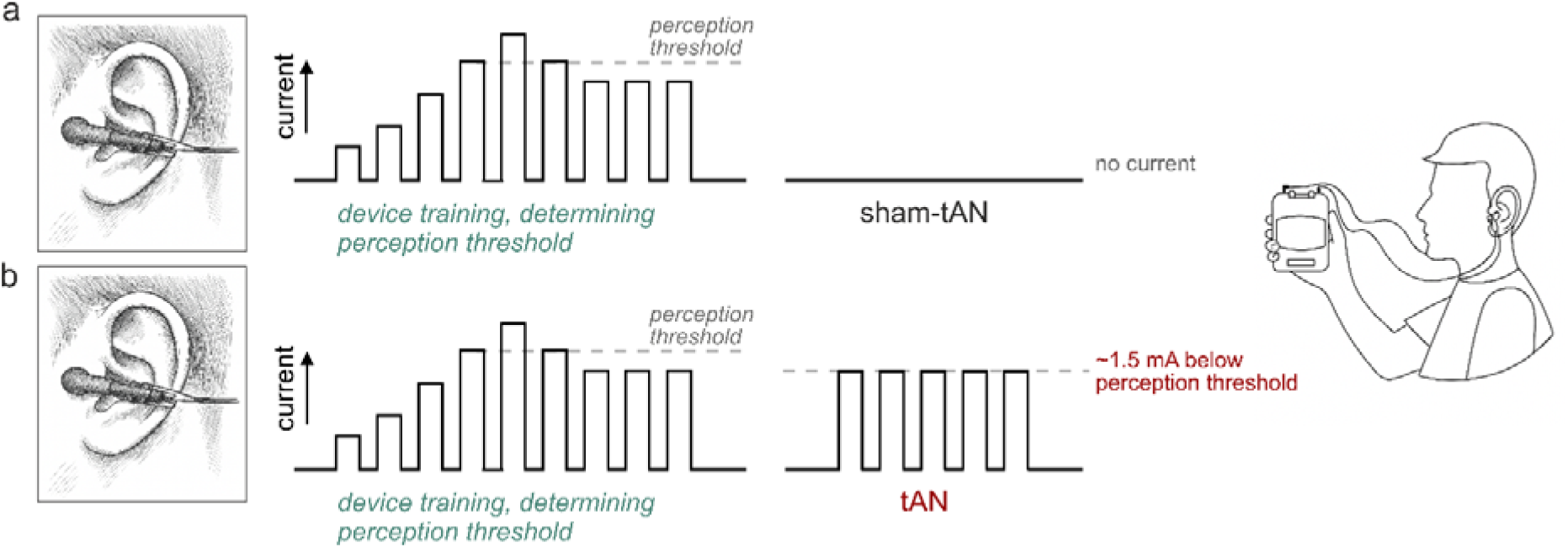
AffeX-CT device settings. The individual sensitivities of the auricular tragi regions to tAN stimulation were determined during the randomization visit using a dedicated training device. During device training, participants were instructed to place the electrode clips on the left and right tragi. The current amplitude was gradually increased by the investigator conducting the training, starting from 0.1 mA, until the participant felt a tingling sensation. The current was then reduced to set the level of stimulation at ∼1.5 mA below this threshold. Once this threshold current was determined, participants were issued their personal device units, identical to the training device but with concealed controls. The stimulation current was set to 0 mA (a; sham-tAN) or ∼1.5 mA below the individual perception threshold (b; tAN), with 200 μs pulses generated at a frequency of 30 Hz.

Participants were instructed to use the device independently at home, ideally setting aside 30 minutes in the evening when relaxed and not engaging in strenuous activity. Light activities such as reading or watching TV were recommended during stimulation. Before using the device, participants cleaned the outer ear. The device’s two ear clip leads (left and right) were moistened with a wet tissue at the contact points and attached to the left and right tragus, respectively. The device was then activated to stimulate the auricular branch of the vagus nerve.

For participants in the sham-tAN arm, the same training procedure was followed. However, the sham AffeX-CT device was programmed to deliver no stimulation current to the tragus. Otherwise, the sham device was identical in all other aspects. The participant, clinical research team, and data analysis team were all blinded to the intervention.

### Procedure

Trial participants were required to attend five visits spanning 12 weeks/84 days: visit 1 (screening), visit 2 (baseline and randomisation - Day 0), visit 3 at Day 14, visit 4 at Day 28 and visit 5 (End of Treatment - Day 84) (see Figure 2 for Flowchart). Participants also received several telephone calls: one between days 1-3, one at Day 7, one at Day 56 and one at Day 112 (post-trial follow up). Participants received text and/or email reminders at Day 42 and Day 70. The expected total duration of participation was 16 weeks. No changes in antihypertensive medications were permitted during the trial (between randomization on Day 0/visit 2 and end of treatment on Day 84/visit 5). Table 2 summarises the assessments and data collected at each visit with a full detailed schedule in Supplementary Table 4.

**Figure 2.**
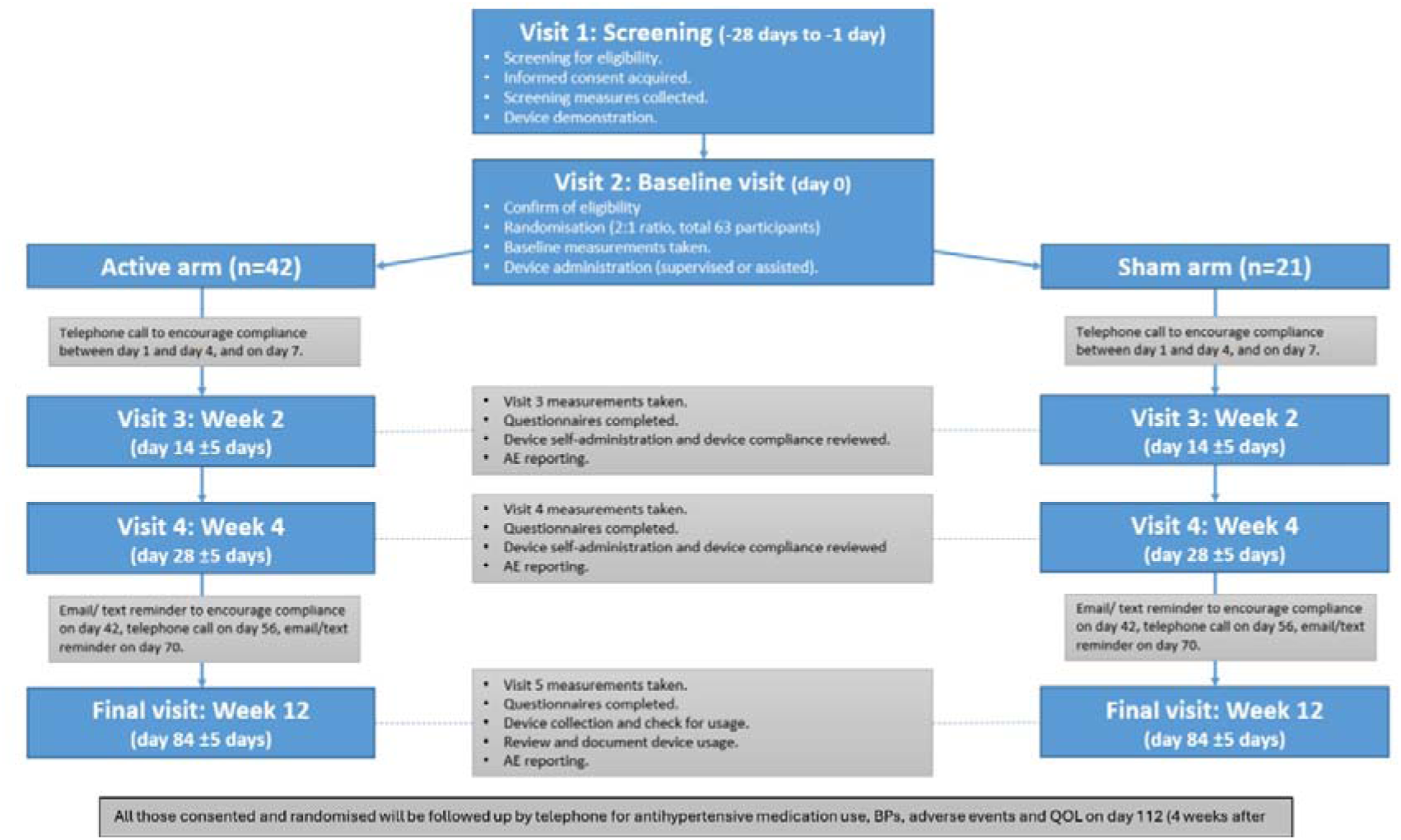
Flowchart for SCRATCH-HTN main trial.

**Table 2.**
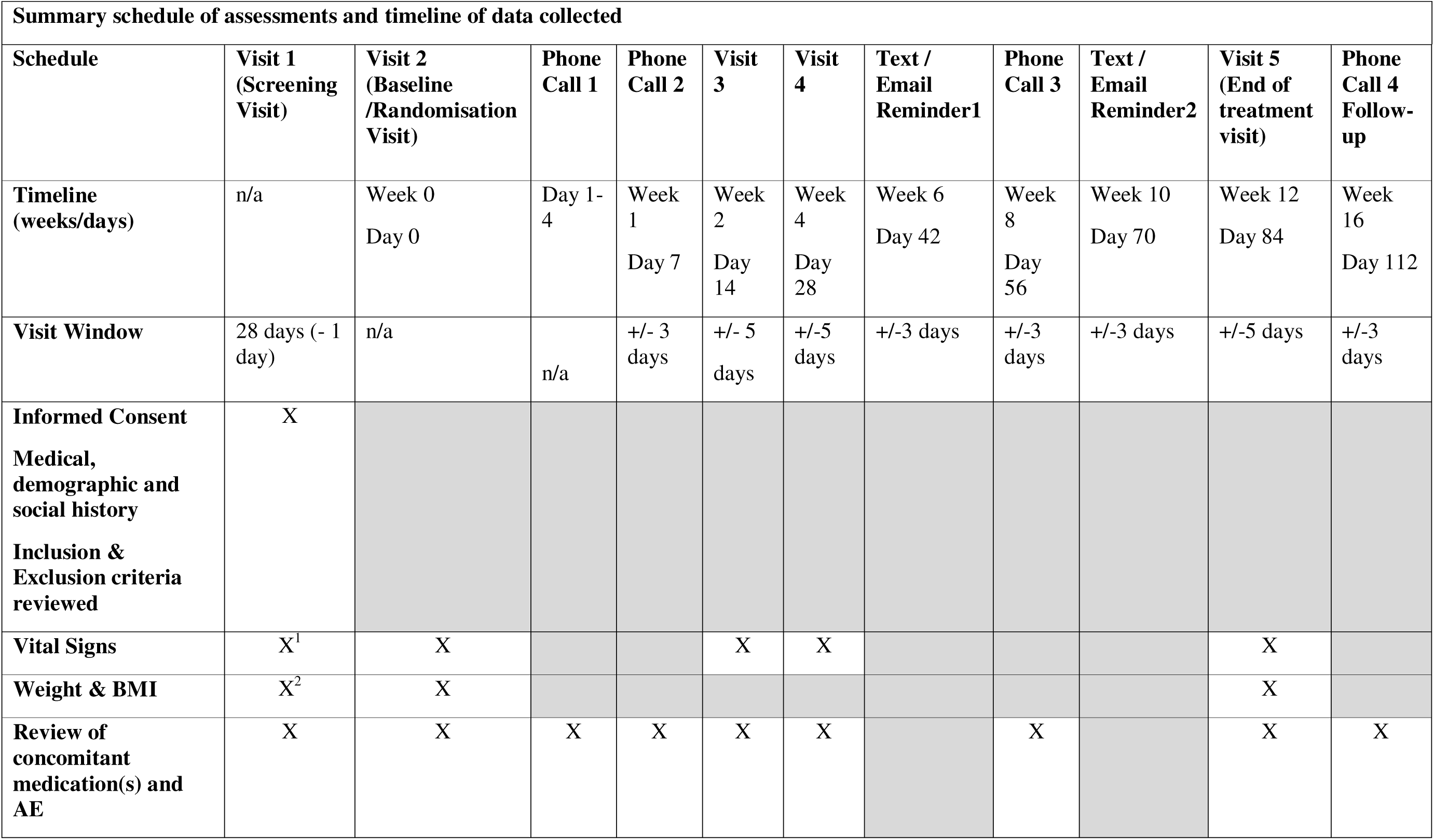
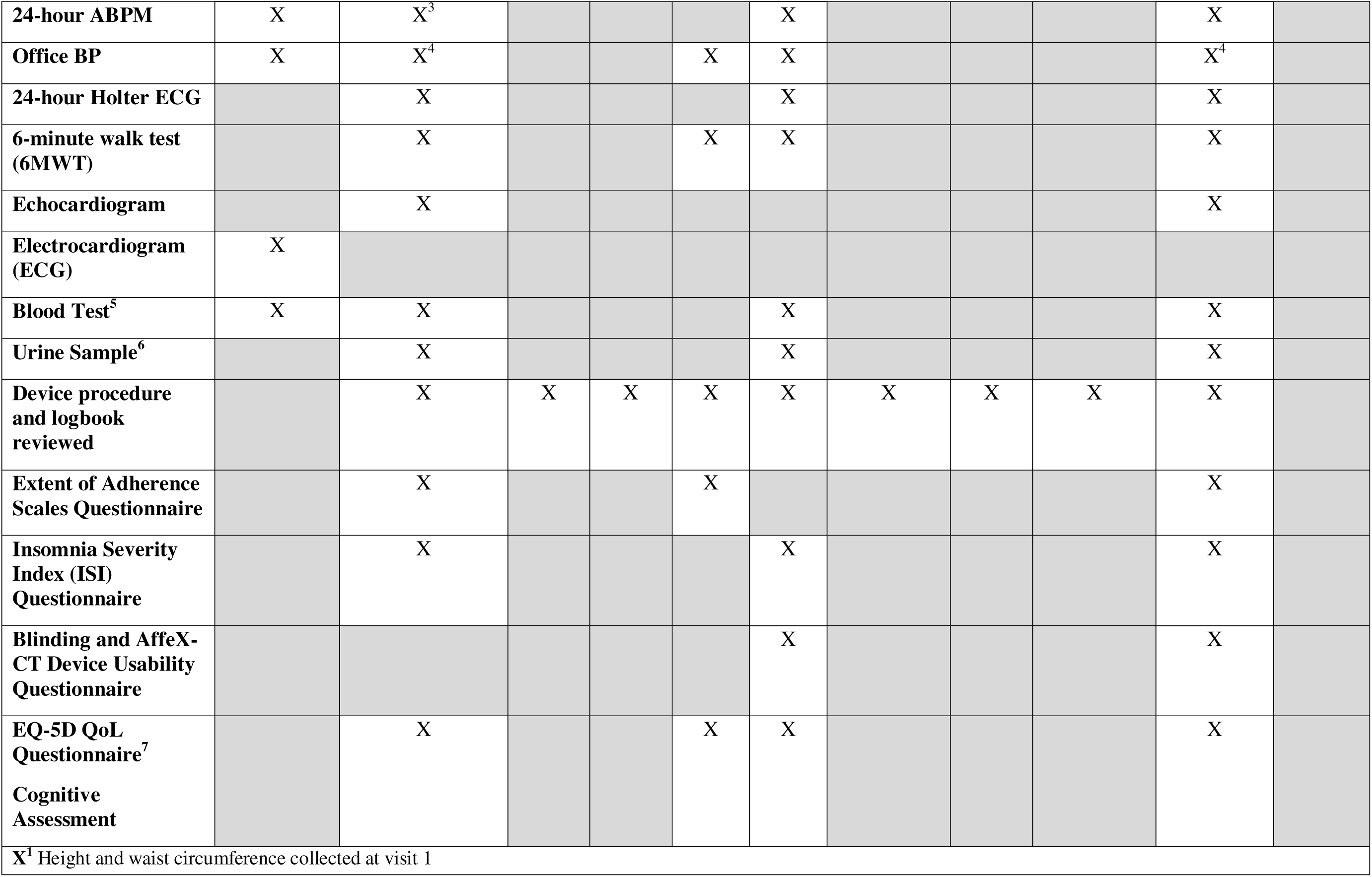
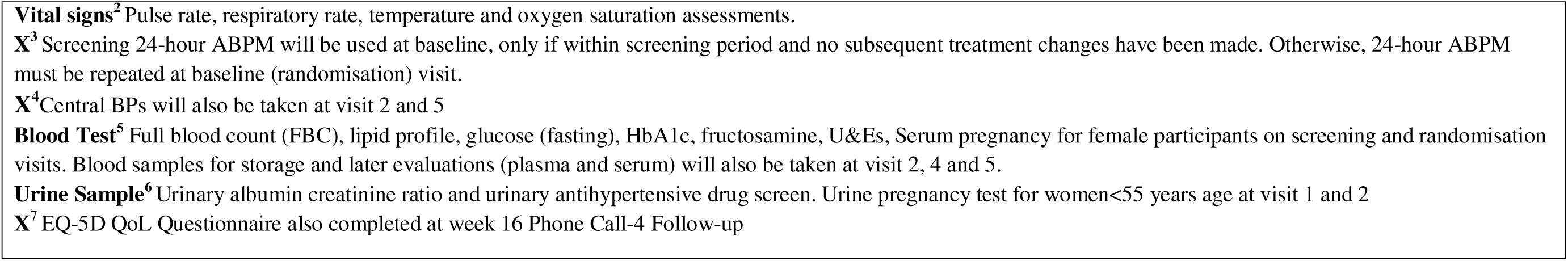
Summary schedule of assessments and timeline of data collected.

A maximum of 28 days was allowed between screening and randomization visit. At the screening visit (visit 1), all participants gave informed signed consent to enrol on the study, provide demographic information, medical and social history, and current concomitant medication. Vital signs, height & waist circumference, weight & BMI, ECG, 24-hour ABPM and office BP assessments were completed. Blood samples were taken from all participants and for those female participants <55 years of age, serum and urine pregnancy testing was completed. Inclusion and exclusion criteria were evaluated during the screening period. At the chief investigator’s (CI) judgement, the screening visit was allowed to be repeated once. Only after all screening assessments were completed, and if the participant was eligible and consented to the trial, were they invited to a baseline/randomisation, visit 2.

At baseline and randomisation (visit 2, day 0) participants were trained to use the device and provided with their personal AffeX-CT devices (including a user manual, see Supplementary Material 2 – User Guide), device logbook (in which they were instructed to record the date, time and duration of each stimulation session and the presence of any adverse effects) and participant ID card. Patients also underwent the following assessments: vital signs, weight and BMI, 24-hour ABPM, Office BP, Central BP, 24-hour Holter ECG, 6-minute walk test (6MWT), echocardiogram, blood tests (for pathology assessment and for storage and later evaluations), urine tests (for urinary albumin creatinine ratio and for urinary antihypertensive drug screening to objectively assess adherence to medications).The trial participants were trained how to use the device, prescribed an individualised stimulation intensity as described above, issued their assigned randomised device and asked to do the first self-stimulation under the direct supervision of the clinical research staff. Participants had their safety BPs recorded after 30 minutes of their first self-stimulation. The participant also completed three questionnaires (Extent of Adherence scale^39^, Insomnia Severity Index (ISI)^40^, EuroQol Five-Dimensional scale for Quality of Life (EQ-5D QoL))^41^ and a cognitive assessment (PEBL 2.0)^42^.

Participants were asked to self-administer the treatment using the device for 14 consecutive days for 30 minutes each evening. During this period, participants received two phone calls as reminders to apply treatment daily for the duration of the initial course of treatment and complete device procedure and device logbook entries. Concomitant medication and adverse events (AEs) were simultaneously reviewed.

After 14 days, visit 3 was conducted where medications, logbooks and AEs were reviewed. The participant also had office BPs, vital signs and 6MWT assessment and completed the cognitive assessment, adherence and QoL questionnaires. Participants were then asked to self-administer therapy once a week for a further 10 weeks.

During this time, visit 4 took place on day 28. At this visit, vital signs, 24-hour ABPM, office BP, 24-hour Holter ECG, 6MWT were assessed and a review of concomitant medication(s), use of the device and logbook and any AEs were completed. Participants had their blood and urine taken for safety and medication adherence assessments and also completed a cognitive assessment and questionnaires on device usability, QoL, insomnia and blinding. Email/text reminders to complete the device procedure and logbook entry were sent on day 42. A phone call was conducted on day 56 reminding participants to continue using the device and complete logbook entries and concomitant medication(s) and AEs were reviewed. Another email/text reminder was sent on Day 70.

The end of treatment visit 5 took place at the end of week 12 (day 84.) Participants completed the following assessments: vital signs, weight & BMI, 24-hour ABPM, office BP, central BP, 24-hour Holter ECG, 6MWT, echocardiogram, safety blood and urine samples. Concomitant medication(s), device logbooks and AEs were reviewed, and the device was returned. Participants then completed all trial questionnaires and the final cognitive assessment.

After the trial, participants were followed up at week 16 (day 112) with a phone call to review concomitant medication(s) and AEs, and the QoL was completed remotely.

In addition, we offered an extra-visit/clinical assessment as per the participants’ clinical need, particularly if BPs were either high (SBP >180 mmHg) or low (SBP <100 mmHg), or if they were symptomatic with postural hypotension or there were other clinical concerns.

### Outcomes

The primary endpoint was the change in average daytime ambulatory SBP from baseline to the end of treatment (3 months). Both assessments of average daytime ambulatory SBP were calculated using ABPM recordings between 7am and 11pm on the day of the assessment. Secondary endpoints are detailed in Supplementary Table 5 which include the change in 24-hour ambulatory and office SBP and DBP, ambulatory HR, BP variability, HR variability, quality of life, and sleep quality from baseline to end-of-treatment.

### Exploratory endpoints and sub-studies

The trial also collected data throughout the study (Supplementary Table 4) that will be used to assess various exploratory endpoints (see Supplementary Table 6 for details.) Amongst others, these include feasibility endpoints to assess the safety and acceptability of the device for patients in addition to echocardiographic markers and central BP measurements at baseline and end-of-treatment to explore the potential broader cardiovascular effects of tAN. Plasma and serum samples have also been stored at −80°C from blood samples collected at baseline, week 4 and week 12 visits for all patients for subsequent analysis of proteomics, inflammatory markers and metabolomics.

For a subgroup of consented patients (n=22), we completed a sub-study of several detailed physiological tests, commonly known as Autonomic Target-Organs Neurophysiological tests (ATONT), at baseline and week 12 (end of treatment), to explore the potential mechanism underlying the effects of tAN. ATONT takes approximately 90 minutes to complete, and includes continuous measurements of heart rate, beat-to-beat BP, tissue oxygenation, respiratory rate, and ECG monitoring. Participants are also asked to complete simple physiological manoeuvres: standing from lying position, hand-grip, sit up, Valsalva manoeuvre and carotid massage.

### Sample size

The trial was powered in relation to the primary endpoint (change in daytime ambulatory SBP between baseline and the end of the treatment at 12 weeks) in the active treatment arm. This was done using a paired t-test approach.

Using a conservative assumption of a mean change in SBP of 5.5 mmHg with a standard deviation of 11 mmHg, based on the existing data for hypertensive patients, 34 participants would be required to give 80% power to detect such a change at the two-sided alpha level of 0.05. After inflation for a potential 10% drop-out and a further 10% non-compliance level, we required 42 subjects in the intervention (tAN) arm.

The study also recruited participants to be randomised to the sham-tAN arm to compare changes in SBP in those undergoing treatment to consider potential Hawthorne and placebo effects. In order to collect more data regarding safety on the active treatment, the trial recruited double the number of subjects for the active treatment than for the sham treatment, a randomisation ratio of 2:1. Hence the sample size for the sham-tAN treatment was 21, giving a total sample size across both arms of 63 subjects.

With the sample size of 63 subjects and 2:1 randomisation ratio, a mean difference in SBP between the groups of 8.4 mmHg with no drop-out or non-compliance, or 9.3 mmHg if 20% of subjects drop-out or are non-compliant would be required, to detect the difference at the two-sided alpha level of 0.05, with 80% power.

### Statistical methods

Baseline characteristics data with demographic and physiological breakdown will be presented separately by trial group. The main analyses of the primary and secondary endpoints will be conducted on an intention-to-treat population (all randomised participants with available data irrespective of their compliance with the device) and on a per protocol population (all participants with treatment compliance on 80% or more days during the trial). Any participants who have a change in antihypertensive medication during the treatment period for safety/ethical reasons or otherwise will be analysed in the intention-to-treat but not in the per-protocol population.

The primary endpoint, the change in daytime ambulatory SBP, will be calculated for each participant by subtracting the end of treatment measurement (at 3 months) from the baseline measurement. The mean change in daytime SBP within each group will be presented with 95% confidence intervals (CIs) within the active treatment arm. The crude difference in the mean change in daytime SBP between treatment arms will be calculated between treatment arms with a 95% CI, and a two-sample t-test with equal variances will be used to test the null hypothesis of no difference in the change in SBP between groups.

Adjusted analysis will be conducted using linear regression comparing change in SBP between treatment groups while adjusting for baseline daytime ABPM SBP (ANCOVA), as well as adjusting for age, sex, and BMI. The estimated adjusted difference in change in SBP will be presented, along with 95% confidence intervals (CIs) and p-value. The primary endpoint will also be analysed according to subgroups (i) BMI at baseline (<30 kg/m2 vs. ≥30 kg/m2), (ii) diabetes status at baseline, (iii) age at baseline (<65 years vs. ≥65 years), mean ABPM daytime SBP at baseline (<160 mmHg vs. ≥160 mmHg), and a test for interaction will be conducted between groups for each of the 4 risk factors listed. Additionally, data from the urinary antihypertensive drug screening samples collected at visit 2, 4 and 5 that provide objective longitudinal data on antihypertensive medication adherence for each participant from the point of randomisation to end of treatment will be used to adjust the analysis of the primary outcome for partial or noncompliance to medications.

All secondary endpoints that are continuous will be analysed in the same way as for the primary endpoint. Binary endpoints will be shown as the number with each endpoint and total number in each group along with the percentage. For binary endpoints, the crude and adjusted odds ratios (ORs) will be estimated.

All feasibility endpoints for this phase 2a study will be presented descriptively or analysed qualitatively.

We will also summarise AEs using counts and percentages and present these overall for the study and by treatment arm. The number of subjects with AEs of mild/moderate/severe intensity will be shown overall and by treatment arm using the maximum severity experienced for each participant. The total number of AEs for each treatment, allowing multiple events per participant, will also be presented.

Serious AEs (SAEs; both non-fatal and fatal) will be listed separately along with details of the treatment and whether the event is unexpected or thought to be related to the treatment.

No interim analyses of any endpoint were planned. The full statistical analysis plan is provided in Supplemental Material 3 and full clinical investigational plan is provided in Supplemental Material 4.

## Discussion

The SCRATCH-HTN trial is a double-blind, sham-controlled study evaluating the safety and acceptability of tAN using the Affex-CT device and novel algorithm of stimulation for the first time in human subjects. The trial employs an improved protocol involving bilateral tragus stimulation. In addition to assessing safety, the study investigates the potential efficacy of tAN in reducing BP in patients with uncontrolled hypertension despite ongoing treatment. The findings are expected to pave the way for further evaluation of this innovative technology and establish it as a viable therapeutic option. Through the number of secondary and exploratory endpoints and the sub-study, the SCRATCH-HTN trial also intends to be hypothesis generating; by examining the effects of tAN on sleep quality, exercise tolerance, heart rate variability, central BP, cardiac structure, and autonomic function, the study aims to provide mechanistic insights into tAN and data on other related potential therapeutic benefits that might inform future studies. If effective, or if tAN demonstrates efficacy within the constraints of statistical power considerations, tAN could represent a transformative approach to hypertension management, fostering further research into this device and its applications in the field.

The study was carefully designed to mitigate known risks associated with device-based and non-pharmacological interventions in hypertension trials. The trial participants included were those with uncontrolled BP receiving medication, who were asked to avoid any changes in their medications. Whilst there is an increased risk of cardiovascular sequelae from uncontrolled hypertension, the trial duration was just 12 weeks, a shorter period than that which patients invariably wait between community or secondary care visits following a dose escalation. Those patients at higher risk with recent complications of hypertension such as stroke, myocardial infarction, or decompensated heart failure that require strict control of BP were excluded at screening from the trial. Moreover, we carefully monitored these patients during the trial period and if, in the unlikely case at any stage during the trial, their BPs were at levels that can potentially damage target organs acutely, for example, mean average office SBP > 180 mmHg or DBP > 120 mmHg, we allowed the introduction of another medication or escalation of existing antihypertensive treatment as per the current guidelines and their physician advice.

Participants in the device trials are often particularly motivated by the prospect of innovative treatments, which can introduce biases. To minimise this, all enrolled participants were already on established antihypertensive therapy at baseline. At both screening and randomisation, they were explicitly informed that their medication regimen must remain unchanged and be taken consistently throughout the study. To reinforce adherence and detect any compliance issues, multiple checks were implemented: verbal confirmation, self-reported data via participant device logbooks, and regular urinary screening for antihypertensive agents. These measures aimed both to support protocol fidelity and to capture any non-adherence.

Regarding potential risks directly related to the use of AffeX-CT device, there are no known serious risks of applying this form of autonomic neuromodulation. In a study of 6 healthy volunteers, no adverse effects of tAN administered using AffeX-CT device on heart rate, BP or ECG parameters (QT interval) were observed during 30 min of stimulation and for 60 min after the stimulation. No side effects and/or AEs were reported by the patients recruited in the proof-of-concept study (19 participants). Moreover, as for tAN more generally, systematic reviews and meta-analyses show that tAN/tVNS is a safe and well-tolerated treatment^43,44^. Incidence of AEs, in general, are low with one meta-analysis calculating 12.84/100,000 person-minutes-days of stimulation^44^. Another meta-analysis reported that out of 1,322 human subjects the most common side effects were local skin irritation from electrode placement (240 subjects, 18.2%), headache (47 subjects, 3.6%) and nasopharyngitis (23 subjects, 1.7%)^43^. The following side effects with frequency of <1% associated with tAN applied via electrical stimulation of the tragus may also be anticipated: light-headedness, fatigue/tiredness, mood changes, neck pain, tooth pain, pain/local skin irritation due to attachment of the device ear clips, tingling sensation due to the use of the device, and increased frequency of ventricular extrasystoles. Most of these side effects are transient and temporary in nature, and are likely to reduce in intensity and frequency over regular usage. Another challenge of this device trial is to maintain blinding amongst participants—particularly those in the active arm. Theoretically, they could self-adjust the stimulation level and may infer their group allocation. Our training protocol for the participants reduces this risk, as do constant reminders throughout the study period. A recent study using a similar device in healthy volunteers^45^ reported minimal risk of unblinding across two arms of the study. Additionally, in our study, we have collected specific data to assess whether perceptions of treatment assignment differed between groups. We will do a sensitivity analysis regarding that, if required.

If the findings of the proof-of-concept study are confirmed (reduction in SBP by >10 mmHg, lasting for >1 month after the discontinuation of therapy), tAN will prove to be more efficacious than renal denervation or the majority of single antihypertensive medications. The implication of lowering BP by 5-10 mmHg is significant. The largest and most detailed individual patient-level meta-analysis of data obtained in 348,854 participants from 48 randomised clinical trials (that evaluated the effects of BP -lowering treatments on the risk of major cardiovascular events and death in patients with and without cardiovascular disease) demonstrated that over an average of 4 years of follow-up, each 5 mmHg reduction in SBP lowered the relative risk of major cardiovascular events by ∼10%. The risks of stroke, ischaemic heart disease, heart failure and death from cardiovascular disease are reduced by 13%, 8%, 13% and 5%, respectively, with each 5 mmHg reduction in SBP ^46^. For 21 participants randomised to receive the sham-tAN treatment, we anticipate participation in the trial is still likely to be beneficial not only due to the Hawthorne effect but because of the overall care and comprehensive cardiovascular assessment that they receive in being a part of the trial.

Overall, this study is expected to provide evidence not only on the safety and acceptability of tAN, but also to lay the groundwork for a more physiological and mechanistic understanding of whether non-invasive autonomic neuromodulation via auricular stimulation can improve BP control, overall wellbeing, exercise tolerance, and other outcomes outlined in the primary and secondary objectives. Among these, evidence supporting a potential effect on BP control would be particularly significant, potentially paving the way for more in-depth investigations and offering a novel therapeutic option for people with uncontrolled hypertension.

## Ethics and Dissemination

The trial was registered in ClinicalTrials.gov (NCT05179343) and ISRCTN (14509154) and received Clinical Trial No Objection (CI/2021/0069/GB) from the Medicines and Healthcare products Regulatory Agency (MHRA), a favourable opinion from the West of Scotland NHS research ethics committee (21/WS/0157), and Health Research Authority Approval. After the publication of the main results and planned sub-studies, the data, analysis code and fully disclosed results (including those of the secondary, exploratory and sub-study outcomes) may be shared on reasonable request to the chief investigator to promote reproducibility and future collaborations.

Recruitment for the SCRATCH-HTN study was completed in May 2025. Final participant follow-ups are due to be completed in September 2025 with the analysis and subsequent presentation of results, in peer-reviewed scientific journals and at scientific conferences, anticipated in early 2026.

## Supporting information

AffeX-CT User Guide

Statistical Analysis Plan

Clinical Investigation Plan

## Data Availability

All data produced in the present work are contained in the manuscript.

## Collaborators

We acknowledge the support of Afferent Medical Solutions Ltd (Device Manufacturer) for this study, who have helped us with the device-related regulatory aspects.

## Acknowledgements

We thank all the patients and their families who have helped in completing this study. We also thank all of the clinicians and allied health professionals (particular the cardiac diagnostics department) at St Bartholomew’s Hospital and Royal London Hospital (particularly from the BP Centre of Excellence). We thank the Barts Cardiovascular Clinical Trials Unit (CVCTU), a branch of the Barts CTU UKCRC Reg No. 4, and acknowledge the Sponsor, Queen Mary University of London. We also thank the NIHR Barts Biomedical Research Centre for their support. We also acknowledge the support of NIHR Be Part Of Research Volunteer Service (BPORVS), who were key to achieving our recruitment target.

## SCRATCH HTN Study investigators

### 1. SCRATCH-HTN investigators

**William Harvey Research Centre, Queen Mary University of London, and Barts Health NHS Trust:** Ajay Gupta, Jane Field, David Collier, James Steckelmacher, Olivier Zongo, Mital Patel, Patrizia Ebano, Manish Saxena, Abubaker Eltayeb, Annastazia Learoyd, Georgia Mannion-Krase, Clovel David, Mussadiq Shah, Peter Julu, Amrita Ahluwalia.

**Imaging team**: Jing Deng, James Malcolmson, Sanjeev Bhattacharyya.

**PIC sites investigators: Royal Free London NHS Foundation Trust**: Alfredo Petrosino (PI), Stephen Walsh* and Manoj Makharia; **University College London Hospitals NHS Foundation Trust**: Marc George (PI) and Mollie Little; **St George’s University Hospitals NHS Foundation Trust:** Teck Khong (PI); **Homerton University Hospital NHS Foundation Trust:** Louise Abram (PI); **Broomfield Hospital, Mid and South Essex Hospitals NHS Foundation Trust**: Tehreem Butt (PI), Rachael Arnold and Caroline Mitchell; **Imperial College Healthcare NHS Trust:** Neil Poulter (PI) and Farhat Ghafoor.

* Posthumous

### 2. SCRATCH HTN Trial Committees

**Trial Management Committee (TMG):** Ajay Gupta (chair), David Collier, James Steckelmacher, Jane Field, Olivier Zongo, Mital Patel, George Collett, Annastazia Learoyd, Abubaker Eltayeb, Manish Saxena, Georgia Mannion-Krase, Patrizia Ebano.

**Trial Steering Committee (TSC):** Peter Sever (chair), Ian Wilkinson, Rasha Al-Lamee, Ken Peters, Peter Davison, Ajay Gupta, David Collier, Annastazia Learoyd, Alexander Gourine, Everard Mascarenhas, Lucia Bianchi, Stuart Haylock, Jane Field.

**Data Safety Monitoring Committee (DSMC):** Neil Chapman (chair), Vikas Kapil, Tim Collier, Ajay Gupta, Kamran Khan, Annastazia Learoyd, Jane Field, Raj Mehta.

**Research Steering Group (RSG):** Ajay Gupta (chair), Paul Robinson, Gareth Ackland, Alexander Gourine, Everard Mascarenhas, Rayan Altayeb, Mamta Bajre, Jane Field.

## References

1. WHO. Global report on hypertension: the race against a silent killer. In: World Health Organization; 2023.

2. Collaborators GBDRF. Global burden and strength of evidence for 88 risk factors in 204 countries and 811 subnational locations, 1990-2021: a systematic analysis for the Global Burden of Disease Study 2021. Lancet. 2024;403:2162–2203. doi: 10.1016/S0140-6736(24)00933-4

3. Collaborators GBDCoD. Global burden of 288 causes of death and life expectancy decomposition in 204 countries and territories and 811 subnational locations, 1990-2021: a systematic analysis for the Global Burden of Disease Study 2021. Lancet. 2024;403:2100–2132. doi: 10.1016/S0140-6736(24)00367-2

4. Collaborators GBDRF. Global burden of 87 risk factors in 204 countries and territories, 1990-2019: a systematic analysis for the Global Burden of Disease Study 2019. Lancet. 2020;396:1223–1249. doi: 10.1016/S0140-6736(20)30752-2

5. Collaboration GBDCKD. Global, regional, and national burden of chronic kidney disease, 1990-2017: a systematic analysis for the Global Burden of Disease Study 2017. Lancet. 2020;395:709-733. doi: 10.1016/S0140-6736(20)30045-3

6. Collaborators GBDS. Global, regional, and national burden of stroke and its risk factors, 1990-2019: a systematic analysis for the Global Burden of Disease Study 2019. Lancet Neurol. 2021;20:795–820. doi: 10.1016/S1474-4422(21)00252-0

7. Mensah GA, Fuster V, Murray CJL, Roth GA, Global Burden of Cardiovascular D, Risks C. Global Burden of Cardiovascular Diseases and Risks, 1990-2022. J Am Coll Cardiol. 2023;82:2350–2473. doi: 10.1016/j.jacc.2023.11.007

8. Statistics OfN. Health Survey for England 2021, part 2. In: NHS Digital; 2023.

9. Graham C SJ, Prashar J, et al.. Trends in hypertension prevalence, control and antihypertensive use in England over the last 2 decades: insights from annual, nationwide Health Surveys for England from 2003 to 2021. BMJ Medicine. 2025.

10. England PH. Tackling high blood pressure: from evidence into action. In: Public Health England; 2014.

11. Lee EKP, Poon P, Yip BHK, Bo Y, Zhu MT, Yu CP, Ngai ACH, Wong MCS, Wong SYS. Global Burden, Regional Differences, Trends, and Health Consequences of Medication Nonadherence for Hypertension During 2010 to 2020: A Meta-Analysis Involving 27 Million Patients. J Am Heart Assoc. 2022;11:e026582. doi: 10.1161/JAHA.122.026582

12. Abegaz TM, Shehab A, Gebreyohannes EA, Bhagavathula AS, Elnour AA. Nonadherence to antihypertensive drugs: A systematic review and meta-analysis. Medicine (Baltimore). 2017;96:e5641. doi: 10.1097/MD.0000000000005641

13. Bourque G, Ilin JV, Ruzicka M, Hundemer GL, Shorr R, Hiremath S. Nonadherence Is Common in Patients With Apparent Resistant Hypertension: A Systematic Review and Meta-analysis. Am J Hypertens. 2023;36:394–403. doi: 10.1093/ajh/hpad013

14. Dahal K, Khan M, Siddiqui N, Mina G, Katikaneni P, Modi K, Azrin M, Lee J. Renal Denervation in the Management of Hypertension: A Meta-Analysis of Sham-Controlled Trials. Cardiovasc Revasc Med. 2020;21:532–537. doi: 10.1016/j.carrev.2019.07.012

15. Gupta A, Prince M, Bob-Manuel T, Jenkins JS. Renal denervation: Alternative treatment options for hypertension? Prog Cardiovasc Dis. 2020;63:51–57. doi: 10.1016/j.pcad.2019.12.007

16. Kandzari DE, Bohm M, Mahfoud F, Townsend RR, Weber MA, Pocock S, Tsioufis K, Tousoulis D, Choi JW, East C, et al. Effect of renal denervation on blood pressure in the presence of antihypertensive drugs: 6-month efficacy and safety results from the SPYRAL HTN-ON MED proof-of-concept randomised trial. Lancet. 2018;391:2346–2355. doi: 10.1016/S0140-6736(18)30951-6

17. Mufarrih SH, Qureshi NQ, Khan MS, Kazimuddin M, Secemsky E, Bloch MJ, Giri J, Cohen D, Swaminathan RV, Feldman DN, et al. Randomized Trials of Renal Denervation for Uncontrolled Hypertension: An Updated Meta-Analysis. J Am Heart Assoc. 2024;13:e034910. doi: 10.1161/JAHA.124.034910

18. Farmer AD, Strzelczyk A, Finisguerra A, Gourine AV, Gharabaghi A, Hasan A, Burger AM, Jaramillo AM, Mertens A, Majid A, et al. International Consensus Based Review and Recommendations for Minimum Reporting Standards in Research on Transcutaneous Vagus Nerve Stimulation (Version 2020). Front Hum Neurosci. 2020;14:568051. doi: 10.3389/fnhum.2020.568051

19. Clancy JA, Mary DA, Witte KK, Greenwood JP, Deuchars SA, Deuchars J. Non-invasive vagus nerve stimulation in healthy humans reduces sympathetic nerve activity. Brain Stimul. 2014;7:871–877. doi: 10.1016/j.brs.2014.07.031

20. Yuan H, Silberstein SD. Vagus Nerve and Vagus Nerve Stimulation, a Comprehensive Review: Part II. Headache. 2016;56:259–266. doi: 10.1111/head.12650

21. Yuan H, Silberstein SD. Vagus Nerve and Vagus Nerve Stimulation, a Comprehensive Review: Part III. Headache. 2016;56:479–490. doi: 10.1111/head.12649

22. Yuan H, Silberstein SD. Vagus Nerve and Vagus Nerve Stimulation, a Comprehensive Review: Part I. Headache. 2016;56:71–78. doi: 10.1111/head.12647

23. Bretherton B, Atkinson L, Murray A, Clancy J, Deuchars S, Deuchars J. Effects of transcutaneous vagus nerve stimulation in individuals aged 55 years or above: potential benefits of daily stimulation. Aging (Albany NY). 2019;11:4836–4857. doi: 10.18632/aging.102074

24. Frangos E, Ellrich J, Komisaruk BR. Non-invasive Access to the Vagus Nerve Central Projections via Electrical Stimulation of the External Ear: fMRI Evidence in Humans. Brain Stimul. 2015;8:624–636. doi: 10.1016/j.brs.2014.11.018

25. Garcia RG, Lin RL, Lee J, Kim J, Barbieri R, Sclocco R, Wasan AD, Edwards RR, Rosen BR, Hadjikhani N, et al. Modulation of brainstem activity and connectivity by respiratory-gated auricular vagal afferent nerve stimulation in migraine patients. Pain. 2017;158:1461–1472. doi: 10.1097/j.pain.0000000000000930

26. Kraus T, Kiess O, Hosl K, Terekhin P, Kornhuber J, Forster C. CNS BOLD fMRI effects of sham-controlled transcutaneous electrical nerve stimulation in the left outer auditory canal - a pilot study. Brain Stimul. 2013;6:798–804. doi: 10.1016/j.brs.2013.01.011

27. Antonino D, Teixeira AL, Maia-Lopes PM, Souza MC, Sabino-Carvalho JL, Murray AR, Deuchars J, Vianna LC. Non-invasive vagus nerve stimulation acutely improves spontaneous cardiac baroreflex sensitivity in healthy young men: A randomized placebo-controlled trial. Brain Stimul. 2017;10:875–881. doi: 10.1016/j.brs.2017.05.006

28. De Couck M, Cserjesi R, Caers R, Zijlstra WP, Widjaja D, Wolf N, Luminet O, Ellrich J, Gidron Y. Effects of short and prolonged transcutaneous vagus nerve stimulation on heart rate variability in healthy subjects. Auton Neurosci. 2017;203:88–96. doi: 10.1016/j.autneu.2016.11.003

29. Yang H, Shi W, Fan J, Wang X, Song Y, Lian Y, Shan W, Wang Q. Transcutaneous Auricular Vagus Nerve Stimulation (ta-VNS) for Treatment of Drug-Resistant Epilepsy: A Randomized, Double-Blind Clinical Trial. Neurotherapeutics. 2023;20:870–880. doi: 10.1007/s13311-023-01353-9

30. He W, Jing X, Wang X, Rong P, Li L, Shi H, Shang H, Wang Y, Zhang J, Zhu B. Transcutaneous auricular vagus nerve stimulation as a complementary therapy for pediatric epilepsy: a pilot trial. Epilepsy Behav. 2013;28:343–346. doi: 10.1016/j.yebeh.2013.02.001

31. Zamotrinsky A, Afanasiev S, Karpov RS, Cherniavsky A. Effects of electrostimulation of the vagus afferent endings in patients with coronary artery disease. Coron Artery Dis. 1997;8:551–557.

32. Stavrakis S, Stoner JA, Humphrey MB, Morris L, Filiberti A, Reynolds JC, Elkholey K, Javed I, Twidale N, Riha P, et al. TREAT AF (Transcutaneous Electrical Vagus Nerve Stimulation to Suppress Atrial Fibrillation): A Randomized Clinical Trial. JACC Clin Electrophysiol. 2020;6:282–291. doi: 10.1016/j.jacep.2019.11.008

33. Stavrakis S, Humphrey MB, Scherlag BJ, Hu Y, Jackman WM, Nakagawa H, Lockwood D, Lazzara R, Po SS. Low-level transcutaneous electrical vagus nerve stimulation suppresses atrial fibrillation. J Am Coll Cardiol. 2015;65:867–875. doi: 10.1016/j.jacc.2014.12.026

34. Mbikyo MB, Wang A, Ma Q, Miao L, Cui N, Yang Y, Fu H, Sun Y, Li Z. Low-Level Tragus Stimulation Attenuates Blood Pressure in Young Individuals With Hypertension: Results From a Small-Scale Single-Blind Controlled Randomized Clinical Trial. Journal of the American Heart Association. 2024;13. doi: 10.1161/jaha.123.032269

35. Stavrakis S, Elkholey K, Morris L, Niewiadomska M, Asad ZUA, Humphrey MB. Neuromodulation of Inflammation to Treat Heart Failure With Preserved Ejection Fraction: A Pilot Randomized Clinical Trial. Journal of the American Heart Association. 2022;11. doi: 10.1161/jaha.121.023582

36. Hays SA, Rennaker RL, Kilgard MP. Targeting Plasticity with Vagus Nerve Stimulation to Treat Neurological Disease. In: Changing Brains - Applying Brain Plasticity to Advance and Recover Human Ability. 2013:275–299.

37. Kwong PWH, Ng GYF, Chung RCK, Ng SSM. Bilateral Transcutaneous Electrical Nerve Stimulation Improves Lower-Limb Motor Function in Subjects With Chronic Stroke: A Randomized Controlled Trial. Journal of the American Heart Association. 2018;7. doi: 10.1161/jaha.117.007341

38. Kuznetsova OM, Tymofyeyev Y. Preserving the allocation ratio at every allocation with biased coin randomization and minimization in studies with unequal allocation. Stat Med. 2012;31:701–723. doi: 10.1002/sim.4447

39. Voils CI, Maciejewski ML, Hoyle RH, Reeve BB, Gallagher P, Bryson CL, Yancy WS, Jr. Initial validation of a self-report measure of the extent of and reasons for medication nonadherence. Med Care. 2012;50:1013–1019. doi: 10.1097/MLR.0b013e318269e121

40. Bastien CH, Vallieres A, Morin CM. Validation of the Insomnia Severity Index as an outcome measure for insomnia research. Sleep Med. 2001;2:297–307. doi: 10.1016/s1389-9457(00)00065-4

41. Balestroni G, Bertolotti G. [EuroQol-5D (EQ-5D): an instrument for measuring quality of life]. Monaldi Arch Chest Dis. 2012;78:155–159. doi: 10.4081/monaldi.2012.121

42. Mueller ST, Piper BJ. The Psychology Experiment Building Language (PEBL) and PEBL Test Battery. J Neurosci Methods. 2014;222:250–259. doi: 10.1016/j.jneumeth.2013.10.024

43. Redgrave J, Day D, Leung H, Laud PJ, Ali A, Lindert R, Majid A. Safety and tolerability of Transcutaneous Vagus Nerve stimulation in humans; a systematic review. Brain Stimul. 2018;11:1225–1238. doi: 10.1016/j.brs.2018.08.010

44. Kim AY, Marduy A, de Melo PS, Gianlorenco AC, Kim CK, Choi H, Song JJ, Fregni F. Safety of transcutaneous auricular vagus nerve stimulation (taVNS): a systematic review and meta-analysis. Sci Rep. 2022;12:22055. doi: 10.1038/s41598-022-25864-1

45. Ackland GL, Patel ABU, Miller S, Gutierrez del Arroyo A, Thirugnanasambanthar J, Ravindran JI, Schroth J, Boot J, Caton L, Mein CA, et al. Non-invasive vagus nerve stimulation and exercise capacity in healthy volunteers: a randomized trial. European Heart Journal. 2025;46:1634–1644. doi: 10.1093/eurheartj/ehaf037

46. Blood Pressure Lowering Treatment Trialists C. Pharmacological blood pressure lowering for primary and secondary prevention of cardiovascular disease across different levels of blood pressure: an individual participant-level data meta-analysis. Lancet. 2021;397:1625–1636. doi: 10.1016/S0140-6736(21)00590-0

