## Supplementary Tables and Figures for "A Sham Controlled Randomised Trial evaluating the Safety, Acceptability and Efficacy of Autonomic neuromodulation using Trans-Cutaneous vagal sensory stimulation in uncontrolled Hypertensive patients: Rationale and study design of the SCRATCH-HTN Study"

**Supplementary Materials**

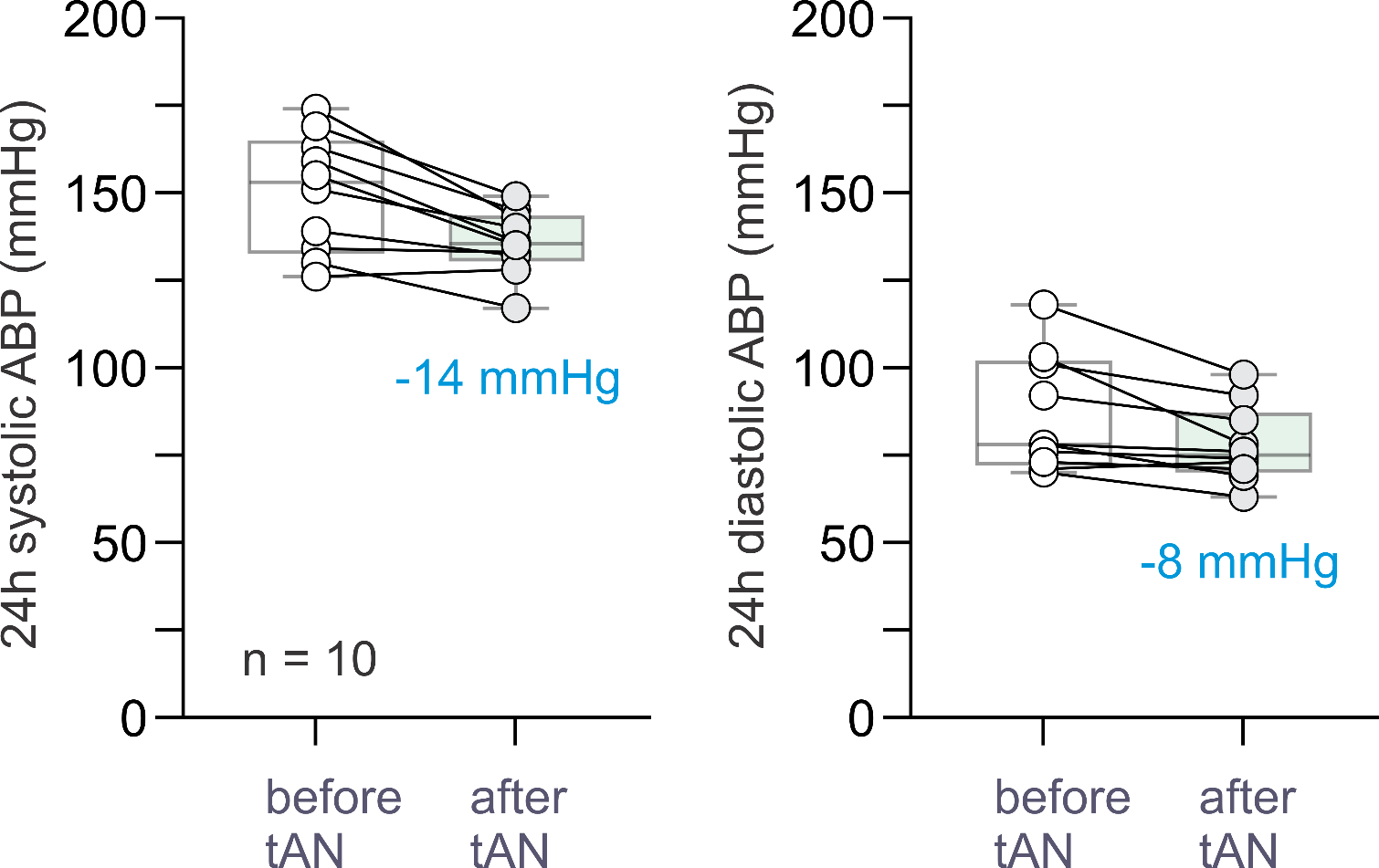
**Supplementary Figure 1.** The effect of tAN on 24-h systolic and diastolic blood pressure in drug-resistant (n=10) patients with systemic arterial hypertension. Data shown are before and one month after the course of tAN treatment (results of unpublished proof-of-concept study).

**Supplementary Table 1.** List of Patient Identification Centres (PIC).

**Patient Identification Centres (PIC)**

Imperial College Healthcare NHS Trust

St George’s University Hospitals NHS Foundation Trust

Homerton University Hospital NHS Foundation Trust

University College London Hospitals NHS Foundation Trust

Royal Free London NHS Foundation Trust

Broomfield Hospital, Mid and South Essex Hospitals NHS Foundation Trust

**Supplementary Table 2.** The inclusion criteria for SCRATCH-HTN.

**SCRATCH-HTN Inclusion Criteria.**

1.	Participant has given written informed consent.

2.	Participant has sufficient knowledge of the English language to be able understand the participant information sheet and trial materials including outcome assessments.

3.	Participant is aged ≥18 years and <80 years at the time of screening visit.

4.	Participant is taking between 1 to 4 antihypertensive medications (inclusive) at time of screening and baseline (randomisation) visit and is willing to adhere to no change in medication during the trial until end of the trial visit (visit 5). (NB. Participant on only one antihypertensive medication should be taking that medication for at least six weeks prior to the screening visit).

5.	Participant has confirmed diagnosis of hypertension.

6.	Participant meets BP criteria:

- 24-hour ambulatory BP monitoring (ABPM) at either screening visit or baseline (randomisation) visit, with mean daytime systolic BP of ≥135 mmHg and <170 mmHg and mean daytime DBP of ≥85 mm Hg and <115 mmHg (N.B. By default, Ambulatory Blood Pressure Monitoring [ABPM] at screening visit will be used at baseline visit. However, if there has been an addition of new medication after participants screening visit, 24-hour ABPM must be repeated at baseline visit, but the screening ABPM will be used for eligibility criteria)..

7.	Participant has one or more of the following associated conditions:

- Obesity: body mass index (BMI >30 or waist circumference >94 cm (men) or > 80cm (women). (NB. For participants of South-East Asian/Chinese/Japanese origin these cut-offs are >90 cm (men) or >80 cm (women)).
- Type 2 diabetes – controlled or sub-optimally controlled (HbA1c ≤8.5% or ≤69 mmol/mol) on diet and/ or medications except insulin.
- Heart rate (average) ≥70 bpm at screening or baseline (randomisation) visit (measurement taken after 5 minutes of rest in a seated position and when finger probe has been placed for a minimum of 30 seconds thereafter) or a heart rate (average) ≥60 bpm at screening or baseline (randomisation) visit if the patient is taking beta-blocker medication or a rate-limiting calcium channel-blocker medication.
- HbA1c ≥42 mmol/mol or fasting blood glucose (if available) ≥5.6 mmol/L AND either low HDL cholesterol (≤1.03 mmol/L for men and ≤1.29 mmol/L for women) or high triglyceride (triglycerides ≥1.7 mmol/L).
- Both low HDL cholesterol (≤1.03 mmol/L for men and ≤1.29 mmol/L for women) AND high triglyceride (triglycerides ≥1.7 mmol/L).
- Diagnosed or known case of polycystic ovarian syndrome.

8.	Female participants of child-bearing potential (all those <55 years except if they are surgically sterile, meaning they have undergone a hysterectomy, bilateral tubal ligation, or bilateral oophorectomy, or formally diagnosed by their doctors to be post-menopausal) must agree to use the acceptable methods of contraception from the time of consent until last follow up visit.

9.	Participant can communicate satisfactorily with the Investigator and Investigation Site staff, and to participate in, and comply with all clinical study requirements.

10.	Participant agrees to have all trial procedures performed and is able and willing to comply with all trial visits and protocol requirements.

**Supplementary Table 3.** The exclusion criteria for SCRATCH-HTN.

**SCRATCH-HTN Exclusion Criteria.**

1.	Participant is unable and unwilling to use the AffeX-CT device daily.

2.	Participant has a small tragus (ie. the size or shape of the tragus is such that it doesn’t allow the application of the ear-clips of the AffeX-CT device for a sustained period of time).

3.	Participant has a piercing on the tragus of the ear.

4.	Participant is diagnosed with atrial fibrillation or other form of cardiac arrhythmia

5.	Participant has eGFR <45 ml/min/1.73 m2 at screening visit.

6.	Participant has type 1 diabetes mellitus.

7.	Participant has type 2 diabetes mellitus on Insulin or those on oral antidiabetic medications with poor glycaemic control defined as HbA1c above 8.5% (or >69 mmol/mol).

8.	Participant has a history of falls or symptoms of orthostatic hypotension in the last 3 months prior to baseline (randomisation) visit.

9.	Participant is pregnant, nursing or planning to become pregnant within the next 6 months.

10.	Participant suffers from chronic pain and has taken anti-inflammatory drugs for two or more days per week over the last month prior to baseline (randomisation) visit.

11.	Participant has clinically significant or symptomatic hypertension-mediated target organ damage such as severe heart failure with NYHA 4, end stage renal damage, medically diagnosed/imaging proven stroke, symptomatic peripheral vascular disease, or severe retinopathy.

12.	Participant has a history of stable or unstable angina or had an acute coronary event within 3 months prior to baseline (randomisation) visit or had a myocardial infarction within the last six months of enrolment prior to baseline (randomisation) visit.

13.	Participant has history of renal denervation within 1 year prior to baseline (randomisation) visit.

14.	Participant has a therapeutic implantable electronic/electrical device such as pacemaker, implantable cardioverter-defibrillators (ICDs), implanted vagal stimulators.

15.	Participant has history of hospitalization (>24 hours) for heart failure or cerebrovascular accidents, or a history of stroke diagnosed based on imaging or evidence of specialist diagnosis or any other indirect evidence such as discharge summary or clinical letter (at any time in the past)

16.	Participant has mean daytime ABPM pulse pressure ≥80 mmHg at screening or baseline (randomisation) visit.

17.	Participant has a heart rate <50 bpm at screening or baseline (randomisation) visit (measurement taken after 5 minutes of rest in a seated position and when finger probe has been placed for a minimum of 30 seconds thereafter).

18.	Participant has auricular dermatitis.

19.	Participant has postural hypotension, defined as a fall >20mmHg in systolic BP on standing at 3 minutes (compared with sitting).

20.	Participant has a history of hospitalization for hypertensive emergency or urgency in the last six months of enrolment prior to baseline (randomisation) visit. Hospitalisation’ is defined as admission for more than 24 hours or between 12-24 hours with an overnight stay.

21.	Participant is identified as unsuitable to participate by the CI/Co-Investigator(s) and/or Investigation site team for another reason (e.g., for other medical reasons, laboratory abnormalities, limited life expectancy, etc.).

22.	Participants with history of epilepsy and are currently on anti-epileptic medication or those who are not on any anti-epileptic medication but have history of a seizure within last 10 years

| **Supplementary Table 4.** Schedule of assessments and timeline of data collected. | | | | | | | | | | | |
| --- | --- | --- | --- | --- | --- | --- | --- | --- | --- | --- | --- |
| **Schedule of assessments and timeline of data collected** | | | | | | | | | | | |
| **Schedule** | **Visit 1 (Screening Visit)** | **Visit 2 ** (Baseline /Randomisation Visit)** | **Phone Call-1** | **Phone Call-2** | **Visit 3**** | **Visit 4**** | **Text / Email Reminder -1** | **Phone Call-3** | **Text / Email Reminder-2** | **Visit 5** (End of treatment visit)** | **Phone Call-4 Follow-up*** |
| **Timeline (weeks/days)** | n/a | Week 0  Day 0 | Day 1-4 | Week 1  Day 7 | Week 2  Day 14 | Week 4  Day 28 | Week 6  Day 42 | Week 8  Day 56 | Week 10  Day 70 | Week 12  Day 84 | Week 16  Day 112 |
| **Visit Window** | 28 days (- 1 day) | n/a | n/a | +/- 3 days | +/- 5  days | +/-5 days | +/-3 days | +/-3 days | +/-3 days | +/-5 days | +/-3 days |
| **Informed Consent** | X |  |  |  |  |  |  |  |  |  |  |
| **Medical History, including demographic information and social history** | X | X^1^ |  |  |  |  |  |  |  |  |  |
| **Vital Signs^2^** | X | X |  |  | X | X |  |  |  | X |  |
| **Height & Waist Circumference** | X |  |  |  |  |  |  |  |  |  |  |
| **Weight & BMI** | X | X |  |  |  |  |  |  |  | X |  |
| **Concomitant Medication** | X | X | X | X | X | X |  | X |  | X | X |
| **24-hour ABPM** | X | X^3^ |  |  |  | X |  |  |  | X |  |
| **Office BP^4^** | X | X |  |  | X | X |  |  |  | X |  |
| **Central BP**^†^ |  | X |  |  |  |  |  |  |  | X |  |
| **24-hour Holter ECG** |  | X*** |  |  |  | X |  |  |  | X**** |  |
| **6-minute walk test (6MWT)** |  | X |  |  | X | X |  |  |  | X |  |
| **Echocardiogram** |  | X*** |  |  |  |  |  |  |  | X**** |  |
| **Electrocardiogram (ECG)** | X |  |  |  |  |  |  |  |  |  |  |
| **Blood Test^5^** | X | X |  |  |  | X |  |  |  | X |  |
| **Blood Samples for storage and later evaluations (Plasma & Serum)** |  | X |  |  |  | X |  |  |  | X |  |
| **Urine Pregnancy** | X | X |  |  |  |  |  |  |  |  |  |
| **Urine Sample^6^** |  | X |  |  |  | X |  |  |  | X |  |
| **ATONT Assessment**^†^ |  | X^7^ |  |  |  |  |  |  |  | X^8^ |  |
| **Inclusion and Exclusion** | X | X^9^ |  |  |  |  |  |  |  |  |  |
| **Randomisation** |  | X |  |  |  |  |  |  |  |  |  |
| **Device procedure and logbook** |  | X^10^ | X^11^ | X^11^ | X^12^ | X^12^ | X | X^11^ | X^11^ | X^12, 13^ |  |
| **Extent of Adherence Scales Questionnaire** |  | X |  |  | X |  |  |  |  | X |  |
| **Insomnia Severity Index (ISI) Questionnaire** |  | X |  |  |  | X |  |  |  | X |  |
| **Blinding Questionnaire** |  |  |  |  |  | X |  |  |  | X |  |
| **AffeX-CT** **Device Usability Questionnaire** |  |  |  |  |  | X |  |  |  | X |  |
| **EQ-5D QoL Questionnaire** |  | X |  |  | X | X |  |  |  | X | X |
| **Cognitive Assessment** |  | X |  |  | X | X |  |  |  | X |  |
| **AE Reporting** | X | X | X | X | X | X |  | X |  | X | X |
| *Notes.*  Participants are required to complete device procedure daily from Day 0 to Day 14 and once weekly after Day 14 to Day 82.  Participants will receive introduction to device and overview of how the device works with opportunity to demo and ask questions at screening visit.  * All participants will be offered a safety reporting follow-up and permitted to change antihypertensive medication (if needed) after tAN procedure has ceased/ participants are not using the device. Participants will be encouraged to keep a record of their HR & BPs at home during this period however this is not mandatory. This follow-up is to assess and collate AffeX-CT device safety and efficacy prolonged data.  ** All participants will be asked to not take their morning medications prior to the visits; they will be asked to bring their medication with them and administer their medication(s) in the Investigation Site, after their office BP measurements have been taken. If the participant has taken the medication prior to the visit, site staff will make a note on the medical files and eCRF of the participant. If the participant has not taken their medication to the site and did not receive their medication prior to the visit – and site staff cannot arrange to obtain the medication locally – the visit will be re-scheduled.  *** Baseline Holter and/or Echocardiogram that is conducted within a period of -28 to +3 days from the randomization/baseline visit, will be allowed if there are no changes in the blood pressure treatment between the time of that investigation and randomization  **** Visit 5 Holter and/or Echocardiogram that is conducted within a period of -5 to +14 days of the Visit 5 clinic date  † To be completed for sub-study only.  **X^1^** Any incomplete medical history sections to be reviewed and completed.  **Vital signs^2^** Pulse rate, respiratory rate, temperature and oxygen saturation assessments.  **X^3^** Screening 24-hour ABPM will be used at baseline, only if within screening period and no subsequent treatment changes have been made. Otherwise, 24-hour ABPM must be repeated at baseline (randomisation) visit.  **Office BP^4^** 3 readings will be performed and average mean (after excluding first of 3 BP readings) calculated.  **Blood Test^5^** 1 SST and 2 EDTA; Full blood count (FBC), lipid profile, glucose (fasting), HbA1c, fructosamine, U&Es, Serum pregnancy for female participants on screening and randomisation visits.  Blood samples for storage and later evaluations (plasma and serum): 2 SSTs and 1 EDTA; These will be collected and stored for future analysis, if need (i.e., in case new information arises or if there is a safety signal, and or further investigation is required. The stored samples will be used for the sub-study analysis in a batch basis, and will be used if there are any signals that will indicate that a more mechanistic or pathophysiological insight is required.  **Urine Sample^6^** Urinary albumin creatinine ratio and urinary antihypertensive drug screen.  **X^7^** For subgroup only - ATONT assessment on both arms will completed with participants’ consent at baseline visit and/or within 5 days after baseline & randomisation visit; Investigation site team will endeavour to complete assessment at baseline & randomisation visit.  **X^8^** For subgroup only – ATONT assessment on both arms will completed at end of treatment visit and/or within +/-5 days visit window.  **X^9^** Inclusion and Exclusion criteria will be reviewed and any incomplete sections subsequent to screening visit will be completed.  **X^10^** Device and training will be provided to the participant. Following training participants will be observed using the device during the visit to ensure they are competent with AffeX-CT device handling. Participants will be observed for 30 minutes after self-stimulation within the research facility, and their sitting blood pressure (X3 times) will be recorded before they are allowed to go home.  **X^11^** Participants will be reminded to complete AffeX-CT device procedure and logbook entry.  **X^12^** AffeX-CT device logbook will be reviewed.  **X^13^** Participants will stop tAN Procedure and will return the device. | | | | | | | | | | | |

**Supplementary Table 5.** Secondary endpoints.

**Secondary Endpoints**

•	Change in average daytime ambulatory systolic BP from baseline and 1 month.

•	Change in average daytime ambulatory diastolic BP from baseline to the end of treatment (3 months).

•	Controlled BP at the end of treatment (3 months) defined as mean daytime ambulatory systolic BP<135 mmHg and mean daytime ambulatory DBP< 85 mmHg.

•	Change in average 24-hour ambulatory systolic BP and DBP from baseline to the end of treatment (3 months).

•	Change in average office systolic BP and DBP from baseline to 1 month, and from baseline to the end of treatment (3 months).

•	Change in average daytime ambulatory HR, and in average night-time ambulatory HR from baseline to the end of treatment (3 months).

•	Change in BP variability defined as the coefficient of variation (SD/mean) of 24-hour ambulatory systolic BP, and of within-visit office systolic BP from baseline to the end of treatment (3 months).

•	Change in HR variability defined as the coefficient of variation (SD/mean) of 24-hour ambulatory HR, and of within-visit office HR from baseline to the end of treatment (3 months).

•	Occurrence of a serious adverse event (SAE), fatal or non-fatal, within 3 months.

•	The occurrence of a major cardiovascular event (MACE), including myocardial infarction (MI), stroke, and cardiovascular-related mortality within 3 months.

•	Change in Quality of life between baseline and the end of the treatment (3 months) using the EuroQol Visual Analogue score (0-100), and the EuroQol 5 Dimension (EQ5D) quality of life (QoL) questions.

•	Change in sleep quality between baseline and the end of the treatment (3 months) using the insomnia severity index (ISI), a 7-item questionnaire with each question allowing responses on a 5-point Likert scale from 0-4. Responses to the 7 questions can be summed to give an overall score of 0 to 28.

•	Adherence to trial therapy, assessed as the proportion of days out of total days in follow-up when therapy was self-administered, and the average daily duration of self-administered therapy over the 3 months of follow-up (90 days).

**Supplementary Table 6.** Exploratory endpoints.

**Exploratory endpoints**

•	Change in left ventricular ejection fraction (LVEF), left ventricular mass (LVM), relative mass thickness (RMT), left atrial volume (LAV), left ventricular end-diastolic pressure (LVEDP), and E/e´ ratio from baseline to the end of treatment (3 months), in participants receiving treatment with tAN therapy.

•	Changes in anti-hypertensive medication between baseline and the end of follow-up (4 months) in participants receiving treatment with tAN therapy.

•	Average daily number of antihypertensive medications that participants are on assessed through urinary drug screening (UDS).

•	Change in Quality of life between baseline and the end of follow-up (4 months) in a sub-group of participants receiving treatment with tAN therapy, using the EuroQol Visual Analogue score (0-100), and the EuroQol 5 Dimension (EQ5D) quality of life questions.

•	Change in average central BP, measured by Sphygmocor Vx device, from baseline to 1 month, and from baseline to the end of treatment (3 months) – only for sub study participants.

•	Change in the cognitive functions from baseline to the end of treatment (3 months)

•	Central respiratory modulation of cardiac vagal tone.

•	Overall baroreflex gain (central baroreflex responsiveness).

•	Cardiodepressor and vasodepressor responses of the carotid sinus to digital massage.

2.4.1 Feasibility Endpoint (s)

•	AffeX-CT device-related AEs within 3 months.

•	Adherence to use of the AffeX-CT device assessed using the Extent of Adherence (EoA) questionnaire 23

•	Ease of use of AffeX-CT device assessed using a participant feedback visual analogue scale (VAS).

•	Reasons behind any early withdrawals from the study.

•	Success of the blinding procedure assessed using a blinding index at 1 month and 3 months 24
