## Supplementary material for "A Sham Controlled Randomised Trial evaluating the Safety, Acceptability and Efficacy of Autonomic neuromodulation using Trans-Cutaneous vagal sensory stimulation in uncontrolled Hypertensive patients: Rationale and study design of the SCRATCH-HTN Study": AffeX-CT User Guide

#### Instructions for Use (IFU)

| APPROVAL SIGNATORIES | | | | |
|---|---|---|---|---|
| **Role** | **Name** | **Position** | **Signature** | **Date** |
| Author | Everard Mascarenhas | CEO | | **26.01.2022** |
| Reviewer | Nikolai Gourine | PM | | **26.01.2022** |

### AffeX - CT

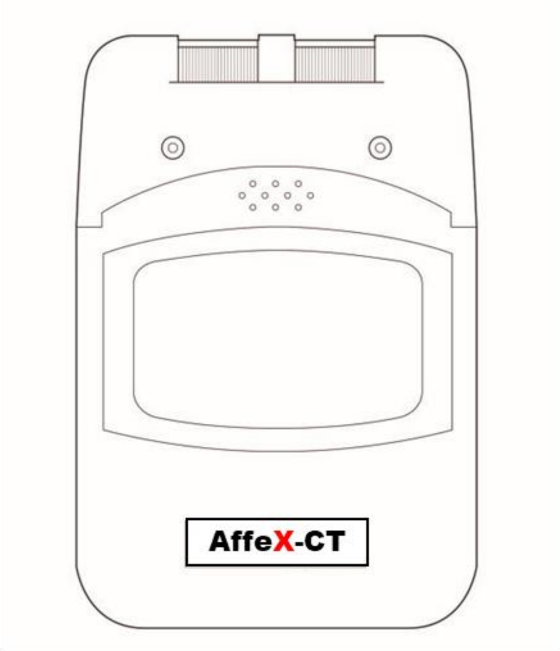


**Operation Manual**
Read Before Using

**Exclusively for clinical investigations**

### TABLE OF CONTENTS


| | |
|---|---|
| What Is Hypertension? | 2 |
| How does AffeX-CT work? | 2 |
| Indications and Contraindications | 3 |
| Warnings and Precautions | 4 |
| Precautions/Adverse Reactions | 5 |
| About This Device | 6 |
| Unit Controls | 7 |
| Attaching The Lead Wires | 7 |
| Ear clips use and Care | 8 |
| Tips For Skin Care | 8 |
| Connecting the AffeX-CT device | 9 |
| Caring For Your AffeX-CT Device | 10 |
| Troubleshooting | 11 |
| System Components | 11 |
| Technical Specifications | 12 |
| Output Parameters | 12 |
| Description of symbols | 13 |
| Warranty | 14 |

### INTRODUCTION TO AFFEX-CT

#### What is Hypertension?

Hypertension, also known as high or raised blood pressure, is a condition in which the blood vessels have persistently raised pressure. Blood is carried from the heart to all parts of the body in the vessels. Each time the heart beats, it pumps blood into the vessels. Blood pressure is created by the force of blood pushing against the walls of blood vessels (arteries) as it is pumped by the heart. The higher the pressure, the harder the heart has to pump.

Hypertension is a serious medical condition and, if left untreated, can increase the risk of heart and circulatory system diseases like heart attack and stroke, as well as lead to kidney failure, heart failure, vision problems, vascular dementia and a range of other diseases.

#### How does AffeX-CT work?

Cardiovascular disease is one of the leading causes of death and illness, therefore, managing high blood pressure being a key focus of treatment. AffeX-CT is a small battery-operated medical device, which has been designed to use transcutaneous autonomic neuromodulation to externally stimulate the nerve endings that control blood pressure, via the tragi of the ear, to treat patients with uncontrolled blood pressure or drug-resistant hypertension.

.

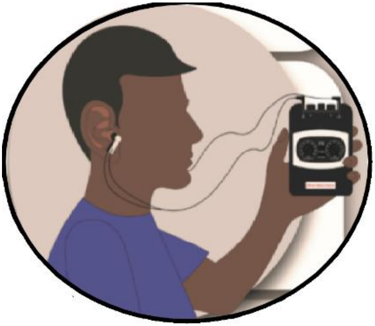

### INDICATIONS AND CONTRAINDICATIONS

Read the operation manual before using AffeX-CT

#### INDICATIONS

AffeX-CT is to be used exclusively for clinical investigations.

#### CONTRAINDICATIONS

- Patients with implanted electronic devices (for example, a pacemaker) or electronic metallic implants should not undertake AffeX-CT treatment
- Patients with all other non-electronic implants should only use this device after discussion with a physician to ensure that the implant and stimulation do not interact.
- The use of Affex-CT device is contra-indicated if a person is suffering from chronic headaches, neck pains or tooth pain symptoms which have no clear diagnosis. These patients can only use this device on recommendation from their consultant, and after the cause of the symptoms is known.

- Patients with history of hospitalization for heart failure, or cerebrovascular accidents, or stroke should not use this device, unless specifically recommended by their specialist / consultant aware of this history.

- Patient with epilepsy or taking antiepileptic agents. Those with history of seizure more than 10 years ago and currently not on medications can use this device only after recommendation from and under supervision of their consultant.

- Patients with history of stable or unstable angina in the last one year. Those with history of these symptoms beyond one year can only use this device on recommendation from and under supervision of their consultant / cardiologist.

- Patients with history of hospitalization for hypertensive emergency or urgency, should not use this device, unless specifically recommended by their specialist/consultant aware of this history.

- Patients suffering from chronic pain where anti-inflammatory drugs have been taken for two or more days per week over the last month should not use this device.

- Patients who are pregnant, nursing or planning to become pregnant within the next 6 months should not use this device.

- Patients with piercing in tragus/ tragi should not use this device.

### WARNINGS AND PRECAUTION

##### WARNINGS

- AffeX-CT devices must be kept out of reach of children.

- If AffeX-CT treatment becomes difficult to tolerate even after reduction in the stimulation intensity (or on minimum levels) then please discontinue using this device until evaluation by a physician.

- Always turn the AffeX-CT device OFF before attaching or removing the ear-clips.

- Electrodes should not be placed over the eyes, in the mouth, or internally.

- The patient is an intended operator. If someone else is helping the patient to use the device, a higher level of stimulation intensity may be applied and can potentially cause pain and discomfort.

**PRECAUTIONS/ADVERSE REACTIONS**

- Isolated cases of skin irritations may occur at the site of the ear-clip placement.

- Skin irritation and redness are potential adverse reactions. These may become evident if the stimulation exceeds the prescribed level.

- Patients must not alter device settings at any time.

- Patients must not stimulate at higher settings then prescribed as this may result in discomfort or mild pain at the site of stimulation.

- Patients may experience discomfort or pain at the level of stimulation which they have tolerated before. In that case, they should reduce the level of stimulation one intensity dial division by one until they are at a comfortable level of stimulation.

- The applied part is an ear-clip electrode.

- Device must not be disposed of and must be returned to the issuing hospital or clinic.

- No modification of this equipment is allowed.

- Headphones, AirPods or Bluetooth ear devices should not be used when using the AffeX-CT device

#### ABOUT THIS DEVICE

Your AffeX-CT is a battery-operated device that includes two controllable output channels. The Affex-CT device generates electrical stimulation pulses whose intensity (amplitude) can be altered with the controls or switches, however, the duration and other parameters of stimulation are fixed (pre-set).

The Indicator Lights (located below the intensity knobs) indicate that the stimulation is being applied and also that there is sufficient battery power to operate the AffeX-CT device.

Users must consult a physician/clinician before using the AffeX-CT device.

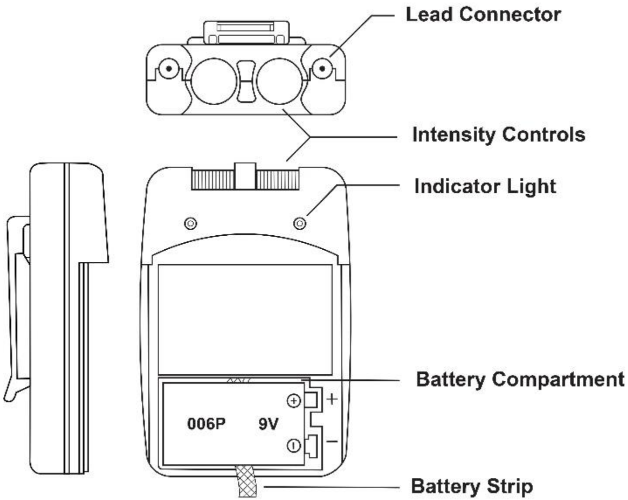

### UNIT CONTROLS

#### Intensity

The intensity knobs located on the top of the device unit, control the strength of the stimulation and also function as ON/OFF switches.

#### Resetting the Timer

To resume operation or to reset the timer, simply turn the intensity control OFF and then ON again.

### ATTACHING THE LEAD WIRES

Insert the lead wires provided with the device into the jack sockets located on the top panel of the unit. Holding the insulated portion of the connector, push the plug end of the wire into one of the jacks; two pairs of the electrodes should be used. (NB: ear-clips attached to the leads are colour coded to help the participants identify which one to use for the right and left ear: red coloured ear clip is for the left ear, and the white coloured ear clip is to be attached to the right ear).

**NOTE: Use care when plugging and unplugging the wires. Pulling on the lead wire instead of its insulated connector may cause wire or connector damage.**

**CAUTION: Never insert the plug of the lead wire into an AC power supply socket.**

#### EAR CLIPS USE AND CARE

Follow the ear-clip application procedures outlined in the Instructions For Use

It is recommended that the ear-clips are applied after moistening the skin (see details on tips for skin care).

Ear-clips are applied to both tragi

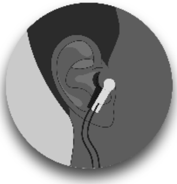

For detailed use of the device, please read the instructions below and also you may find useful information in User Guide given separately.

After stimulation, please switch OFF the device and remove the ear-clips. Moisten the area again. Clean the ear-clips and take out the leads from the device and store them in the box.

#### TIPS FOR SKIN CARE

Good skin care is important for comfortable use of the AffeX-CT device.

- Always clean and moisturise the ear-clip pads with tissue dipped in water
- Always wipe the skin clean on both side of the tragus with a wet wipe
- Any excess hair should be removed to ensure good ear-clip contact with the skin.
- You may choose to use a skin treatment or preparation that is recommended by your physician/clinician. Apply, let it dry, and attach ear-clips as directed. This will both reduce the chance of skin irritation and extend the life of your ear-clip electrodes.

### CONNECTING THE AFFEX-CT DEVICE

<table>
<tr><td>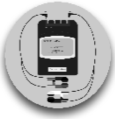</td><td>Insert red ear-clip lead into the left CH1 and white ear-clip lead into the right CH2 channel receptacles, gently pushing the plug as far as it will go.</td></tr>
<tr><td>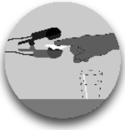</td><td>Moisturise the part of the clip in contact with your ear, by wiping it with a wet tissue (e.g., dipping in water)</td></tr>
<tr><td>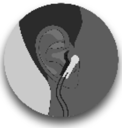</td><td>Attach the ear-clips to the tragi on both sides, ensuring the red part of the first clip is attached to the outer side of the left tragus and the white part of the second clip is attached to the outer side of the right tragus.</td></tr>
<tr><td>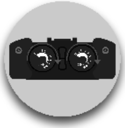</td><td>The device is turned ON by turning both the left and right dials to the right (clockwise).<br>A green light will be ON when the stimulation is applied</td></tr>
<tr><td colspan="2">Locate the rotary intensity control knob at the top of the unit.  Turn knobs 1 and 2 clockwise.  Check the indicator light is ON, this will light up as long as the unit is in operation. Slowly turn the channel control clockwise until you reach the prescribed intensity. Repeat for both channels.</td></tr>
</table>

| | |
|---|---|
| 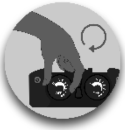 | For each ear:<br>Turn the device on by turning the dial slowly towards the right (clockwise) until you reach the correct stimulation level prescribed at your training session. You may or may not feel a tingling sensation. |
| 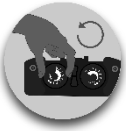 | If you feel a tingling sensation and it is uncomfortable, turn the dial down one number lower. You may need to dial it one number further down during the treatment session if the sensation is uncomfortable. If after going down a level, the tingling sensation still persist, please go further down the level and repeat this until the stimulation is tolerable. |
| 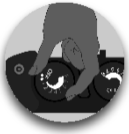 | Do NOT turn the dial above the stimulation level prescribed to you for each ear. Please note that when you are using your device at the prescribed stimulation level settings, you may or may not experience the sensation you felt during the assessment at the time of setting these levels. However, if treatment at this level produces clear discomfort you are advised to reduce the stimulation level by one unit. |
| After 30 minutes the stimulation will stop automatically | |
| Turn both control knobs anticlockwise to the OFF position. | |

Please refer to the User Guide for further guidance and information on using AffeX-CT. There is also a video with advice and a demonstration on using AffeX-CT. Please see the following link [Video link here]

#### CARING FOR YOUR AffeX-CT DEVICE

Your AffeX-CT device may be cleaned by wiping gently with a damp cloth moistened with mild soap and water. Never immerse the device in water or other liquids.

- Handle the device with care.
- Wipe the lead wires with a damp cloth as above if they become soiled.

- Store AffeX-CT device in a cool and dry place.

#### TROUBLESHOOTING

If the AffeX-CT device does not function properly:

1. If the green indicator light on the front of the unit does not stay lit when the unit is turned on, turn both intensity control knobs to the OFF position (anticlockwise), wait for ~1 min, and then turn the intensity control knobs ON. If the unit is not functioning after turning it OFF and ON, contact the device team to replace the device;
2. If the electrode leads are broken (e.g., the ear-clip or connector plugs are detached from the lead) use the spare electrode lead provided or contact the device team to replace the damaged leads.

If there is any other problem with the device, please contact the device team or study supervisor. Do not try to repair your device.

#### SYSTEM COMPONENTS

Your AffeX-CT device set includes the following components and accessories:

- AffeX-CT unit
- Four electrode ear-clips/connection leads
- 6F22 Battery (Installed)
- Information for Users (Operating Manual)
- User Guide
- Case/box

### TECHNICAL SPECIFICATIONS

| | |
|---|---|
| Channel: | Dual, isolated between channels |
| Modes of Operations: | Continuous |
| Pulse Intensity: | Adjustable 0-80mA<br>Constant current |
| Pulse Rate: | 30 Hz (fixed) |
| Pulse Width: | 200 µsec (fixed) |
| Timer: | Continuous, 30 min (fixed) |
| Wave Form: | Asymmetrical Bi-Phasic square pulse |
| Power Source: | 9 volt battery (Type 6F22) |
| Dimensions: | 95(H) x 60(W) x 23 (T) mm |
| Weight: | 115 grams (battery included) |
| Operating temperature | 5$^{o}$C - 30$^{o}$C |

#### Output Parameters

| Mode | Intensity (mA) | Width (µsec) | Pulse Rate Freq(Hz) | Cycle Time (Sec) |
|---|---|---|---|---|
| Continuous | Adj. 0-80 | Fixed 200 | Fixed 30 Hz | N/A |

Prescription Intensity Range

| | | |
|---|---|---|
| Prescribed Intensity Range | 0.1-8mA | Users will be provided with an individual prescription level of stimulation and instructed not to exceed this level |

#### Description of Symbols:

There are a number of technical symbols on your unit explained as follows:

| | | | |
|---|---|---|---|
| | **WARNING:** Failure to follow instructions may result in serious injury or death to the patient or user | | **PRECAUTION:** Failure to follow instructions may result in damage to the equipment or degradation in the quality of treatment |
| | Manufacturer | **IP22** | Protection from solid foreign objects ≥12.5 mm and ingress of water at 15° |
| REF | Reference Number / Catalogue Number | | Type BF applied parts |
| SN | Serial Number | | Refer to instructions |
| | Non-sterile | ***Exclusively for clinical investigations*** | AffeX - CT is only to be used in regulatory-approved clinical studies |
| | Separate collection for waste of electrical and electronic equipment | | |

### Warranty

This AffeX-CT device carries a one-year warranty from the date of delivery.

The warranty does not apply to damage resulting from failure to follow the operating instructions, accidents, abuse, alterations or disassembly by unauthorized individuals.

The warranty applies to the main device and necessary parts and labour relating thereto. Battery, ear-clip electrodes, and other accessories are warranted to be free from defects in workmanship and materials at the time of delivery.

The Manufacturer reserves the right to replace or repair the unit at its discretion.

**Manufactured and distributed by:**

Afferent Medical Solutions Ltd.
6 Almond Avenue, Ickenham
UB10 8NA
United Kingdom
afferentmedicalsolutions.com


Afferent Medical Solutions Ltd
6 Almond Avenue, Ickenham
UB10 8NA
