## Supplementary material for "A Sham Controlled Randomised Trial evaluating the Safety, Acceptability and Efficacy of Autonomic neuromodulation using Trans-Cutaneous vagal sensory stimulation in uncontrolled Hypertensive patients: Rationale and study design of the SCRATCH-HTN Study": Statistical Analysis Plan

**SCRATCH–HTN**

***Sham controlled Randomized Control Trial evaluating the Safety, Acceptability and Efficacy of Autonomic neuromodulation using trans-cutaneous vagal sensory stimulation in uncontrolled hypertensive patients: a pilot study evaluating a novel non-invasive device-based strategy.***

### *Statistical Analysis Plan*

***Version 2.0***

Version 1.0 started: 11/10/2023.

Version 2.0 completed: 22/04/2025

Author of Version 1.0: Kamran KHAN

Author of Version 2.0: Annastazia LEAROYD

### Signatures

Chief Investigator

**Signature:** 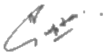

Blinded Study Statistician

**Signature:** 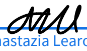
Annastazia Learoyd (May 20, 2025 15:31 GMT+1)

### Other Contributors/Reviewers

**Hui Zhen Tam**

Independent Statistician within Barts CTU

**Jane Field**

Trial Co-ordinator

### Version History

| SAP Version | CIP Version | Date | Reason for revision | Summary of changes made |
|---|---|---|---|---|
| 1.0 | V9.0 15JAN24 | | First draft | - |
| 2.0 | V10.0 29OCT24 | 22/04/25 | Substantial amendment of CIP to v10.0 | Change reference to protocol to reference to CIP matching correct nomenclature.<br>Changes to the inclusion and exclusion criteria to match the updated CIP.<br>Clarification of the term baseline ABPM in section 4.1 to match the updated CIP.<br>Removal of the requirement to use two sample t-tests with equal variances in the first instance to match the updated CIP.<br>Addition of analysis population removing participants with >120% days of device usage (overcompliance), as well as subgroups based on days of device usage.<br>Addition of the description of covariates indicating compliance being included in sensitivity analyses.<br>Addition of possible additional statistical methods that may be employed to complement the main analysis.<br>Addition of exploratory endpoints and analysis for these endpoints, matching the CIP.<br>Addition of exploratory analysis of collected study data not defined as a study endpoint in the CIP. |

#### Abbreviations and Definitions

| | |
|---|---|
| 6MWT | Six Minute Walk Test |
| ABPM | Ambulatory Blood Pressure Monitoring |
| ABS | Acrylonitrile butadiene styrene |
| ACEI | Angiotensin-Converting Enzyme Inhibitors |
| ADE | Adverse Device Effect |
| AE | Adverse Event |
| AFT | Autonomic Function Test |
| APR | Annual Progress Report |
| AR | Adverse Reaction |
| ARBs | Angiotensin II receptor blockers |
| ATONT | Autonomic Target-organs Neurophysiological tests |
| BMI | Body Mass Index |
| BP | Blood Pressure |
| BPM | Beats Per Minute |
| BBR | Baroreflex Responsiveness |
| CAPA | Corrective and Preventive Action |
| CCBs | Calcium-Channel Blockers |
| CI | Chief Investigator |
| CIP | Clinical Investigational Plan |
| CKD | Chronic Kidney Disease |

| | |
|---|---|
| CRC | Clinical Research Centre |
| CSB | Cardiac Sensitivity to Baroreflex |
| CVCTU | Cardiovascular Clinical Trials Unit |
| CVD | Cardiovascular Disease |
| CVT | Cardiac Vagal Tone |
| DBP | Diastolic Blood Pressure |
| DD | Device Deficiency |
| DSUR | Development Safety Update Report |
| ECG | Electrocardiogram |
| eCRF | Electronic Case Report form |
| eGFR | Estimated Glomerular Filtration Rate |
| EoA | Extent of Adherence |
| EQ5D | EuroQol 5 Dimension |
| EUDAMED | European Medical Devices Regulatory Database |
| FBC | Full Blood Count |
| GCP | Good Clinical Practice |
| GP | General Practitioner |
| HR | Heart Rate |
| HRA | Health Research Authority |
| IB | Investigator Brochure |
| ICH | International Conference on Harmonisation |
| ISI | Insomnia Severity Index |
| DSMC | Data Safety Monitoring Committee |
| IMD | Investigational Medical Device |
| ISF | Investigator Site File |
| JRMO | Joint Research Management Office |

| | |
|---|---|
| LAV | Left Atrial Volume |
| LPLV | Last Patient Last Visit |
| LVEDP | Left Ventricular End-Diastolic Pressure |
| LVEF | Left Ventricular Ejection Fraction |
| LVM | Left Ventricular Mass |
| LVS | Linear Vagal Scale |
| MACE | Major Adverse Cardiovascular Events |
| MAP | Mean Supine Arterial Blood Pressure |
| MDR | Medical Device Regulations |
| MHRA | Medicine and Healthcare Regulatory Authority |
| mm Hg | Millimetre of Mercury |
| mmo\L | Millimoles per litre |
| MRA | Mineralocorticoid receptor antagonists |
| NHS | National Health Service |
| NICE | The National Institute for Health and Care Excellence |
| NSAIDs | Non-Steroid-Anti-inflammatory Drugs |
| OTC | Over-The-Counter |
| PIC | Patient Identification Center |
| PIS | Participant Information sheet |
| PoC | Proof of Concept |
| PPI | Patient and Public Involvement |
| QA | Quality Assurance |
| QC | Quality Control |
| QoL | Quality of Life |
| RSG | Research Steering Committee |
| RSI | Reference Safety Information |

| | |
|---|---|
| RMT | Relativeness Mass Thickness |
| SADE | Serious Adverse Device Effect |
| SAE | Serious Adverse Event |
| SAR | Serious Adverse Reaction |
| SBP | Systolic Blood Pressure |
| SD | Standard deviation |
| SDA | Source Data Agreement |
| SOP | Standard Operating Procedure |
| tAN | Trans-cutaneous Autonomic Neurostimulation |
| TMF | Trial Master File |
| TMG | Trial Management Group |
| TSC | Trial Steering Committee |
| UDS | Urinary Drug Screen |
| UK | United Kingdom |
| USADE | Unanticipated Serious Adverse Device Effect |
| VAS | Visual Analogue Scale |

### 1 Summary of trial

| Trial Information | |
|---|---|
| Chief Investigator | **Dr Ajay Gupta**<br>Clinical Reader and Consultant<br>Barts Cardiovascular Clinical Trials Unit (CVCTU)<br>William Harvey Research Institute – Heart Centre<br>Barts and The London School of Medicine and Dentistry, Queen Mary University of London, Charterhouse Square<br>London EC1M 6BQ<br><br>Phone number: 020 7882 2858 |
| Sponsor Name | **Queen Mary University of London**<br>Contact person:<br>Mays Jawad<br>Research Governance and Operations Manager<br>Joint Research Management Office (JRMO)<br>Barts Health NHS Trust & Queen Mary University of London<br>Research Services \| Dept. W \| 69-89 Mile End Rd \| London \| E1 4UJ T: (020) 7882 7207 \| \| W: www.jrmo.org.uk<br>Department inbox: |
| Trial identifiers: | IRAS: 302061<br>ISRCTN: 14509154<br>NCT05179343 |
| REC Number | 21/WS/0157 |

| | |
|---|---|
| Trial Design | Double-blinded sham-controlled parallel group trial with a block randomization on a 2:1 basis |
| Trial Phase | II |
| Study Design | Single-site pilot/Phase II |
| Study Objectives | ***Primary:*** To determine whether treatment with tAN therapy can reduce daytime SBP in uncontrolled hypertensive patients compared to the placebo sham therapy.<br><br>***Secondary:***<br>• To determine whether treatment with tAN therapy can reduce daytime DBP to a greater extent than treatment with a sham therapy.<br>• To determine whether treatment with tAN therapy can reduce 24-hour SBP and DBP to a greater extent than treatment with a sham therapy.<br>• To determine whether treatment with tAN therapy can lead to a higher proportion of patients with controlled BP than treatment with a placebo sham therapy.<br>• To evaluate differences in BP variability between those receiving tAN therapy compared to those receiving sham therapy.<br>• To evaluate differences in HR variability between those receiving tAN therapy compared to those receiving sham therapy.<br>• To evaluate differences in reported serious adverse events (SAEs) and major adverse cardiovascular events (MACE) between those receiving tAN therapy compared to those receiving sham therapy.<br>• To determine whether treatment with tAN therapy can improve quality of life and well-being to a greater extent than treatment with a placebo sham therapy. |

| | |
|---|---|
| | • To determine whether treatment with tAN therapy can improve quality of sleep to a greater extent than treatment with a placebo sham therapy.<br>• To evaluate differences in cumulative adherence to medications between |
| Target Sample Size | 63 patients with a 2:1 ratio: 42 in the active and 21 in the placebo sham controlled. |
| Randomisation Method | Dynamic minimisation approach using Sealed Envelope.<br>Balancing factors are: age at baseline (categories: <65; 65+ years), sex, BMI at baseline (categories: <30; 30+ kg/m2), and baseline mean daytime SBP (categories: <160 mmHg; 160+ mmHg) |
| PICOT Summary | |
| **P**opulation (medical condition or disease under investigation) | Systemic hypertension |
| **I**ntervention (description, frequency, details of delivery) | Using a non-invasive Transcutaneous Autonomic Neuromodulation (tAN) device (Affex-CT) for 12 weeks with a 30 min of tAN once per day for the first two weeks and once every week for the rest of the trial period. |
| **C**omparator intervention (description, frequency, details of delivery) | The comparator will be a non-active sham tAN device |
| **O**utcomes (including key endpoints) | ***Primary:***<br>• Change in average daytime ambulatory SBP from baseline to the end of treatment (3 months)<br>***Secondary:***<br>• Change in average daytime SBP and DBP from baseline and 1 month. |

| | |
|---|---|
| | • Change in average daytime ambulatory DBP from baseline to the end of treatment (3 months).<br>• Controlled BP at the end of treatment (3 months) defined as mean daytime ambulatory SBP < 135 mmHg and mean daytime ambulatory DBP < 85 mmHg.<br>• Change in average 24-hour ambulatory SBP and DBP from baseline to the end of treatment (3 months).<br>• Change in average office SBP and DBP from baseline to 1 month, and from baseline to the end of treatment (3 months).<br>• Change in average daytime ambulatory HR, and in average night-time ambulatory HR from baseline to the end of treatment (3 months).<br>• Change in BP variability defined as the coefficient of variation (SD/mean) of 24-hour ambulatory SBP, and of within-visit office SBP from baseline to the end of treatment (3 months).<br>• Change in HR variability defined as the coefficient of variation (SD/mean) of 24-hour ambulatory HR, and of within-visit office HR from baseline to the end of treatment (3 months).<br>• Occurrence of a serious adverse event (SAE), fatal or non-fatal, within 3 months.<br>• The occurrence of a major cardiovascular event (MACE), including myocardial infarction (MI), stroke, and cardiovascular-related mortality within 3 months.<br>• Change in Quality of life between baseline and the end of the treatment (3 months) using the EuroQol Visual Analogue score (0-100), and the EuroQol 5 Dimension (EQ5D) quality of life (QoL) questions.<br>• Change in sleep quality between baseline and the end of the treatment (3 months) using the insomnia severity index (ISI), a 7-item questionnaire with each question allowing responses |

| | |
|---|---|
| | on a 5-point Likert scale from 0-4. Responses to the 7 questions can be summed to give an overall score of 0 to 28.<br>• Adherence to trial therapy, assessed as the proportion of days out of total days in follow-up when therapy was self-administered, and the average daily duration of self-administered therapy over the 3 months of follow-up (90 days). |
| Treatment duration per participant (minimum and maximum duration planned) | The total treatment duration is 12 weeks.<br>Self-administration of 30 min of tAN or sham stimulations once per day for the first two weeks, and then once every week for the rest of the trial period. |
| Length of Patient Follow-up | Telephone follow-up at 16 weeks (4 weeks after the end of treatment/trial period of 12 weeks). |
| Eligibility Criteria | |
| Inclusion Criteria | 1) Participant has given written informed consent.<br>2) Participant has sufficient knowledge of the English language to be able to understand the participant information sheet and trial materials including outcome assessments.<br>3) Participant is aged ≥18 years and <80 years at the time of screening visit.<br>4) Participant is taking between 1 to 4 antihypertensive medications (inclusive) at time of screening and baseline (randomisation) visit and is willing to adhere to no change in medication during the trial until end of the trial visit (visit 5). (NB. Participant on only one antihypertensive medication should be taking that medication for at least six weeks prior to the screening visit).<br>5) Participant has confirmed diagnosis of hypertension.<br>6) Participant meets the following BP criteria:<br>• 24-hour ambulatory BP monitoring (ABPM) at either screening visit or baseline (randomisation) visit, with mean daytime SBP of ≥135 mmHg and <170 mmHg and mean daytime DBP of ≥85 mm Hg and <115 mmHg (N.B. By default, Ambulatory Blood Pressure Monitoring [ABPM] at |

| | |
|---|---|
| | screening visit will be used at baseline visit. However, if there has been an addition of new medication after participants screening visit, 24-hour ABPM must be repeated at baseline visit, but the screening ABPM will be used for eligibility criteria).<br>7) Participant has one or more of the following associated conditions:<br>a) Obesity: BMI >30 OR waist circumference >94 cm (men) or > 80cm (women). (NB. For participants of South-East Asian/Chinese/Japanese origin these cut-offs are >90 cm (men) or >80 cm (women)).<br>b) Type 2 diabetes – controlled or sub-optimally controlled (HbA1c ≤8.5% or ≤69 mmol/mol) on diet and/ or medications except insulin.<br>c) Heart rate (on any one of the three heart rate recordings at that visit) ≥70 bpm at screening or baseline (randomisation) visit (measurements taken after 5 minutes of rest in a seated position and when finger probe has been placed for a minimum of 30 seconds thereafter) or a heart rate (on any one of the three heart rate recordings) ≥60 bpm at screening or baseline (randomisation) visit if the patient is taking beta-blocker medication or a rate-limiting calcium channel-blocker medication.<br>d) HbA1c ≥42 mmol/mol or fasting blood glucose (if available) ≥5.6 mmo/L, and either low HDL cholesterol (≤1.03 mmol/L for men and ≤1.29 mmol/L for women) or high triglyceride (triglycerides ≥1.7 mmol/L)<br>e) Both low HDL cholesterol (≤1.03 mmol/L for men and ≤1.29 mmol/L for women) and high triglyceride (triglycerides ≥1.7 mmol/L)<br>f) Diagnosed or known case of polycystic ovarian syndrome.<br>8) Female participant of child-bearing potential (all those below 55 years except if they are surgically sterile, meaning they have undergone a hysterectomy, bilateral tubal ligation, or bilateral oophorectomy, or formally diagnosed by their doctors to be |

| | |
|---|---|
| | post-menopausal) must agree to use the acceptable methods of contraception from the time of consent until last follow up visit.<br>9) Participant is able to communicate satisfactorily with the Investigator and Investigation Site staff, and to participate in, and comply with all clinical study requirements.<br>10) Participant agrees to have all trial procedures performed and are able and willing to comply with all trial visits and protocol requirements. |
| Exclusion Criteria | If any of the following exclusion criteria is met, the participant cannot be randomised:<br>1) Participant is unable and unwilling to use AffeX-CT device daily.<br>2) Participant has a small tragus (ie. the size or shape of the tragus is such that it doesn't allow the application of the ear-clips of the AffeX-CT device for a sustained period of time).<br>3) Participant has a piercing on the tragus of the ear.<br>4) Participant is diagnosed with atrial fibrillation or other form of cardiac arrhythmia.<br>5) Participant has eGFR <45 ml/min/1.73 $m^2$ at screening visit.<br>6) Participant has type 1 diabetes mellitus.<br>7) Participant has type 2 diabetes mellitus on Insulin or those on oral antidiabetic medications with poor glycaemic control defined as HbA1c above 8.5% (or >69 mmol/mol).<br>8) Participant has a history of falls or symptoms of orthostatic hypotension in the last 3 months prior to baseline (randomisation) visit.<br>9) Participant is pregnant, nursing or planning to become pregnant within the next 6 months.<br>10) Participant suffers from chronic pain and has taken anti-inflammatory drugs for two or more days per week over the last month prior to baseline (randomisation) visit.<br>11) Participant has clinically significant or symptomatic hypertension-mediated target organ damage such as severe heart |

| | |
|---|---|
| | failure with NYHA 4, end stage renal damage, medically diagnosed/imaging proven stroke, symptomatic peripheral vascular disease, or severe retinopathy.<br>12) Participant has a history of stable or unstable angina or had an acute coronary event within 3 months prior to baseline (randomisation) visit or had a myocardial infarction within the last six months of enrolment prior to baseline (randomisation) visit.<br>13) Participant has a history of renal denervation within last 1 year prior to baseline (randomisation) visit.<br>14) Participant has a therapeutic implantable electronic/ electrical device such as pacemaker, implantable cardioverter-defibrillators (ICDs), implanted vagal stimulators.<br>15) Participant has history of hospitalization (>24 hours) for heart failure, or cerebrovascular accidents, or history of stroke diagnosed based on imaging or evidence of specialist diagnosis or any other indirect evidence such as discharge summary or clinical letter (at any time in the past).<br>16) Participant has mean daytime ABPM pulse pressure ≥ 80 mmHg at screening or baseline (randomisation) visit.<br>17) Participant has a heart rate <50 bpm at screening or baseline (randomisation) visit (measurement taken after 5 minutes of rest in a seated position and when finger probe has been placed for a minimum of 30 seconds thereafter).<br>18) Participant has auricular dermatitis.<br>19) Participant has postural hypotension, defined as a fall > 20mmHg in SBP on standing at 3 minutes (compared with sitting).<br>20) Participant has a history of hospitalization for hypertensive emergency or urgency in the last six months of enrolment prior to baseline (randomisation) visit. Hospitalisation' is defined as admission for more than 24 hours or between 12-24 hours with an overnight stay. |

| | |
|---|---|
| | 21) Participant is identified as unsuitable to participate by the CI/Sub-Investigator(s) and/or Investigation site team for another reason (e.g., for other medical reasons, laboratory abnormalities, limited life expectancy, etc.)<br>22) Participants with history of epilepsy and are currently on anti-epileptic medication or those who are not on any anti-epileptic medication but have history of a seizure within last 10 years. |

### 2 Additional study methods

#### 2.1 Sample size calculation

The trial's primary endpoint is the change in daytime ambulatory SBP between baseline and the end of treatment at 3 months in the active treatment arm. As this is a feasibility study the primary aim is to estimate the mean reduction in daytime SBP in the intervention group only, not to test for differences between arms. Based on existing data for hypertension treatment (Figure 1 in CIP, section 1.1, page 23), there is a conservative assumption of a mean change in ambulatory SBP of -5.5 mmHg with a standard deviation of 11mmHg. Using a paired t-test examining mean change in SBP within patients, 34 participants would give 80% power to detect such a change at the two-sided alpha level of 0.05. We assume a 10% drop-out and a further 10% non-compliance level and inflate the sample size to 42 subjects in the intervention arm.

Participants will be recruited to the comparator or sham treatment arm to evaluate the feasibility of a randomised control sham group. For this, we decided to recruit double the number of participants with a 2:1 randomization ratio to the active arm compared to the sham treatment arm to collect more safety data. Therefore, the sample size in the sham arm will be 21 while in the active arm it will be 42 and the total number of participants will be 63 across both the arms.

#### 2.2 Randomisation procedures

Eligible participants will be randomised at the baseline visit to either the active (tAN) or sham (sham-tAN) device arm of the trial at a ratio 2:1 ratio. Double the number of participants will be randomised to active therapy in order to collect more data, particularly safety data for the active intervention.

The dynamic minimisation approach will be used for randomisation with balancing factors of age at baseline (categories: <65; 65+ years), sex, BMI at baseline (categories: <30; 30+ kg/m$^2$), and baseline mean daytime SBP (categories: <160 mmHg; 160+ mmHg) to help achieve a balance in these prognostic risk factors between the randomised trial groups.

Sealed Envelope, an online EDC randomisation and unblinding tool will be used and recorded as part of the eCRF for this trial. The randomisation tool will be available to CI, Co-Investigator (s), Investigation site, Sponsor and CVCTU Coordinating team. A master randomisation/allocation list will also be held by Afferent Medical Solutions Ltd. The Investigation site team will complete a corresponding enrolment log to track recruitment.

#### 2.3 Blinding/unblinding procedures

The CI is responsible for the medical care of each individual trial participant (Declaration of Helsinki section 3, and GCP section 4.3) and the coding system in blinded studies should include a mechanism that permits rapid un-blinding (ICH GCP 5.13.4).

As per Sponsor requirements, a 24-hour emergency unblinding procedure will be available in this trial. Participants will also be given participant trial card with emergency CI, and Investigation site contact details. The CI and Sub-investigators are responsible for unblinding on the trial, and unblinding of a participant allocation will be via Sealed Envelope.

If the person requiring the unblinding is not associated with the CI/Co-Investigator (s) and/or the Investigation site, the requesting health care professional will notify the Investigation site team that an unblinding is required for a trial participant and an assessment to unblind the participant will be made in consultation with the Investigation site and CI. The CI will be notified in writing as soon as possible of the necessity of the code break. Once the participant allocation is revealed, and the requester is notified, the CI/Co-Investigator (s) or Investigation site team will detail the reason for the unblinding on the eCRF, participant trial file and electronic participant records/clinical notes.

The sponsor's and manufacturer's safety reporting teams will be unblind to the trial intervention. In the event that a safety event must be reported to the MHRA unblind, a member of these teams will check the participant's allocation via Sealed Envelope and submit an unblind report, without the need to unblind the study team.

All trial unblinding will be disseminated to the Data and Safety Monitoring Committee for review in accordance with the charter. Participant unblinding will be documented in the End of Trial and statistical reports.

#### 2.4 Patient follow-up

##### 2.4.1 Definition of treatment adherence

Participants will be asked to self-administer tAN therapy using AffeX-CT device for 14 days (after Day 0) each day for the duration of 30 minutes in the late evening, preferably between 18.00-22.00. After 14 days, the participants will be asked to self-administer tAN therapy once a week for 10 weeks- i.e. until end of treatment period of 12 weeks. The device logbooks will be examined and the compliance with the device use will be assessed during the 3 visits (visit 3, 4 and 5) by the study team.

Per the CIP, we will use 80% or more days of device usage out of the maximum possible to define as good compliance, and less than 33% days of device usage as poor compliance. A per-protocol analysis will be completed using data from those who have good device usage (i.e. 80+% days of device usage or ≥19 days of usage out of the maximum of 24). Additional analyses considering excluding those with more than 120% days of device usage will also be considered.

Treatment adherence is considered a secondary outcome for this study. As a result, endpoints related to adherence will be reported as described in the Main analysis (Section 5.2) below.

##### 2.4.2 Frequency and duration of follow-up

Participants will be required to attend five visits, the sequence is as follows: visit 1 (screening), visit 2 (baseline and randomisation - Day 0), visit 3 at Day 14, visit 4 at Day 28 and visit 5 (End of Treatment - Day 84). Participants will also receive telephone conversations; one phone call between days 1-3, one at Day 7, and one at Day 56 and Day 112 (post trial follow up). Participants will also receive a text and/or email reminders at Day 42 and Day 70, respectively.

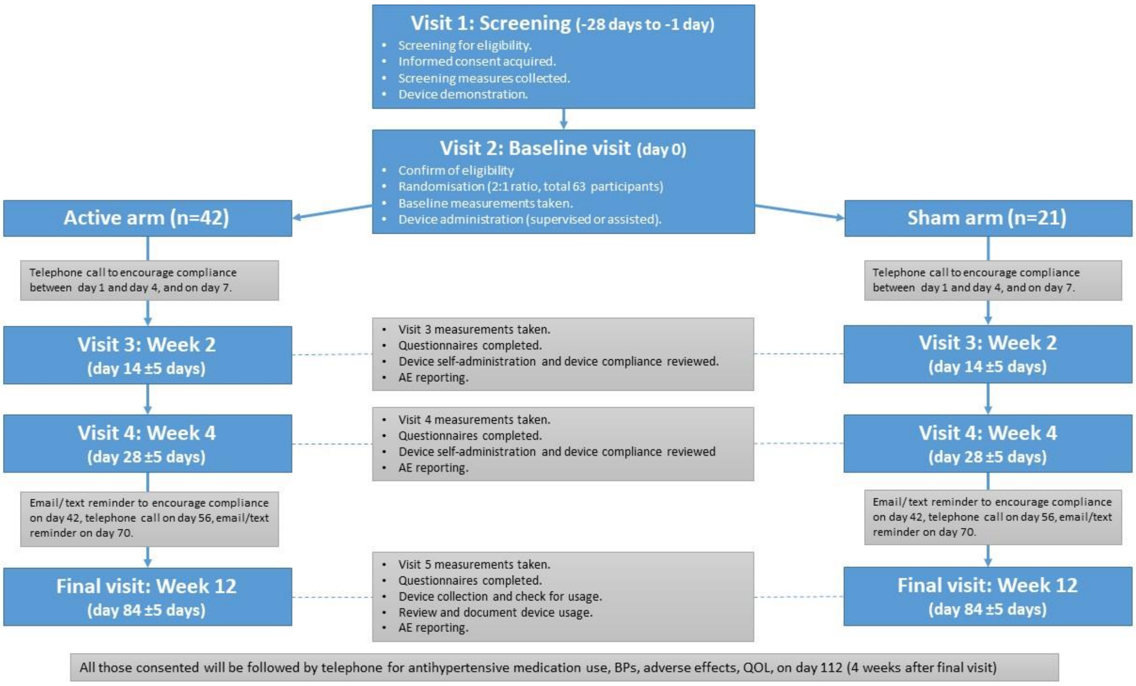

*Figure 1: Trial Flowchart*

##### 2.4.3 Patient withdrawal

Participants can withdraw from the study for appropriate medical reasons. This can be (but is not limited to) individual adverse events, new information gained about a treatment, or if it is felt to be in the participant's best interest.

Early termination of the participant from the trial could happen for the following reasons:

1. Withdrawal of consent
2. Development of physical or mental health condition that in the opinion of the participant or the investigator will not allow self-administration of the device stimulation
3. For other medical conditions that are temporary, and are not related to the device therapy, we will allow temporary disruption in the schedule at the discretion of the principal investigator and in consultation with the participant and his/her healthcare providers.

The number of withdrawals and the reasons for these withdrawals is considered a feasibility outcome and will be reported as part of a summary of trial feasibility.

#### 2.5 Recording of protocol deviations

Investigation site will record all (site level) protocol deviation on site deviation log. CVCTU will record all (coordinating delegated duty) protocol deviations on CVCTU deviation log. Respective committees (where deemed necessary) will receive a summary of all protocol deviations.

CI and CVCTU Coordinating team will assess non-compliances and will action a timeframe to deal with them (dependent on the severity), including whether there is a need to escalate to Sponsor, where required CI will report to Sponsor as per their guidelines.

Any event with the potential to affect participant safety or data integrity will be reported to the Sponsor within 24 hours of the CI or the CVCTU Coordinating team becoming aware.

The Sponsor will maintain a log of non-compliances to ascertain if there are any trends developing which need to be escalated.

A "serious breach" will be defined as something which is likely to affect to a significant degree:

- The safety or physical or mental integrity of the participants of the study, or
- The scientific value of the study

The CI will be responsible for reporting any potential serious breaches to the sponsor and to the CVCTU coordinating team within 24 hours of becoming aware of the event.

The sponsor will be responsible for determining whether a potential breach constitutes a serious breach and will work with the CI to investigate and notify and report to the MHRA and REC within 7 working days of becoming aware of the serious breach.

#### 2.6 Schedule of data collection

The schedule of treatment and data collection at each visit is described in the table below. More details are given in the CIP.

| Schedule | Visit 1 (Screening Visit) | Visit 2 ** (Baseline /Randomisation Visit) | Phone Call-1 | Phone Call-2 | Visit 3** | Visit 4** | Text / Email Reminder -1 | Phone Call-3 | Text / Email Reminder-2 | Visit 5** (End of Treatment Visit) | Phone Call-4 Follow-up* |
|---|---|---|---|---|---|---|---|---|---|---|---|
| **Timeline (weeks/days)** | n/a | Week 0 Day 0 | Day 1-4 | Week 1 Day 7 | Week 2 Day 14 | Week 4 Day 28 | Week 6 Day 42 | Week 8 Day 56 | Week 10 Day 70 | Week 12 Day 84 | Week 16 Day 112 |
| **Visit Window** | 28 days (-1 day) | n/a | n/a | +/- 3 days | +/- 5 days | +/-5 days | +/-3 days | +/-3 days | +/-3 days | +/-5 days | +/-3 days |
| **Informed Consent** | X | | | | | | | | | | |
| **Medical History, including demographic information and social history** | X | X[1] | | | | | | | | | |
| **Vital Signs[2]** | X | X | | | X | X | | | | X | |
| **Height & Waist Circumference** | X | | | | | | | | | | |
| **Weight & BMI** | X | X | | | | | | | | X | |

| | | | | | | | | | | | |
|---|---|---|---|---|---|---|---|---|---|---|---|
| **Concomitant Medication** | X | X | X | X | X | X | | X | | X | X |
| **24-hour ABPM** | X | X[3] | | | | X | | | | X | |
| **Office BP[4]** | X | X | | | X | X | | | | X | |
| **Central BP†** | | X | | | | | | | | X | |
| **24-hour Holter ECG** | | X*** | | | | X | | | | X**** | |
| **6-minute walk test (6MWT)** | | X | | | X | X | | | | X | |
| **Echocardiogram** | | X*** | | | | | | | | X**** | |
| **Electrocardiogram (ECG)** | X | | | | | | | | | | |
| **Blood Test[5]** | X | X | | | | X | | | | X | |
| **Blood Samples for storage and later evaluations (Plasma & Serum)** | | X | | | | X | | | | X | |
| **Urine Pregnancy** | X | X | | | | | | | | | |
| **Urine Sample[6]** | | X | | | | X | | | | X | |
| **ATONT Assessment†** | | X[7] | | | | | | | | X[8] | |
| **Inclusion and Exclusion** | X | X[9] | | | | | | | | | |

| | | | | | | | | | | | |
|---|---|---|---|---|---|---|---|---|---|---|---|
| **Randomisation** | | X | | | | | | | | | |
| **Device procedure and logbook** | | X[10] | X[11] | X[11] | X[12] | X[12] | X | X[11] | X[11] | X[12, 13] | |
| **Extent of Adherence scale (Voils[23]) Questionnaire** | | X | | | X | | | | | X | |
| **Insomnia Severity Index (ISI) Questionnaire** | | X | | | | X | | | | X | |
| **Blinding Questionnaire** | | | | | | X | | | | X | |
| **AffeX-CT Device Usability Questionnaire** | | | | | | X | | | | X | |
| **EQ-5D QoL Questionnaire** | | X | | | X | X | | | | X | X |
| **Cognitive Assessment** | | X | | | X | X | | | | X | |
| **AE Reporting** | X | X | X | X | X | X | | X | | X | X |

| | |
|---|---|
| * | All participants will be offered a safety reporting follow-up and permitted to change antihypertensive medication (if needed) after tAN procedure has ceased/ participants are not using the device. Participants will be encouraged to keep a record of their HR & |

| | |
|---|---|
| | BPs at home during this period however this is not mandatory. This follow-up is to assess and collate AffeX-CT device safety and efficacy prolonged data. |
| ** | All participants will be asked to **not** take their morning medications prior to the visits; they will be asked to bring their medication with them and administer their medication(s) in the Investigation Site, after their office BP measurements have been taken. If the participant has taken the medication prior to the visit, site staff will make a note on the medical files and eCRF of the participant. If the participant has not taken their medication to the site and did not receive their medication prior to the visit – and site staff cannot arrange to obtain the medication locally – the visit will be re-scheduled. |
| *** | Baseline Holter and/or Echocardiogram that is conducted within a period of -28 to +3 days from the randomization/baseline visit, will be allowed if there are no changes in the blood pressure treatment between the time of that investigation and randomization |
| **** | Visit 5 Holter and/or Echocardiogram that is conducted within a period of -5 to +14 days of the Visit 5 clinic date |
| † | To be complete for sub-study only. |
| X[1] | Any incomplete medical history sections to be reviewed and completed. |
| Vital signs[2] | Pulse rate, respiratory rate, temperature and oxygen saturation assessments. |
| X[3] | Screening 24-hour ABPM will be used at baseline, only if within screening period <u>and</u> no subsequent treatment changes have been made. Otherwise, 24-hour ABPM must be repeated at baseline (randomisation) visit. |
| Office BP[4] | 3 readings will be performed and average mean (after excluding first of 3 BP readings) calculated. |
| Blood Test[5] | 1 SST, 2 EDTA and 1 Oxalate; Full blood count (FBC), lipid profile, glucose (fasting), HbA1c, fructosamine, U&Es, Serum pregnancy for female participants on screening and randomisation visits. |
| Urine Sample[6] | Urinary albumin creatinine ratio and urinary antihypertensive drug screen. |

| | |
|---|---|
| Blood samples for storage and later evaluations (plasma and serum) | 2 SSTs and 1 EDTA; These will be collected and stored for future analysis, if need (i.e., in case new information arises or if there is a safety signal, and or further investigation is required. The stored samples will be used for the sub-study analysis in a batch basis, and will be used if there are any signals that will indicate that a more mechanistic or pathophysiological insight is required. |
| X[7] | For subgroup only - ATONT assessment on both arms will be completed with participants' consent at baseline visit and/or within 5 days after baseline & randomisation visit; Investigation site team will endeavour to complete assessment at baseline & randomisation visit. |
| X[8] | For subgroup only – ATONT assessment on both arms will be completed at end of treatment visit and/or within +/-5 days visit window. |
| X[9] | Inclusion and Exclusion criteria will be reviewed and any incomplete sections subsequent to screening visit will be completed. |
| X[10] | Device and training will be provided to the participant. Following training participants will be observed using the device during the visit to ensure they are competent with AffeX-CT device handling. Participants will be observed for 30 minutes after self-stimulation within the research facility, and their sitting blood pressure (X3 times) will be recorded before they are allowed to go home. |
| X[11] | Participants will be reminded to complete AffeX-CT device procedure and logbook entry. |
| X[12] | AffeX-CT device logbook will be reviewed. |
| X[13] | Participants will stop tAN Procedure and will return the device. |

Notes:

- Participants are required to complete device procedure daily from Day 0 to Day 14 and once weekly after Day 14 to Day 82.
- Participants will receive introduction to device and overview of how the device works with opportunity to demo and ask questions at screening visit.

### 3 Interim Analysis

The are no formal interim analyses planned during this pilot study, and hence no opportunity for the early termination of the study based on trial results from accumulating data.

The DSMC will meet to review accumulating data every 6 months. The DSMC will be presented with summary baseline data (patient characteristics and blood pressure measures) and the following summary outcome data:

- Daytime ambulatory SBP at 3 months/end of treatment (primary outcome)
- Daytime ambulatory DBP and heart rate at 3 months/end of treatment
- Daytime ambulatory SBP, DBP and heart rate at 1 month
- Office SBP, DBP and heart rate at 3 months/end of treatment
- Office SBP, DBP and heart rate at 1 month
- EQ-5D VAS score at 3 months/end of treatment
- Insomina Severity Index summary score at 3 months/end of treatment

This data will be presented as descriptive statistics only (mean and standard deviation or median and range) for each treatment group in the closed report only. Each outcome reported to the DSMC is chosen to be reflective of the completeness of key outcome measures but is not reflective of the chosen endpoint that will be reported at the end of the study.

The DSMC will also be presented with unblinded safety event data in the closed report.

### 4 Summary of Study Data

#### 4.1 Baseline Characteristics of Patient Cohort

Baseline data will be presented for the following factors, separately by randomised trial group:

- Age
- Sex
- BMI
- Smoking status
- Alcohol consumption
- Mean 24-hour, daytime, and night-time ABPM SBP and DBP (collected at screening unless required to be collected again)
- Office SBP and DBP
- Vital signs
- Holter ECG
- Blood test measures
- Urine test measures
- Echocardiogram measures
- 6MWT distance

The above data will be analysed using descriptive statistics such as mean and standard deviations or median and interquartile range if data is non-normally distributed. Categorical data will be presented as number (percentage). Descriptive statistics will be reported as overall participant and between the treatment groups. Any missing data will also be reported as percentages.

#### 4.2 Reporting of randomisation factors

The randomization process in this trial employed the dynamic minimization approach with the following factors as balancing criteria:

- Age at baseline: Categories: <65 years; 65+ years
- Sex: Categories: Male; Female
- BMI at baseline: Categories: <30 $kg/m^2$; 30+ $kg/m^2$
- Baseline mean daytime SBP (Systolic Blood Pressure): Categories: <160 mmHg; 160+ mmHg

**Descriptive Statistics**: For each of the randomization factors, the following descriptive statistics will be used to provide a summary of participant characteristics.

**Age at Baseline**:

Mean (± SD) for each category: <65 years, 65+ years.

Median (IQR) for each category: <65 years, 65+ years

Range for each category: <65 years, 65+ years

**Sex**:

Frequency and percentage of males and females in the study population.

**BMI at Baseline**:

Mean (± SD) for each category: <30 $kg/m^2$, 30+ $kg/m^2$

Median (IQR) for each category: <30 $kg/m^2$, 30+ $kg/m^2$

Range for each category: <30 $kg/m^2$, 30+ $kg/m^2$

**Baseline Mean Daytime SBP**:

Mean (± SD) for each category: <160 mmHg, 160+ mmHg.

Median (IQR) for each category: <160 mmHg, 160+ mmHg

Range for each category: <160 mmHg, 160+ mmHg

#### 4.3 Reporting of Longitudinal follow-up

##### 4.3.1 Treatment adherence

Compliance with the device usage is one of the acceptability criteria that is being evaluated and outcomes related to this are described in the Main Analysis (Section 5.2.2) below.

##### 4.3.0 Loss to follow-up and other missing data

In the event of patient withdrawals, the following procedures will be followed:

The specific reasons for patient withdrawals will be documented in detail, including any adverse events, lack of efficacy, or other reasons provided by the participants.

The number and percentage of withdrawals will be summarized for each treatment arm and overall.

Missing data will be dealt with in accordance with section 5.3.2 on handling missing data in this SAP.

##### 4.3.1 Protocol deviations

Investigation site will record all (site level) protocol deviation on site deviation log. CVCTU will record all (coordinating delegated duty) protocol deviations on CVCTU deviation log. Respective committees (where deemed necessary) will receive a summary of all protocol deviations.

CI and CVCTU Coordinating team will assess non-compliances and will action a timeframe to deal with them (dependent on the severity), including whether there is a need to escalate to Sponsor, where required CI will report to Sponsor as per their guidelines.

Any event with the potential to affect participant safety or data integrity will be reported to the Sponsor within 24 hours of the CI or the CVCTU Coordinating team becoming aware.

The Sponsor will maintain a log of non-compliances to ascertain if there are any trends developing which need to be escalated.

A "serious breach" will be defined as something which is likely to affect to a significant degree:

- The safety or physical or mental integrity of the participants of the study, or
- The scientific value of the study

The CI will be responsible for reporting any potential serious breaches to the sponsor () and to the CVCTU coordinating team () within 24 hours of becoming aware of the event.

The sponsor will be responsible for determining whether a potential breach constitutes a serious breach and will work with the CI to investigate and notify and report to the MHRA and REC within 7 working days of becoming aware of the serious breach.

The number of protocol deviations will be reported. Deviations that may impact data integrity will be described.

##### 4.3.2 CONSORT flowchart

Patient recruitment and retainment in the study will be described using a CONSORT flow diagram. The following will be included in the CONSORT diagram at minimum.

1) The number of patients consenting to trial participation
2) The number of patients randomised to each arm.
3) The number of patients withdrawn from the study and the reasons for withdrawal
4) The number of patients included in the statistical analysis.

**Example CONSORT diagram**

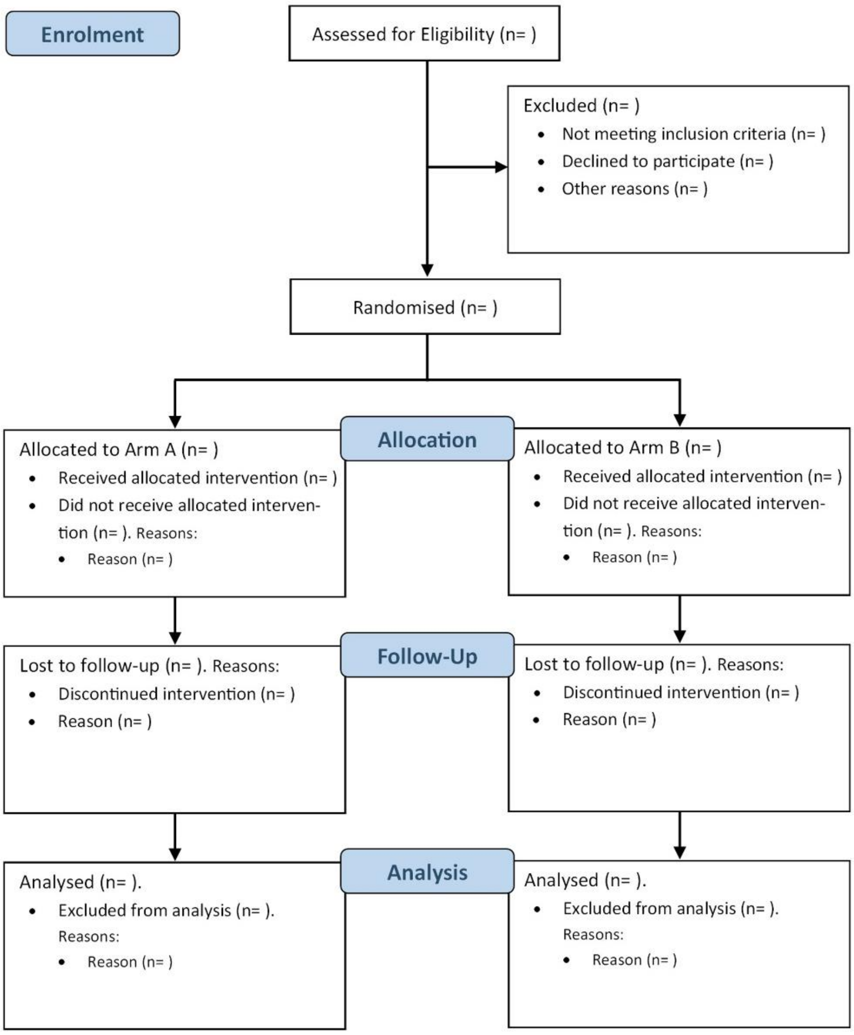

*Figure 2: Consort flow diagram*

#### 4.4 Adverse Event reporting

Adverse events (AEs) will be summarised using counts and percentages. The number of subjects having at least one AE will be presented overall and tabulated by treatment. The number of subjects with AEs of mild/moderate/severe intensity will be shown overall and by treatment group using the maximum severity experienced for each participant. The total number of AEs for each treatment group, allowing multiple events per participant, will also be presented.

Serious AEs (SAEs), both non-fatal and fatal will be listed separately along with details of the treatment and whether the event is unexpected and whether it is thought to be related to the treatment.

AEs and SAEs related to the intervention (Adverse device effects and Serious adverse device effects) will also be summarised using the methodology listed above.

#### 4.5 Data transformations

##### 4.5.1 Recording of blood pressure

Blood pressure is measured in a number of forms. Specific types of blood pressure being described is specified by each endpoint. Blood pressure measurements to be collected are as follows:

- Daytime ambulatory blood pressure: Blood pressure measured by an ambulatory blood pressure monitor. The measurement contributing to the endpoint is the average blood pressure recorded between 7am-11pm (or alternative awake hours if provided by the patient).
- 24hr ambulatory blood pressure: Blood pressure measured by an ambulatory blood pressure monitor. The measurement contributing to the endpoint is the average blood pressure recorded across the 24-hour monitoring period.
- Office blood pressure: Blood pressure measured by a typical static blood pressure monitor three times. The measurement contributing to the endpoint is the mean of the second and third collected measurements.

Blood pressure has been recorded on the eCRF as whole numbers. It is expected that blood pressure will follow a normal distribution despite this loss of granularity and as such average blood pressure will continue to be reported as a mean. This assumption of a normal distribution will be checked as part of standard model assumption checks.

###### 4.5.2 Endpoints assessing change in outcome

Many of the endpoints under investigation examine the change in continuous measures between baseline and a specified time point. The outcomes provided are typically mean SBP, DBP or HR measured over a certain specified period. The change in outcome will be calculated for each participant by subtracting the measure for the time point of interest from the baseline measurement. These changes in outcome will be reported for each treatment group and used in hypothesis testing described in Section 5.2.

###### 4.5.3 Coefficient of variation

The standard deviation of 24-hour ambulatory SBP and HR over a 24hr period for each individual is provided as is the mean SBP, DBP and HR. Office SBP and HR are provided for three readings from which a standard deviation and mean can be calculated. The coefficient of variation (SD/mean) in 24-hour ambulatory and office SBP and HR will be calculated for each individual at both the baseline and the end of treatment (3 month) timepoints.

###### 4.5.4 EuroQol 5 Dimension (EQ5D) questionnaire

The response level to each question will be coded according to the EQR-D guidelines and using the data dictionary accompanied our data and will be assigned correct weighting to each severity level. The EQ5-D questionnaire will be weighted according to the scoring guidelines and missing values recorded as 9. We will then create the EQ5-D dimensions using R code and function and calculate the sum of all individual dimensions utilities to create an overall EQ-5D score.

###### 4.5.5 Insomnia severity index (ISI) questionnaire

The response level to each question will be coded according to the ISI guidelines and using the data dictionary accompanied our data and will be assigned correct weighting to each severity level. Missing values will be recorded as 4 (maximum value for each question). An overall score will be calculated by combining the scores for each of the 7 questions. This overall score will be assigned to a severity (no clinically significant insomnia to severe clinical insomnia) based on the ISI guidelines.

### 5 Efficacy Analysis

#### 5.1 General details

We will be using R and/or Stata for the analyses. The analysis will be based on intention-to-treat and hypothesis testing will be carried out at the 5% (2-sided) significance level. Confidence intervals will be 95% and 2-sided.

Following the requirements of the CIP, a secondary analysis in the per-protocol population (consisting of those with good compliance as defined in Section 2.4.1: 80+% days of device usage) will be conducted.

A tertiary analysis with a stricter definition of a per-protocol population, consisting of those with 80% to 120% days of device usage, will also be considered. This is due to evidence of overcompliance with the treatment (>24 days of device usage) that arose during study recruitment.

All three analysis populations will be clearer indicated in all statistical reports.

#### 5.2 Main analysis

##### 5.2.1 Analysis of primary outcome

The primary objective of this trial is to assess the impact of tAN therapy on daytime systolic blood pressure (SBP) in subjects with uncontrolled hypertension compared to a placebo sham therapy.

Two effect sizes examining the impact of tAN therapy on daytime ABPM SBP will be reported – the change in average daytime ABPM SBP within each arm and the difference in change in average daytime ABPM SBP between study arms.

Average daytime ABPM SBP at baseline and the end of treatment (after 3 months of follow-up) will be presented as mean ± SD for each treatment arm. The mean change in average daytime ABPM SBP will be calculated for each treatment arm and presented with 95% confidence intervals. A paired t-test examining the change in daytime ABPM SBP within the intervention arm (the study arm of interest) has not been considered or specified in the CIP so will not be reported as part of the main analysis.

In addition, the crude difference in mean change in average daytime ABPM SBP will be calculated between trial groups and presented with a 95% confidence interval. Meeting the CIP requirements, a two-sample t-test will be used to test the null hypothesis of no difference in the change in average ABPM SBP from baseline to end of treatment between groups. The assumptions of the t-test will be examined to see whether a t-test with equal variance or one with unequal variance will be used.

Adjusted analysis will be conducted using linear regression comparing change in average ABPM SBP between treatment groups while adjusting for baseline daytime average ABPM SBP (ANCOVA), as well as adjusting for age, sex, and BMI). The estimated adjusted difference in change in SBP will be presented along with 95% confidence intervals (CIs) and p-values. The assumptions of the regression analysis are that the model residuals, i.e., the error terms (difference between predicted and observed values) have a normal distribution with mean zero and equal variance in both treatment groups. It is widely accepted that both SBP and change in SBP are normally distributed, hence that will be our underlying assumption. However, assessment of the distribution will be undertaken, and alternative non-parametric approach will be used as an alternative if the assumption of normality is considered to have been violated or if there are very extreme outliers observed. However, an accurate assessment of underlying distributional assumptions is limited in a small sample size such as this.

##### 5.2.2 Analysis of secondary outcomes

###### (1) Change in Average Daytime Ambulatory SBP and DBP from Baseline to 1 Month

*Descriptive statistics reported for each treatment arm:* Average daytime ABPM SBP and DBP at baseline and after 1 month of follow-up presented as mean ± SD.

*Effect sizes to be reported:* The mean change in average daytime ABPM SBP and DBP from baseline to 1 month within each arm reported with 95% confidence intervals as a within-arm effect size.

An effect size measuring the difference between groups (active-sham) in mean change in average daytime ABPM SBP and DBP will also be reported with 95% confidence intervals. This will be repeated to provide both a crude and an adjusted mean difference between trial arms.

*Analysis Method:* Two-sample t-tests as the main analysis. ANCOVA adjusting for baseline average daytime SBP/DBP, age, sex, and BMI to generate adjusted estimates for the difference between groups in mean change in average daytime ABPM SBP and DBP.

*Rationale:* These tests will assess if there is a significant difference in change in SBP and DBP from baseline to 1 month between treatment groups.

###### (2) Change in Average Daytime Ambulatory DBP from Baseline to the End of Treatment (3 Months)

*Descriptive statistics reported for each treatment arm:* Average daytime ABPM DBP at baseline and end of treatment (3 months of follow-up) presented as mean ± SD.

*Effect sizes to be reported:* The mean change in average daytime ABPM DBP from baseline to end of treatment within each arm reported with 95% confidence intervals as a within-arm effect size.

An effect size measuring the difference between groups (active-sham) in mean change in average daytime ABPM DBP will also be reported with 95% confidence interval. This will be repeated to provide both a crude and an adjusted mean difference between trial arms.

*Analysis Method:* Two-sample t-test as the main analysis. ANCOVA adjusting for baseline average daytime DBP, age, sex, and BMI to generate adjusted estimates for the difference between groups in mean change in average daytime ABPM DBP.

*Rationale:* Similar to endpoint 1, this test evaluates differences in changes in DBP between treatment groups over the course of the treatment period.

##### (3) Controlled BP at the End of Treatment (3 Months)

*Descriptive statistics reported for each treatment arm:* Number and proportion with controlled BP (defined as mean ABPM SBP <135mmHg and mean daytime ABPM DBP <85mmHg) at end of treatment.

*Effect sizes to be reported:* An absolute effect size of the difference between groups (active-sham) in proportion of patients with controlled BP with 95% confidence interval.

Relative effect size will be reported as odds ratios (active/sham) for the odds of having controlled BP at the end of the treatment. This will be repeated to provide both a crude and an adjusted odds ratio between trial arms.

*Analysis Method:* Chi-square test for between-group comparison as the main analysis. Logistic regression adjusting for age, sex, and BMI to generate adjusted odds ratios for the odds of having controlled BP at the end of treatment.

*Rationale:* This test will compare the proportions of subjects achieving controlled BP between the treatment and control groups at the end of the treatment period.

##### (4) Change in Average 24-Hour Ambulatory SBP and DBP from Baseline to the End of Treatment (3 Months)

*Descriptive statistics reported for each treatment arm:* Average 24hrABPM SBP and DBP at baseline and end of treatment (3 months of follow-up) presented as mean ± SD.

*Effect sizes to be reported:* The mean change in average 24hr ABPM SBP and DBP from baseline to end of treatment within each arm reported with 95% confidence intervals as a within-arm effect size.

An effect size measuring the difference between groups (active-sham) in mean change in average 24hr ABPM SBP and DBP will also be reported with 95% confidence interval. This will be repeated to provide both a crude and an adjusted mean difference between trial arms.

*Analysis Method:* Two-sample t-tests as the main analysis. ANCOVA adjusting for baseline average 24hr SBP/DBP, age, sex, and BMI to generate adjusted estimates for the difference between groups in mean change in average 24hr ABPM SBP and DBP.

*Rationale:* These tests assess differences in changes in 24-hour SBP and DBP from baseline to the end of the treatment period between treatment groups.

##### (5) Change in Average Office SBP and DBP from Baseline to 1 Month, and from Baseline to the End of Treatment (3 Months)

*Descriptive statistics reported for each treatment arm:* Average office SBP and DBP at baseline and 1 month of follow-up presented as mean ± SD.

*Effect sizes to be reported:* The mean change in average office SBP and DBP from baseline to end of treatment within each arm reported with 95% confidence intervals as a within-arm effect size.

An effect size measuring the difference between groups (active-sham) in mean change in average office SBP and DBP will also be reported with 95% confidence intervals. This will be repeated to provide both a crude and an adjusted mean difference between trial arms.

*Analysis Method:* Two-sample t-tests as the main analysis. ANCOVA adjusting for baseline average office SBP/DBP, age, sex, and BMI to generate adjusted estimates for the difference between groups in mean change in average office ABPM SBP and DBP.

*Rationale:* Similar to endpoint 1, these tests evaluate differences in the change in average office measured SBP and DBP between treatment groups.

##### (6) Change in Average Daytime Ambulatory HR, and in Average Night-Time Ambulatory HR from Baseline to the End of Treatment (3 Months)

*Descriptive statistics reported for each treatment arm:* Average daytime ambulatory HR and average nighttime ambulatory HR at baseline and end of treatment (3 months of follow-up) presented as mean ± SD

*Effect sizes to be reported:* The mean change in average daytime and nighttime ambulatory HR from baseline to end of treatment within each arm reported with 95% confidence intervals as a within-arm effect size.

An effect size measuring the difference between groups (active-sham) in mean change in daytime ambulatory HR and average nighttime ambulatory HR will also be reported with 95% confidence intervals. This will be repeated to provide both a crude and an adjusted mean difference between trial arms.

*Analysis Method:* Two-sample t-tests as the main analysis. ANCOVA adjusting for baseline average daytime/nighttime ambulatory HR, age, sex, and BMI to generate adjusted estimates for the difference between groups in mean change in daytime ambulatory HR and nighttime ambulatory HR.

*Rationale:* These tests assess difference in changes in heart rate (HR) from baseline to the end of the treatment period between treatment groups.

**(7) Change in BP Variability (Coefficient of Variation) of 24-Hour Ambulatory SBP, and of Within-Visit Office SBP from Baseline to the End of Treatment (3 Months)**

*Descriptive statistics reported for each treatment arm:* Coefficient of variation for 24hr ambulatory SBP and within office SBP at baseline and end of treatment (at 3 months of follow-up) presented as median (interquartile range).

*Effect size to be reported:* The median change in coefficient of variation for 24hr ambulatory SBP and office SBP from baseline to end of treatment within each arm reported as a within-arm effect size.

An effect size measuring the difference between groups (active-sham) in median change in coefficient of variation for 24hr ambulatory SBP and office SBP.

*Analysis Method:* Mann Whitney U tests for between group comparison.

*Rationale:* These tests evaluate differences in changes in blood pressure variability between each treatment group. Coefficient of variation is predicted to be non-normally distributed so an appropriate test without the normality assumption has been selected.

**(8) Change in HR Variability (Coefficient of Variation) of 24-Hour Ambulatory HR, and of Within-Visit Office HR from Baseline to the End of Treatment (3 Months)**

*Descriptive statistics reported for each treatment arm:* Coefficient of variation for 24hr ambulatory HR and within office HR at baseline and end of treatment (at 3 months of follow-up) presented as median (interquartile range).

*Effect size to be reported:* The median change in coefficient of variation for 24hr ambulatory HR and office HR from baseline to end of treatment within each arm reported as a within-arm effect size.

An effect size measuring the difference between groups (active-sham) in median change in coefficient of variation for 24hr ambulatory HR and office HR.

*Analysis Method:* Mann Whitney U tests for between group comparison.

*Rationale:* Similar to endpoint 7, these tests assess differences in changes in heart rate variability between treatment groups using a measure that is not expected to be normally distributed.

**(9) Occurrence of a Serious Adverse Event (SAE) within 3 Months**

*Descriptive statistics reported for each treatment arm:* Number and proportion of patients experiencing an SAE within 3 months.

*Effect sizes to be reported:* An absolute effect size of the difference between groups (active-sham) in proportion of patients experiencing an SAE with 95% confidence interval.

Relative effect size will be reported as odds ratios (active/sham) for the odds of having a SAE within 3 months. This will be repeated to provide both a crude and an adjusted odds ratio between trial arms.

*Analysis Method:* Chi-square test for between-group comparison as the main analysis. Logistic regression adjusting for age, sex, and BMI to generate adjusted odds ratios for the odds of experiencing an SAE by the end of treatment.

*Rationale:* This test compares the incidence of SAEs between the treatment and control groups over the 3-month period.

##### (10) Occurrence of a Major Cardiovascular Event (MACE) within 3 Months

*Descriptive statistics reported for each treatment arm:* Number and proportion of patients experiencing a MACE within 3 months.

*Effect size to be reported:* An absolute effect size of the difference between groups (active-sham) in proportion of patients experiencing a MACE with 95% confidence interval.

Relative effect size will be reported as odds ratios (active/sham) for the odds of having a MACE within 3 months. This will be repeated to provide both a crude and an adjusted odds ratio between trial arms.

*Analysis Method:* Chi-square test for between-group comparison as the main analysis. Logistic regression adjusting for age, sex, and BMI to generate adjusted odds ratios for the odds of experiencing a MACE by the end of treatment.

*Rationale:* This test compares the incidence of MACE between the treatment and control groups over the 3-month period.

##### (11) Change in Quality of Life (EuroQol Visual Analogue Score and EQ5D) between Baseline and the End of Treatment (3 Months)

*Descriptive statistics reported for each treatment arm:* EuroQol VAS and EQ5D score at baseline and end of treatment (at 3 months of follow-up) presented as median (interquartile range).

*Effect size to be reported:* The median change in EuroQol VAS and EQ5D score from baseline to end of treatment within each arm reported as a within-arm effect size.

An effect size measuring the difference between groups (active-sham) in median change in EuroQol VAS and EQ5D score.

*Analysis Method:* Mann Whitney U tests for between group comparison.

*Rationale:* These tests evaluate the difference in changes in quality-of-life scores between treatment groups over the 3-month period.

**(12) Change in Sleep Quality (Insomnia Severity Index) between Baseline and the End of Treatment (3 Months)**

*Descriptive statistics reported for each treatment arm:* ISI overall score at baseline and end of treatment (at 3 months of follow-up) presented as median (interquartile range).

*Effect size to be reported:* The median change in ISI overall score from baseline to end of treatment within each arm reported as a within-arm effect size.

An effect size measuring the difference between groups (active-sham) in median change in ISI overall score.

*Analysis Method:* Mann Whitney U tests for between group comparison.

*Rationale:* Similar to endpoint 11, this test assesses differences in change in sleep quality scores between treatment groups over the 3-month period.

**(13) Adherence to Trial Therapy (Proportion of Days) over 3 Months:**

*Descriptive statistics reported for each treatment arm:* Proportion of days out of the total days in follow-up when therapy was self-administered will be presented as median (interquartile range).

*Effect size to be reported:* Difference between groups (active-sham) in median proportion of days when therapy was self-administered.

*Analysis Method:* Mann Whitney U tests for between group comparison.

*Rationale:* This test assesses differences in adherence between treatment groups.

**(14) Adherence to Trial Therapy (Average Daily Duration) over 3 Months:**

*Descriptive statistics reported for each treatment arm:* Average daily duration of therapy self-administration presented as mean ± SD.

*Effect size to be reported:* Difference between groups (active-sham) in mean daily duration of therapy self-administration with 95% confidence interval.

*Analysis Method:* Descriptive statistics and potentially t-tests or non-parametric tests for between-group comparison.

*Rationale:* This test assesses differences in adherence between treatment groups.

#### 5.3 Statistical considerations

##### 5.3.1 Model assumption checks

Two-sample t-tests will be used if the data is normally distributed and Mann Whitney U tests if the distribution is not normal. Normality of the data will be checked using Q-Q plots for all outcomes.

The test used may be changed from a two-sample t-test to a Mann Whitney U test depending on the appearance of the Q-Q plot.

After the checks for normality, equality of variances will be explored by examining the standard deviation in each group for each relevant outcome. If equal variance is deemed to be an appropriate assumption, two-sample t-tests assuming equal variances will be used. If it is deemed an inappropriate assumption, two-sample t-tests assuming unequal variances will be used.

##### 5.3.2 Methods for handling missing data

This pilot study will assess data completeness to inform decisions about missing data for a future main trial. Analysis will use all participants with available endpoint data. The level of missing data will be tabulated for each outcome, presenting the number of subjects with a data record and number with missing data, separately by trial arm.

Each endpoint will be analysed on a complete case basis.

##### 5.3.3 Methods for handling outliers

**Handling Outliers and Exclusion Criteria:** Outliers, defined as values that deviate substantially from the rest of the dataset, may be identified, and considered for exclusion from the analysis. The following steps will be taken:

**Identification of Outliers**: Outliers will be identified through the examination of the data distribution. Potential outliers will be assessed using statistical methods such as box plots, z-scores, or any other appropriate techniques specific to the nature of the data.

**Definition of Outliers**: Outliers will be defined as values greater than 3 standard deviations from the mean.

**Documentation and Reporting**: The presence of outliers and the criteria used for their identification and potential exclusion will be thoroughly documented. This documentation will include a description of the statistical techniques employed and the rationale for the chosen criteria.

**Sensitivity Analyses**: Sensitivity analyses will be conducted to assess the impact of including or excluding outliers on the study results. These analyses will provide insights into the robustness of the findings to outlier handling procedures.

The handling of outliers will be carried out in a transparent and systematic manner, ensuring that any exclusions are based on scientifically justified criteria. The final analysis will be conducted with and without the inclusion of outliers, and the results of both analyses will be reported.

###### 5.3.4 Methods for handling multiple comparisons

No adjustment will be made for multiple comparisons.

###### 5.3.5 Methods for handling non-adherence to treatment and protocol deviations

As this is a pilot study and adherence is examined as a secondary endpoint, no adjustments for non-adherence to treatment or protocol deviations are planned.

In scenarios where high levels of protocol deviations are witnessed, an alternative analysis population, distinct from the intention-to-treat or per-protocol populations, may be explored as a secondary sensitivity analysis.

The alternative analysis population may consist of a subgroup defined by specific criteria, such as:

- Excluding participants with substantial protocol deviations.
- Including only participants who have demonstrated a high level of adherence to the assigned intervention.

The rationale for investigating an alternative analysis population will be provided, and any predefined criteria for subgroup selection will be outlined. This secondary sensitivity analysis aims to assess the robustness of the primary analysis results to variations in adherence and protocol compliance.

The results of any secondary analysis using an alternative analysis population will be interpreted in conjunction with the main analyses, providing valuable insights into the potential impact of non-adherence or protocol deviations on the study outcomes.

##### 5.4 Planned additional analyses.

###### 5.4.1 Alternative statistical methods for analysing the primary and secondary endpoints

Alternative statistical methods for analysing the primary and secondary endpoints may be used in addition to the main analysis described in Section 5.2. These alternative methods are meant to compliment the main analysis and interpretations regarding statistical significance between groups or timepoints should not be made based on the results of these alternative methods alone.

Alternative statistical methods will include calculating the Area Under the Curve for continuous endpoints measured at baseline, 1 month and 3 months of follow-up (e.g. ABPM and office blood pressure). Area Under the Curve will be compared between treatment arms or subgroups using unpaired t-tests assuming unequal variance.

Any alternative statistical methods performed will be clearly indicated and the rationale for the use of this additional method will be clearly described.

##### 5.4.2 Analysis of exploratory/feasibility outcomes

Exploratory endpoints are defined in the CIP as follows:

- Change in left ventricular ejection fraction (LVEF), left ventricular mass (LVM), relative mass thickness (RMT), left atrial volume (LAV), left ventricular end-diastolic pressure (LVEDP), and E/e´ ratio from baseline to the end of treatment (3 months), in participants receiving treatment with tAN therapy.
- Changes in anti-hypertensive medication between baseline and the end of follow-up (4 months) in participants receiving treatment with tAN therapy.
- Average daily number of antihypertensive medications that participants are on assessed through urinary drug screening (UDS).
- Change in Quality of life between baseline and the end of follow-up (4 months) in a sub-group of participants receiving treatment with tAN therapy, using the EuroQol Visual Analogue score (0-100), and the EuroQol 5 Dimension (EQ5D) quality of life questions.
- Change in average central BP, measured by Sphygmocor Vx device, from baseline to the end of treatment (3 months) – only for sub study participants.
- Change in the cognitive functions from baseline to the end of treatment (3 months)
- Central respiratory modulation of cardiac vagal tone.
- Overall baroreflex gain (central baroreflex responsiveness).
- Cardiodepressor and vasodepressor responses of the carotid sinus to digital massage.

Feasibility endpoints are defined in the CIP as follows:

- AffeX-CT device-related adverse events within 3 months.
- Adherence to use of the AffeX-CT device assessed using the Extent of Adherence (EoA) questionnaire
- Ease of use of AffeX-CT device assessed using a participant feedback visual analogue scale (VAS).
- Reasons behind any early withdrawals from the study.
- Success of the blinding procedure assessed using a blinding index at 1 month and 3 months

All continuous outcomes (e.g. LVEF, central BP) with an endpoint examining a change between baseline and end of treatment/follow-up will be presented as mean ± SD for each treatment arm at each timepoint. The mean change in endpoint will be calculated for each treatment arm and presented with 95% confidence intervals as a within-arm effect size. In addition, the crude difference in mean change in endpoint will be calculated between trial groups and presented with a 95% confidence interval to represent the between-arm effect size. Unpaired t-tests with unequal variances will be used to compare the change in outcome between treatment arms copying the strategy used for the primary endpoint. If an endpoint is non-normally distributed, Mann-Whitney U tests will be used instead.

The endpoint related to quality of life measured using the EuroQol Visual Analogue score (0-100), and the EuroQol 5 Dimension (EQ5D) quality of life questions will be reported as described for the comparable secondary endpoint (endpoint 11). EuroQol VAS and EQ5D score at baseline and end of follow-up (at 4 months of follow-up) will be presented as median (interquartile range) for each treatment arm. The median change in EuroQol VAS and EQ5D score from baseline to end of follow-up within each arm will be reported as a within-arm effect size. The difference between groups (active-sham) in median change in EuroQol VAS and EQ5D score will also be calculated as a between-arm effect size. Mann Whitney U tests will be used to compare the change in outcome between treatment arms.

Categorical outcomes (e.g. number of anti-hypertensive medications, most questionnaires) will be presented as number (percentage) in each category for each treatment arm at all timepoints relevant to the endpoint. The change in proportion experiencing the least severe and the most severe category (or all categories if not ordered) within each arm will be reported as a within-arm effect size. The difference between groups (active-sham) in median change in proportion will also be calculated as a between-arm effect size. Mann Whitney U tests or chi-squared tests will be used to compare the change in outcome between treatment arms.

##### 5.4.3 Exploratory analysis of collected study data not defined as a study endpoint

In addition to the study endpoints, data for several other outcomes of interest have been collected over the course of study follow-up. These outcomes are described in the CIP and in the schedule of data collection (Section 2.6) but are not clearly defined as study endpoints. These outcomes include:

- Change in distance walked during a 6 minute walk test from baseline to the end of treatment (3 months)
- Change in sleep quality VAS scale (collected as part of cognitive assessments) from baseline to the end of treatment (3 months)

Analysis of these outcomes will be completed alongside the analysis of the study endpoints. As these outcomes are not defined study endpoints and the study has not been powered to analyse these outcomes all analysis will be considered exploratory.

The outcomes will be analysed using the same methodology as described in Section 5.4.2.

##### 5.4.4 Subgroup analyses

The primary endpoint will be analysed according to the following subgroups:

- Device compliance (<80% vs 80-120% vs >120% days of device usage)
- BMI at baseline (<30 kg/m$^2$ vs. ≥30 kg/m$^2$)
- Diabetes status at baseline
- Age at baseline (<65 years vs. ≥65 years)
- Mean ABPM daytime SBP at baseline (<160 mmHg vs. ≥160 mmHg).

The analysis will be done using an ANCOVA model where our outcome is change in mean SBP and we will also include interaction terms for each of the above 4 subgroup categories and treatment arms.

For adjusted analysis of the primary end point the statistical model will be adjusted for baseline characteristics such as age at baseline, Sex, BMI and baseline SBP.

##### 5.4.5 Sensitivity analysis

Sensitivity analysis will be performed by testing the robustness of the statistical methods used by

- Varying covariates in the model for example adding or removing variables and checking their effect.
- Using different interaction terms
- Missing data: for example, analysing only complete cases and also using multiple imputations or pattern mixed models to impute the data and redoing the analysis.
- We can also check the influence of outliers.
- The ANCOVA model assumptions will be checked by looking at the data distribution and if the assumptions are violated then we will consider non-parametric methods.

Any sensitivity analysis performed will be clearly indicated and the rationale for the sensitivity analysis will be clearly described.

###### *5.4.5.1 Device and medication compliance*

One group of covariates of particular interest will be compliance with both the tAN device that is the study intervention, and anti-hypertensive medications that participants were taking prior to study onset. When adjusted models (e.g. linear regression or ANCOVA) have been used in the main analysis above, additional statistical models (generalised linear mixed models) including covariates accounting for device and medication compliance will be completed as well to account for the varying compliance seen during study follow-up.

The added covariates for device and medication compliance will be allowed to vary over time with compliance levels indicated at each study visit (visit 3, 4, and 5). Other covariates included in the adjusted models in the main analysis will also be included in the generalised linear mixed model in the form used in the main analysis (fixed across time). Alternate version of the other covariates may be considered if data for these covariates is available at each study visit and has been shown to vary over time.

Two forms of covariates to indicate compliance will be considered. Either binary or categorical covariates of compliance (e.g. compliant/non-compliant indicators) or continuous covariates (e.g. % days of device usage or number of anti-hypertensive medications being taken). The final chosen form of the covariates will be selected based on whether a linear relationship between daytime SBP (primary outcome) and the mean compliance level (as a continuous variable) exists. The same form of the covariates will be used for all secondary outcomes. This criterion is necessary to ensure this extended sensitivity analysis meets the model assumptions of generalised linear mixed models.

### SCRATCH_SAP_v2.0_22042025_clean

Final Audit Report 2025-05-22

| | |
|---|---|
| Created: | 2025-05-20 (Greenwich Mean Time) |
| By: | Annastazia Learoyd |
| Status: | Signed |
| Transaction ID: | CBJCHBCAABAAy9gxgt0qdW5uJKLQMhWDkBRh5rzd4vXZ |

#### "SCRATCH_SAP_v2.0_22042025_clean" History

Document created by Annastazia Learoyd
2025-05-20 - 2:30:33 PM GMT

2025-05-20 - 2:31:18 PM GMT

Document e-signed by Annastazia Learoyd
Signature Date: 2025-05-20 - 2:31:28 PM GMT - Time Source: server

2025-05-20 - 2:31:31 PM GMT

2025-05-20 - 2:43:02 PM GMT

Document e-signed by Ajay Gupta
Signature Date: 2025-05-22 - 3:31:02 PM GMT - Time Source: server

Agreement completed.
2025-05-22 - 3:31:02 PM GMT
