## Supplementary material for "A Sham Controlled Randomised Trial evaluating the Safety, Acceptability and Efficacy of Autonomic neuromodulation using Trans-Cutaneous vagal sensory stimulation in uncontrolled Hypertensive patients: Rationale and study design of the SCRATCH-HTN Study": Clinical Investigation Plan

### Clinical Investigational Plan for Medical Device Studies

| | |
|---|---|
| **Full Title** | **S**ham **c**ontrolled **R**andomized Control Trial evaluating the Safety, Acceptability and Efficacy of **A**utonomic neuromodulation using **t**rans-**c**utaneous vagal sensory stimulation in uncontrolled **h**ypertensive patients: a pilot study evaluating a novel non-invasive device-based strategy. |
| **Short Title** | **SCRATCH–HTN** |
| **Sponsor** | **Queen Mary University of London**<br><br>Contact person:<br>Mays Jawad<br>Research Governance Operations Manager<br><br>Joint Research Management Office (JRMO)<br><br>Barts Health NHS Trust & Queen Mary University of London<br><br>Research Services \| Dept. W \| 69-89 Mile End Rd \| London \| E1 4UJ T: (020) 7882 7207 \| \| W: www.jrmo.org.uk<br><br>Department inbox: |
| **IRAS Number** | 302061 |
| **REC Reference** | 21/WS/0157 |
| **Sponsors Number** | 2358563 |
| **Chief Investigator** | Dr Ajay Gupta<br>Clinical Reader and Consultant<br>Barts Cardiovascular Clinical Trials Unit (CVCTU)<br>William Harvey Research Institute – Heart Centre<br>Barts and The London School of Medicine and Dentistry, Queen Mary University of London, Charterhouse Square<br>London EC1M 6BQ<br><br><br>Phone number: 020 7882 2858 |

### Trial Contacts

| | |
|---|---|
| **Investigation Site** | William Harvey Clinical Research Centre<br>William Harvey Research Institute<br>Charterhouse Square<br>Queen Mary University of London<br>London EC1M 6BQ |
| **Sub-Investigators** | Dr David Collier<br>Director of William Harvey Clinical Research Centre<br>Deputy Director, Barts CTU, Wolfson Institute of Preventive Medicine, Barts & The London Faculty of Medicine & Dentistry, Queen Mary University of London Charterhouse Square, London EC1M 6BQ<br>Phone Number: 07961 383925<br><br><br>Dr Manish Saxena<br>Deputy I Director<br>William Harvey Clinical Research Centre, Barts & London Faculty of Medicine & Dentistry, Queen Mary University of London, Charterhouse Square<br>London EC1M 6BQ<br>Phone number: 020 7882 3856<br><br><br>Professor Amrita Ahluwalia<br>Dean for Research<br>Director of the Barts Cardiovascular CTU<br>Prof of Vascular Pharmacology<br>William Harvey Research Institute,<br>Barts & London Faculty of Medicine & Dentistry,<br>Queen Mary University of London, Charterhouse Square<br>London EC1M 6BQ<br>Phone number: 020 7882 8377<br><br><br>Dr Mamta Bajre<br><br>Health Innovation Oxford and Thames Valley was formerly the Oxford Academic Health Science Network (Oxford AHSN), Oxford University NHS Foundation Trust<br>Magdalen Centre North,<br>Oxford Science Park,<br>Robert Robinson Ave, Littlemore,<br>Oxford OX4 4GA<br> |
| **Clinical Trials Unit / Trial Manager / Trial Coordinator** | Barts Cardiovascular Clinical Trials Unit (CVCTU)<br>Barts Clinical Trials Unit (No. 4)<br>William Harvey Research Institute – Heart Centre |

| | |
|---|---|
| | Barts and The London Faculty of Medicine and Dentistry, Queen Mary University of London, Charterhouse Square London EC1M 6BQ<br>Phone number: 020 7882 6838<br> |
| **Funder** | National Institute for Health Research<br>Invention for Innovation i4i<br>Ref NIHR202116<br>39 Victoria Street, Westminster, London SW1H 0EU |
| **Device Manufacturer** | Afferent Medical Solution Ltd<br>Dr Everard Mascarenhas<br>C/O Clockwise , Brunel House, Fitzalan Road, Cardiff, Wales, CF24 0EB<br>Phone number: 07815444305<br> |
| **Trial Statistician** | Annastazia Learoyd<br>Barts Cardiovascular Clinical Trials Unit (CVCTU), William Harvey Research Institute , Centre of Cardiovascular Medicines & Devices,<br>Barts and The London Faculty of Medicine and Dentistry, Queen Mary University of London, Charterhouse Square, London EC1M 6BQ<br> |
| **List of laboratories** | Professor Amrita Ahluwalia<br>William Harvey Research Institute, Centre of Centre of Cardiovascular Medicines & Devices, Queen Mary University of London, Charterhouse Square London EC1M6BQ<br>Phone number: 020 7882 8377<br> |
| **List of Patient Identification Centres (PICs)** | **Imperial College Healthcare NHS trust and associated hospitals.**<br>Prof Neil Poulter, Professor and Consultant Physician, Peart Rose Clinic, Hammersmith Hospital, Imperial NHS Healthcare Trust<br>**St George's University Hospitals NHS Foundation Trusts.**<br>Dr Teck Khong, Consultant, St George's Hospital NHS Trust,<br>**Homerton University Hospital Foundation Trust.**<br>Dr Louise Abrams, Consultant, Homerton Hospital.<br>**University College London Hospitals NHS Foundation Trust**<br>Dr Marc George, Consultant, Blood pressure clinic, University College London Hospitals NHS Trust,<br>**Royal Free London NHS Foundation Trust**<br>Dr Stephen Walsh, Associate Professor and |

| | |
|---|---|
| | Consultant Nephrologist, Royal Free Hospital<br><br>**King's College London NHS Foundation Trust**<br>Dr Andrew Webb, Clinical Senior Lecturer, King's College London NHS Foundation Trust.<br><br>**Broomfield Hospital, Mid and South Essex Hospitals Foundation NHS Trust**<br>Dr Tehreem Butt, Consultant in Acute Medicine & Clinical Pharmacology/Hypertension, Broomfield Hospital<br> |
| **List of central facilities** | **Sealed Envelope**<br>Peter Madden<br>501 Clerkenwell Workshops<br>27-31 Clerkenwell Close<br>London EC1R 0AT<br><br><br>**Castor**<br>Joanna Jedrzejczak<br>35 Ludgate Hill<br>London EC4M7JN<br><br><br>**Barts Bioresource**<br>Queen Mary University of London<br>William Harvey Research Institute<br>1 St Martin Le-Grand<br>London EC1A 4NP<br>02078826903<br> |
| **Committees** | **Trial Management Committee (TMG)**<br>Role: To discuss all aspects of the trial progression<br><br>**Trial Steering Committee (TSC)**<br>Role: To provide overall supervision of the trial and ensure that it is being conducted according to the CIP, GCP and relevant regulations. To monitor trial progress in relation to recruitment, data capture and completeness, protocol deviations and participant withdrawals.<br><br>**Data Safety Monitoring Committee (DSMC)**<br>Role: To review the trial data throughout the study and assess whether there are any safety issues that need to be brought to the attention of the TSC, or any ethical reasons why the trial should not continue.<br><br>**Research Steering Group (RSG)**<br>Role: To monitor the performance and technical content of the study and access the result of the study. To identify and address any weaknesses or delays in the trial. To operate as the key forum of the trial reporting to the funder. |

| | |
|---|---|
| | For further details please see respective committee charters. |
| **Key CIP Contributor (s)** | Dr Ajay Gupta<br>Chief Investigator<br>Barts Cardiovascular Clinical Trials Unit (CVCTU)<br>William Harvey Research Institute, Barts and The London Faculty of Medicine and Dentistry, Queen Mary University of London, Charterhouse Square Campus, London EC1M 6BQ<br><br>Jane Field<br>CVCTU Clinical Trials Coordinator<br>Barts Cardiovascular Clinical Trials Unit (CVCTU)<br>William Harvey Research Institute Centre of Cardiovascular Medicines & Devices, Barts and The London School of Medicine and Dentistry, Queen Mary University of London, Charterhouse Square Campus, London EC1M 6BQ<br><br>Thomas Godec<br>(Previous CVCTU Statistician)<br>Barts Cardiovascular Clinical Trials Unit (CVCTU)<br>William Harvey Research Institute, Centre of Cardiovascular Medicines & Devices, Barts and The London Faculty of Medicine and Dentistry, Queen Mary University of London, Charterhouse Square<br>London EC1M 6BQ<br><br>Annastazia Learoyd<br>(Current CVCTU Statistician)<br>Barts Cardiovascular Clinical Trials Unit (CVCTU)<br>William Harvey Research Institute, Centre of Cardiovascular Medicines & Devices, Barts and The London Faculty of Medicine and Dentistry, Queen Mary University of London, Charterhouse Square<br>London EC1M 6BQ |

**Summary of Changes**

| Version Number | Version Date | Amendment Number | Summary of Changes |
|---|---|---|---|
| V1.0 | 12 Oct 2021 | N/A | N/A |
| V2.0 | 14 Dec 2021 | 1 | Added: exclusion criteria 3 and 14, clarification about cognitive assessment, clarified that urine and serum pregnancy tests will be done at screening and baseline visits (for females only), reference 21 |
| V3.0 | 24 Dec 2021 | 2 | Updated the device labels and the pulse intensity. Updated inclusion criterion 4. |
| V4.0 | 29 Dec 2021 | 3 | Added clarification to instruction for use of the device. Labels updated. Exclusion criteria 22 |

| | | | |
|---|---|---|---|
| | | | added. Clarification on risk management. Extra visit description. |
| V5.0 | 05 Jan 2022 | 4 | Modified the wording of Exclusion criterion 2. |
| V6.0 | 06 Jan 2022 | 5 | Added details about BP monitoring during the study under the section 'Assessment and Management of risk' |
| V7.0 | 10 May 2022 | 6 (SA 01) | Updated sponsor's contact details, added ECG for screening visit on the schedule of assessments, updated blood volumes. |
| V8.0 | 01 December 2022 | 7 (SA 01) | • Updated Inclusion criterion 4 to increase the maximum number of antihypertensive medications patients can be taking at screening or baseline from 3 to 4.<br>• Updated inclusion criterion 6 to simplify definition of uncontrolled blood pressure solely based on the gold-standard measure, daytime average on ABPM and have removed the additional requirement for office BP being uncontrolled in addition to the daytime average on ABPM.<br>• Updated inclusion criterion 7c to refer to any one of three HR recordings as opposed to an average of the recordings, to simplify inclusion criteria.<br>• Updated exclusion criterion 5 to refer to eGFR at the screening visit only and remove the statement 'during the three months prior to randomisation' to simplify and make this criterion more objective.<br>• Updated section on reporting device related AEs to manufacturer to clarify that device related SAEs will be reported as soon as possible and device related AEs will be reported on an ongoing monthly basis.<br>• Added blood tube to blood test footnote, there is no change to total blood volume.<br>• Added two additional PIC sites, King's College London NHS Foundation Trust and Royal Free Hospitals NHS Foundation Trust.<br>• Added section to 24 Hour ECG procedure specifying that if patients find the procedure too burdensome the 24 hour ECG at visit 4 can be made optional, as this is not included in any endpoints.<br>• Corrected CI's email address. |
| V9.0 | 15 JAN 2024 | SA4 | • Updated trial contacts sections<br>• Minor updates to abbreviations table<br>• Correction of the following typographical errors and inconsistencies with CIP V8.0<br>(1) The inclusion criteria 6 on page 17 and on page 44: On page 17 we have corrected 'and mean daytime DBP of >85 mm Hg' (greater than or **equal** to) for consistency |

| | | | |
|---|---|---|---|
| | | | (as correctly documented on page 43, Section Inclusion 6.1, inclusion criteria 6)<br>(2) The inclusion criteria in CIP V8.0 was updated to ABPM readings, but the sentence about office BPs in 2.6 Trial design was incorrect. The following sentence has been deleted and replaced with the current ABPM BP inclusion criteria: 'or mean office BP (which is mean of last two readings from 3 BP recordings) with recorded SBP of ≥140 mmHg and <180 mmHg and DBP ≥90 mmHg at screening or baseline (randomisation) visit (either one of the two visits or both)' remained on' .<br>(3) Correction in Section 1.1 Background (page 22) and Section 2.6 Trial Design (page 38) to confirm one to **four** oral antihypertensives.<br>• Clarified Exclusion criteria 11 to confirm clinically **significant or symptomatic** hypertension-mediated target organ damage<br>• Clarified Exclusion criteria 14 to confirm 'use of a **therapeutic** implantable electronic/electrical device'<br>• Clarified Exclusion criteria 15 to confirm 'Participant has history of hospitalization **(> 24 hour)** for heart failure, or cerebrovascular accidents, or **history of** stroke diagnosed **based on imaging or evidence of specialist diagnosis or any other indirect evidence such as discharge summary or clinical letter** (at any time in the past)'<br>• Clarification for Exclusion criteria 16 to state 'Participant has **mean daytime ABPM** pulse pressure ≥ 80 mmHg at screening or baseline (randomisation) visit'. This change reflects the previously approved amendment for inclusion criteria 6 (approved in CIP V8.0) from office to ABPM BP readings for eligibility.<br>• Update in Synopsis to change to planned enrolment months of **32 months** and planned duration to **40 months**, following sponsor recruitment suspension in September 2023 and required study extension.<br>• Update to Figure 1, Trial Flowchart (page 40) for the statement at the bottom to confirm ' All participants consented **and randomised** will be followed up by telephone …..on day 112'<br>• Update in Section 3.2 for Participant identification procedures and reference to self referral methods and updates for contact via |

| | | | |
|---|---|---|---|
| | | | nhs email address in posters, websites and social media advertising<br>• Update in Sections 3.2 and 4.1 for total participant reimbursement increased from £25 to £35, following feedback from participants.<br>• Clarification in Section 7.1 of use of Participant ID number (and not separate screening or randomisation numbers)<br>• Update in Section 7.4 for randomisation and unblinding procedures in Sealed Envelope<br>• Updates to sections 9.3 and 9.5 regarding the Barts Bioresource name and information provided in PIS<br>• Update to Device Manufacturer registered address for Afferent in Section 11.1<br>• Update in Section 11.9 regarding Instructions for Use (IFU) being Appendix B **of the IB**<br>• Further details added in section 13.7 regarding reporting of device deficiencies.<br>• Clarification in section 13.12 Pregnancy to confirm subject will be followed up until their end of **participation in the study**, and not end of trial<br>• Update to Section 15.3 for inclusion of PICs (Royal Free, Kings College) as new PICs added with CIP V8.0, and addition of new PIC (Broomfield Hospital)<br>• Update to Section 16.11 to include Health Economics Analysis being performed<br>• Update to section 17.2 regarding CRFs and data access<br>• Update to section 18 to clarify use of the single participant ID throughout the trial (and not separate screening or randomisation numbers)<br>• Update to Section 25.2 with removal of the requirement for an independent statistician to be on the TSC, as this membership is included in the DSMC, and this change has been approved by all trial committees and sponsor.<br>• Update to Section 26.2 to include the CVCTU and Oxford AHSN as data processors.<br>• Update to Section 26.3, with text regarding the data sharing plan, as updated on the ISRCTN registry.<br>• Change from Co-Investigator to Sub-Investigator, as per Sponsor SOP requirement.<br>• General typographical, grammatical and language changes, including page numbering. |
| 10.0 | 29OCT2024 | SA5 | • Clarification for Inclusion criteria 6 that the screening ABPM will be used for eligibility criteria (page 18 and page 45) |

| | | | |
|---|---|---|---|
| | | | • Clarification for Inclusion criteria 7c for use of rate-limiting calcium channel blocker medication and revised text 'or a heart rate (any one of the three recordings) ≥60 bpm at screening or baseline (randomisation) visit if the patient is taking beta-blocker medication or **a rate-limiting calcium channel-blocker medication.' (**page 18 and page 44**)**<br>• Clarification for Exclusion criteria 20 of 'hospitalisation' definition (page 20 and page 46)<br>• Deletion in Exploratory endpojnts (page 30 and table on page 37) of CBP measurement from baseline to 1 month, as this CBP measurement for the sub-study was never planned at Visit 4 (1Month) and is not mentioned in the study visits descriptions or visit schedule in the CIP or in the PIS for the sub-study (Only CBP at Visit 2 and Visit 5)<br>• Section 2.7 Trial Setting (page 41) – Addition of use of radio advertisements in point 5 (as approved with SA4/CIP V9.0) and addition of point 7 to include involvement with North Thames Clinical Research Network research promotion activities for trial recruitment and point 8 involvement with the NIHR Be Part Of Research Volunteer service (BPORVS).<br>• Clarification of repeat ABPM at baseline, only if medication changes since screening (page 47)<br>• Update to Schedule of Assessments 7.2 to confirm Visit 5 Holter and/or Echocardiogram is conducted within -5 to +14 days of the Visit 5 clinic date (page 54)<br>• Safety Bloods and Urine Assessments - Inclusion of urine pregnancy testing done **at randomisation** (page 59) for consistency with description of Visit 2/Day 0 (page 49) and Schedule of Assessments table 7.2 (page 53)<br>• Annual Progress Report – removal of section 14.1 as submission of APR to REC /HRA is no longer applicable (page 76)<br>• Section 16.4 Primary Endpoint analysis (page 78) - Change in t-test used for statistical analysis, following independent review of the Statistical Analysis Plan |

#### I. Contents

7.2 Schedule of Assessments ... 50
7.3 Randomisation Method ... 54
7.4 Randomisation, Blinding and Unblinding Procedure ... 54
7.5 Trial Assessments ... 55
7.6 Follow up Procedures ... 57
8.0 Participant, Study, and Site Discontinuation ... 58
9.0 Laboratories and Samples ... 58
9.1 Local Laboratories ... 58
9.2 Sample Collection, Labelling, and Logging ... 58
9.3 Sample Transfer, Chain of Custody, and Accountability ... 59
9.4 Sample Analysis Procedures ... 59
9.5 Sample Storage Procedures ... 60
9.6 Sample and Result Recording and Reporting ... 60
9.7 Sample Management at End of study ... 60
10.0 Investigational Device ... 61
10.1 Name and Description of Investigational Device ... 61
11.0 Legal Status of Investigational Device ... 62
11.1 Device Manufacturer (s) and supply arrangement ... 63
11.2 Investigational Device Software ... 63
11.3 Investigational Device Accessories ... 64
11.4 Packaging and labelling of Investigational Devices ... 64
11.5 Accountability and Traceability ... 65
11.6 Assessment of compliance ... 65
11.7 Device storage ... 65
11.8 Device training and experience requirements ... 66
11.9 Administration of investigational device ... 66
11.10 Destruction, return, and recall devices ... 66
11.11 Usage schedules ... 67
11.12 Usage modifications and delays ... 67
11.13 Management of device-specific adverse events ... 68
11.12 Potential interactions with other therapies ... 68
11.13 Recommended concurrent treatment ... 69
11.14 Prohibited therapies ... 69
11.15 Study restrictions ... 69
11.16 Management of incorrect usage ... 69
11.17 Precautions regarding contraception ... 69
11.18 Arrangements for post-study access to the investigational device ... 69
12.0 Equipment and other Devices ... 69
13.0 Safety Reporting ... 69

#### II. Glossary of Terms and Abbreviations

| | |
|---|---|
| 6MWT | Six Minute Walk Test |
| ABPM | Ambulatory Blood Pressure Monitoring |
| ABS | Acrylonitrile butadiene styrene |
| ACEI | Angiotensin-Converting Enzyme Inhibitors |
| ADE | Adverse Device Effect |
| AE | Adverse Event |
| AFT | Autonomic Function Test |
| APR | Annual Progress Report |
| AR | Adverse Reaction |
| ARBs | Angiotensin II receptor blockers |
| ATONT | Autonomic Target-organs Neurophysiological tests |
| BMI | Body Mass Index |
| BP | Blood Pressure |
| BPM | Beats Per Minute |
| BBR | Baroreflex Responsiveness |
| CAPA | Corrective and Preventive Action |
| CCBs | Calcium-Channel Blockers |
| CI | Chief Investigator |
| CIP | Clinical Investigational Plan |
| CKD | Chronic Kidney Disease |
| CRC | Clinical Research Centre |
| CSB | Cardiac Sensitivity to Baroreflex |
| CVCTU | Cardiovascular Clinical Trials Unit |
| CVD | Cardiovascular Disease |
| CVT | Cardiac Vagal Tone |
| DBP | Diastolic Blood Pressure |
| DD | Device Deficiency |
| ECG | Electrocardiogram |
| eCRF | Electronic Case Report form |
| eGFR | Estimated Glomerular Filtration Rate |
| EoA | Extent of Adherence |
| EQ5D | EuroQol 5 Dimension |
| EUDAMED | European Medical Devices Regulatory Database |
| FBC | Full Blood Count |
| GCP | Good Clinical Practice |
| GP | General Practitioner |
| HR | Heart Rate |
| HRA | Health Research Authority |
| IB | Investigator Brochure |
| ICH | International Conference on Harmonisation |
| ISI | Insomnia Severity Index |
| DSMC | Data Safety Monitoring Committee |
| IMD | Investigational Medical Device |
| ISF | Investigator Site File |
| JRMO | Joint Research Management Office |
| LAV | Left Atrial Volume |
| LPLV | Last Patient Last Visit |
| LVEDP | Left Ventricular End-Diastolic Pressure |
| LVEF | Left Ventricular Ejection Fraction |
| LVM | Left Ventricular Mass |
| LVS | Linear Vagal Scale |

| | |
|---|---|
| MACE | Major Adverse Cardiovascular Events |
| MAP | Mean Supine Arterial Blood Pressure |
| MDR | Medical Device Regulations |
| MHRA | Medicine and Healthcare Regulatory Authority |
| mm Hg | Millimetre of Mercury |
| mmo\L | Millimoles per litre |
| MRA | Mineralocorticoid receptor antagonists |
| NHS | National Health Service |
| NICE | The National Institute for Health and Care Excellence |
| NSAIDs | Non-Steroid-Anti-inflammatory Drugs |
| OTC | Over-The-Counter |
| PIC | Patient Identification Center |
| PID | Patient Identifiable Data |
| PIS | Participant Information sheet |
| PoC | Proof of Concept |
| PPI | Patient and Public Involvement |
| QA | Quality Assurance |
| QC | Quality Control |
| QMUL | Queen Mary University of London |
| QoL | Quality of Life |
| RSG | Research Steering Committee |
| RSI | Reference Safety Information |
| RMT | Relativeness Mass Thickness |
| SADE | Serious Adverse Device Effect |
| SAE | Serious Adverse Event |
| SAR | Serious Adverse Reaction |
| SBP | Systolic Blood Pressure |
| SD | Standard deviation |
| SDA | Source Data Agreement |
| SOP | Standard Operating Procedure |
| tAN | Trans-cutaneous Autonomic Neurostimulation |
| TMF | Trial Master File |
| TMG | Trial Management Group |
| TSC | Trial Steering Committee |
| UDS | Urinary Drug Screen |
| UK | United Kingdom |
| USADE | Unanticipated Serious Adverse Device Effect |
| VAS | Visual Analogue Scale |

#### III. Signature Page

**Chief Investigator Agreement**

The trial as detailed within this Clinical Investigational Plan will be conducted in accordance with the principles of Good Clinical Practice, the UK Policy Framework for Health and Social Care Research, the Declaration of Helsinki, and the current regulatory requirements, including the Medical Device Regulations 2002 and all subsequent amendments. I delegate responsibility for the statistical analysis and oversight to a qualified statistician (see declaration below).

**Chief Investigator name:** **Dr Ajay Gupta**

**Signature:** Ajay Gupta
Ajay Gupta (Oct 29, 2024 13:32 GMT)

**Date:** Oct 29, 2024

**Statistician's Agreement**

The statistical aspects of the clinical study as detailed in this clinical investigation plan are in accordance with the principles of Good Clinical Practice, the UK Policy Framework for Health and Social Care Research, the Declaration of Helsinki, and the current regulatory requirements, including the Medical Device Regulations 2002 and all subsequent amendments.

I take responsibility for ensuring the statistical work in this clinical investigation plan is accurate, and for the statistical analysis and oversight of this study.

**Statistician's name:** **Annastazia Learoyd**

**Signature:** 
Annastazia Learoyd (Oct 29, 2024 13:56 GMT)

**Date:** Oct 29, 2024

#### IV. Synopsis

| | |
|---|---|
| Full title | Sham controlled Randomized Control Trial evaluating the Safety, Acceptability and Efficacy of Autonomic neuromodulation using trans-cutaneous vagal sensory stimulation in uncontrolled hypertensive patients: a pilot study evaluating a novel non-invasive device-based strategy. |
| Short title and / or acronym | SCRATCH HTN |
| Sponsor | Queen Mary University of London |
| Medical device Risk classification | The Medical Device used in this investigation is classified as *Class IIa* |
| Phase of the trial | Pilot |
| Medical condition or disease under investigation | Systemic hypertension |
| Trial design and methodology | This is a pilot, sham-controlled, double blind, single-site device clinical trial designed to evaluate the safety, acceptability and efficacy of non-invasive autonomic neuromodulation in a cohort of 63 adult patients with uncontrolled high blood pressure. |
| Planned number of participants | The recruitment target is 63 patients with systemic hypertension. |
| Objectives | To determine whether treatment with tAN therapy can reduce daytime SBP in uncontrolled hypertensive subjects to a greater extent than treatment with a placebo sham therapy |
| Inclusion and exclusion criteria | Participants must meet all the following inclusion criteria to be eligible for the trial:<br>1) Participant has given written informed consent.<br>2) Participant has sufficient knowledge of the English language to be able to understand the participant information sheet and trial materials including outcome assessments.<br>3) Participant is aged ≥18 years and <80 years at the time of screening visit.<br>4) Participant is taking between 1 to 4 antihypertensive medications (inclusive) at time of screening and baseline (randomisation) visit and is willing to adhere to no change in medication during the trial until end of the trial visit (visit 5). (NB. Participant on only one antihypertensive medication should be taking that medication for at least six weeks prior to the screening visit).<br>5) Participant has confirmed diagnosis of hypertension. |

| | |
|---|---|
| | 6) Participant meets the following BP criteria:<br>24-hour ambulatory BP monitoring (ABPM) at either screening visit or baseline (randomisation) visit, with mean daytime SBP of ≥135 mmHg and <170 mmHg and mean daytime DBP of ≥85 mm Hg and <115 mmHg (N.B. By default, Ambulatory Blood Pressure Monitoring [ABPM] at screening visit will be used at baseline visit. However, if there has been an addition of new medication after participants screening visit, 24-hour ABPM must be repeated at baseline visit, but the screening ABPM will be used for eligibility criteria).<br>7) Participant has one or more of the following associated conditions:<br>a) Obesity: BMI >30 OR waist circumference >94 cm (men) or > 80cm (women). (NB. For participants of South-East Asian/Chinese/Japanese origin these cut-offs are >90 cm (men) or >80 cm (women)).<br>b) Type 2 diabetes – controlled or sub-optimally controlled (HbA1c ≤8.5% or ≤69 mmol/mol) on diet and/ or medications except insulin.<br>c) Heart rate (on any one of the three heart rate recordings at that visit) ≥70 bpm at screening or baseline (randomisation) visit (measurements taken after 5 minutes of rest in a seated position and when finger probe has been placed for a minimum of 30 seconds thereafter) or a heart rate (on any one of the three heart rate recordings) ≥60 bpm at screening or baseline (randomisation) visit if the patient is taking beta-blocker medication or a rate-limiting calcium channel-blocker medication.<br>d) HbA1c ≥42 mmol/mol or fasting blood glucose (if available) ≥5.6 mmo/L, and either low HDL cholesterol (≤1.03 mmol/L for men and ≤1.29 mmol/L for women) or high triglyceride (triglycerides ≥1.7 mmol/L)<br>e) Both low HDL cholesterol (≤1.03 mmol/L for men and ≤1.29 mmol/L for women) and high triglyceride (triglycerides ≥1.7 mmol/L)<br>f) Diagnosed or known case of polycystic ovarian syndrome.<br>8) Female participant of child-bearing potential (all those below 55 years except if they are surgically sterile, meaning they have undergone a hysterectomy, bilateral tubal ligation, or bilateral oophorectomy, or formally diagnosed by their doctors to be post-menopausal) must agree to use the acceptable methods of contraception from the time of consent until last follow up visit.<br>9) Participant is able to communicate satisfactorily with the Investigator and Investigation Site staff, and to participate in, and comply with all clinical study requirements.<br>10) Participant agrees to have all trial procedures performed and are able and willing to comply with all trial visits and protocol requirements. |

| | |
|---|---|
| | 1) If any of the following exclusion criteria is met, the participant cannot be randomised:Participant is unable and unwilling to use AffeX-CT device daily.<br>2) Participant has a small tragus (ie. the size or shape of the tragus is such that it doesn't allow the application of the ear-clips of the AffeX-CT device for a sustained period of time).<br>3) Participant has a piercing on the tragus of the ear.<br>4) Participant is diagnosed with atrial fibrillation or other form of cardiac arrhythmia.<br>5) Participant has eGFR <45 ml/min/1.73 $m^2$ at screening visit.<br>6) Participant has type 1 diabetes mellitus.<br>7) Participant has type 2 diabetes mellitus on Insulin or those on oral antidiabetic medications with poor glycaemic control defined as HbA1c above 8.5% (or >69 mmol/mol).<br>8) Participant has a history of falls or symptoms of orthostatic hypotension in the last 3 months prior to baseline (randomisation) visit.<br>9) Participant is pregnant, nursing or planning to become pregnant within the next 6 months.<br>10) Participant suffers from chronic pain and has taken anti-inflammatory drugs for two or more days per week over the last month prior to baseline (randomisation) visit.<br>11) Participant has clinically significant or symptomatic hypertension-mediated target organ damage such as severe heart failure with NYHA 4, end stage renal damage, medically diagnosed/imaging proven stroke, symptomatic peripheral vascular disease, or severe retinopathy.<br>12) Participant has a history of stable or unstable angina or had an acute coronary event within 3 months prior to baseline (randomisation) visit or had a myocardial infarction within the last six months of enrolment prior to baseline (randomisation) visit.<br>13) Participant has a history of renal denervation within last 1 year prior to baseline (randomisation) visit.<br>14) Participant has a therapeutic implantable electronic/electrical device such as pacemaker, implantable cardioverter-defibrillators (ICDs), implanted vagal stimulators.<br>15) Participant has history of hospitalization (> 24 hour) for heart failure, or cerebrovascular accidents, or history of stroke diagnosed based on imaging or evidence of specialist diagnosis or any other indirect evidence such as discharge summary or clinical letter (at any time in the past). |

| | |
|---|---|
| | 16) Participant has mean daytime ABPM pulse pressure ≥ 80 mmHg at screening or baseline (randomisation) visit.<br>17) Participant has a heart rate <50 bpm at screening or baseline (randomisation) visit (measurement taken after 5 minutes of rest in a seated position and when finger probe has been placed for a minimum of 30 seconds thereafter).<br>18) Participant has auricular dermatitis.<br>19) Participant has postural hypotension, defined as a fall > 20mmHg in SBP on standing at 3 minutes (compared with sitting).<br>20) Participant has a history of hospitalisation for hypertensive emergency or urgency in the last six months of enrolment prior to baseline (randomisation) visit. Hospitalisation' is defined as admission for more than 24 hours or between 12-24 hours with an overnight stay.<br>21) Participant is identified as unsuitable to participate by the CI/Sub-Investigator(s) and/or Investigation site team for another reason (e.g., for other medical reasons, laboratory abnormalities, limited life expectancy, etc.)<br>22) Participants with history of epilepsy and are currently on anti-epileptic medication or those who are not on any anti-epileptic medication but have history of a seizure within last 10 years. |
| Investigational device | Transcutaneous Autonomic Neuromodulation (tAN) device (Affex-CT) |
| Treatment duration | The total treatment duration is 12 weeks.<br>Self-administration of 30 min of tAN or sham stimulations once per day for the first two weeks, and then once every week for the rest of the trial period. |
| Follow up duration | Telephone follow-up at 16 weeks (4 weeks after the end of treatment/trial period of 12 weeks). |
| Total duration for participants | The total time that a participant will be involved in the investigation including screening, treatment and follow-up is 16 weeks. |
| Planned enrolment period | The anticipated time needed to recruit the required number of participants is 33 months. |
| Planned duration of Investigation | The total expected duration of the clinical investigation is 41 months. |
| End of Trial definition | The date of the last visit of the last participant recruited into the trial; last patient last visit (LPLV). |

### 1.0 Introduction

#### 1.1 Background

Hypertension or high blood pressure (BP) is the leading health risk factor globally [1, 2] and the leading cause of cardiovascular morbidity and mortality [3, 4]. Hypertension affects more than 1.5bn people worldwide [5]. In the UK >15m people in the 45-75 year age group are hypertensive, representing >60% of this age group [3]. Hypertension is the third biggest risk factor for premature death and disability, causing more than 75,000 deaths annually in England, with >1m disability-adjusted-life-years and estimated 179,857 years of lost life [3, 6]. Hypertension is estimated to cost the NHS in excess of £3.4bn annually.

Despite the widespread availability of antihypertensive drugs (the only regulatory approved intervention), 37% of all hypertensive patients fail to achieve the NICE recommended BP levels (<140mmHg systolic and <90mmHg diastolic) [7]. These patients are classified as *uncontrolled hypertensive patients*. Several factors contribute to sub-optimal BP control with ~14% (1.7m people in England) of all patients being drug-resistant [7, 8]. Non-adherence to prescribed medications is a major challenge, partly due to drug-intolerance/adverse side-effects (cough, fatigue, anxiety, dizziness, headaches). It is estimated that about 84% of uncontrolled and 45% of all hypertensive patients fail to adhere to the prescribed medications[9]. High BP markedly increases the risk of myocardial infraction, stroke, heart failure and renal damage [3, 4].

**Unmet clinical need:** For uncontrolled hypertensive patients, including drug-resistant patients, the lack of an effective therapy is a major health challenge and an urgent unmet clinical need.

**Autonomic neuromodulation as a treatment of high BP**: The brain controls the cardiovascular system by sending commands through specialised autonomic nerves – the sympathetic and parasympathetic. These are vital to produce rapid changes in heart rate and cardiac contractility and exert arteriovenous and cardiac circulatory control. During the development of hypertension, the activities of parasympathetic nerves decline, concomitantly with sympathetic activation, adversely and (if left untreated) permanently changing the fine homeostatic autonomic balance. One potentially highly effective strategy to improve BP control in hypertension is via redressing the autonomic imbalance. The current consensus of experts on the potential interventions that may be effective for the treatment of true drug-resistant hypertensive patients, or those who are intolerant to oral medications, are device-based solutions [10]. Of them, catheter-based renal denervation is the most advanced, and the data suggest that it can potentially provide a safe and efficacious solution [11, 12]. To date there have been 8 sham-controlled randomised trials that have all shown that the renal denervation leads to a significant reduction of daytime systolic and diastolic BP [11]. A recent study showed that the BP lowering effect of renal denervation is sustained over a period of six months [13]. Another study showed that, amongst uncontrolled hypertensive patients on treatment, the use of renal denervation (as compared to a sham procedure) leads to a 7.4 mmHg reduction in 24-h systolic BP, with no evidence of any safety concern [14]. All together these studies have established that amongst hypertensive patients, redressing the autonomic imbalance is an effective and safe approach. The meta-analysis of all renal denervation studies concluded that it leads on average to a 4 mmHg reduction in daytime BP [11]. However, this and other technologies currently under development are invasive, require hospitalisation and tertiary care delivered by experts, and are of high cost [12]. Moreover, whilst they may have sustained effect for six or more months[13], it is unknown how long this effect lasts for, and whether the repeat procedures are required (there is strong evidence obtained in large animal models of gradual re-innervation several months after the renal denervation).

Exploiting the potential of this therapeutic approach, Afferent Medical Solutions (‘Afferent’) developed a novel solution of hypertension treatment involving non-invasive autonomic neuromodulation achieved by transcutaneous electrical stimulation of auricular sensory innervation. This method is called transcutaneous autonomic neuromodulation – tAN. Afferent’s Proof-of-Concept (PoC) study used a specific tAN stimulation algorithm delivered in a course of treatment and demonstrated a significant (on average by 13 mmHg) reductions in 24-h SBP in the most challenging to treat cohort of drug-resistant hypertensive patients. The BP-lowering effect of tAN was found to last for several weeks after the discontinuation of therapy.

**SCRATCH-HTN trial** is a randomised sham-controlled study designed to evaluate the safety, acceptability, and efficacy of tAN in a cohort of uncontrolled medicated hypertensive patients. The study will recruit 63 patients with systemic arterial hypertension (male and female aged ≥18 years) who are receiving between one and four oral antihypertensive medications and remain hypertensive with BP above target levels detailed below. The participants will be randomly allocated to the active (tAN) or sham (sham-tAN) arms of the trial on 2:1 basis.

**Investigational product**: tAN treatment will be administered using AffeX-CT device (referred herein as “AffeX device” or “device”). Recent advances in device-based neuromodulation have made it possible to redress the autonomic balance by non-invasive (i.e. through the skin) stimulation of certain regions of the auricle (the tragus in particular) that receive innervation by the sensory branches of the fifth (V) and the tenth (X) cranial nerves as well as branches of the spinal nerves C2 and C3. There is significant evidence that low-current electrical stimulation of these sensory nerve projections acutely shifts the autonomic balance towards a beneficial net parasympathetic dominance, as evident from the reported changes in baroreflex sensitivity [15] and heart-rate variability [16]. Moreover, the individuals with greater baseline sympathetic drive, demonstrate more pronounced shifts towards parasympathetic prevalence with tAN [15].

AffeX device comprises of a battery-operated control unit, two electrode pairs arranged on ear clips and connected to the control unit with electrical leads. The AffeX device generates an electrical signal that is used to stimulate the sensory cutaneous innervation of the auricle.

**PoC data**: Afferent’s PoC study showed significant reductions in 24-h ambulatory BP following a course of tAN treatment in patients with drug-resistant hypertension (Figure 1a), also leading to a reduction in the number and doses of anti-hypertensive medications taken and in patients with uncontrolled arterial hypertension (Figure 1b). Drug-resistant hypertension was diagnosed in patients that displayed an office systolic BP of >150 mmHg and ambulatory systolic BP of ≥130 mmHg, despite adherence to maximally tolerated doses of at least three antihypertensive medications, including a diuretic. Uncontrolled hypertension was diagnosed in patients with elevated BP that were either untreated for high BP (i.e. not prescribed with any antihypertensive medications), or patients that displayed either an average office systolic BP between 130-180 mmHg and diastolic BP ≥ 80 mmHg, or daytime average systolic BP between 120-160 mmHg and daytime average diastolic BP of >80 mmHg, and prescribed with up to 3 antihypertensive agents. The BP-lowering effect of tAN was sustained for several weeks after discontinuation of therapy. These PoC study data suggest that the antihypertensive effect of tAN treatment can potentially exceed the BP lowering effect of catheter-based renal denervation therapy. No patient’s BP fell to the levels associated with hypotension.

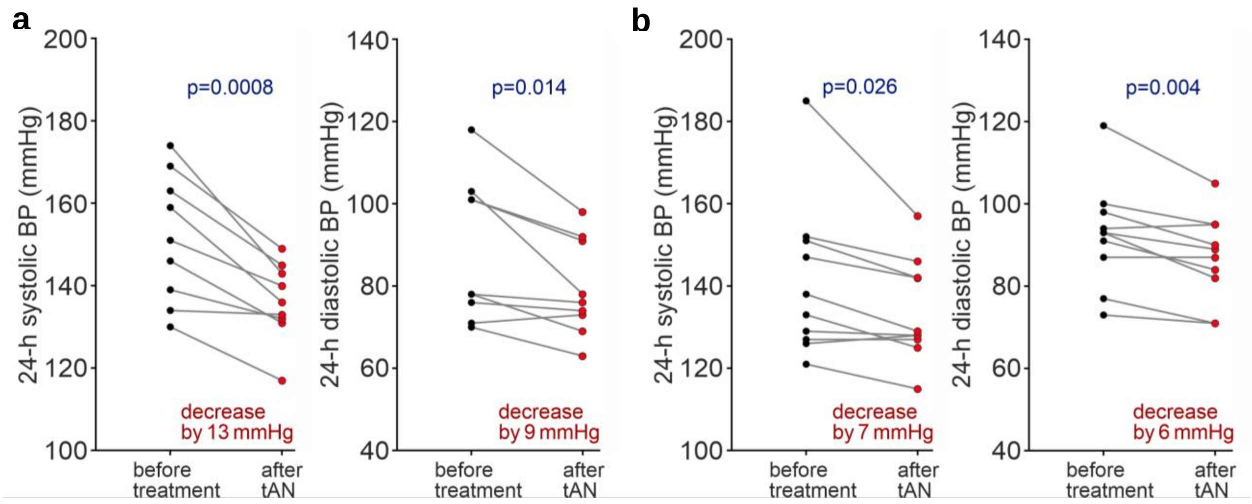

**Figure 1** | The effect of tAN on 24-h blood pressure in drug-resistant (n=9, **a**) and uncontrolled (n=10, **b**) patients with arterial hypertension. Data shown are before and one month after the course of *t*AN treatment.

**Safety**: Results of the systematic review and meta-analysis of all relevant studies published prior to August 2018 (51 studies including 1,322 participants) indicate that tAN is a safe and well-tolerated treatment [17]. In a study on healthy subjects (6 healthy volunteers) no adverse effects of tAN delivered using AffeX-CT device on heart rate, blood pressure or ECG parameters (QT interval) were observed during 30 min of stimulation and for 60 min after the stimulation. No side effects and/or serious adverse events were reported by the patients recruited in the PoC study (19 participants).

#### 1.2 Rationale for Trial Design

SCRATCH-HTN trial is designed to test the hypothesis that tAN treatment is safe and acceptable to the patient, improves the control of blood pressure in hypertension and sense of well-being amongst those who are receiving the active treatment as compared to those on sham treatment. The primary aim of this trial is to establish the safety and acceptability, and to generate additional data regarding the efficacy of this treatment strategy, from which to develop a larger efficacy and cost-effectiveness study.

In the UK, of the 9 million hypertensive patients receiving medication, more than 3 million fail to maintain their BP within the NICE recommended guidelines and, therefore, these patients are at a higher risk of adverse cardiovascular events. A significant proportion of these patients are drug-resistant or drug-intolerant. For these patients a device-based therapy is expected to provide an effective alternative solution. Large epidemiological studies have shown that in hypertension, a 10 mmHg reduction in BP reduces the risk of coronary heart disease (CHD), stroke, heart failure, renal failure and all-cause mortality by 17%, 27% 28%, 5% and 13% respectively [18]. The difficulty of controlling high BP in a significant proportion of patients, the high prevalence of hypertension and its severe consequences, the absence of novel antihypertensive drugs in the development and the limitations of pharmacological approaches have all motivated the development of device-based approaches aimed to provide complementary treatments via modulation of the autonomic nervous system activity.

Autonomic neuromodulation can be achieved by using devices that block or activate specific autonomic nerves directly or indirectly. Whilst several device-based interventions under development (including renal nerve denervation, carotid baroreceptor stimulation, carotid bulb expansion and carotid body ablation) showed some promise, all these solutions are invasive, expensive, require surgical procedures in tertiary health-care settings, significant clinical expertise and special equipment [11, 12, 19]. None of the device-based invasive approaches are currently recommended for routine treatment of arterial hypertension.

In comparison, tAN treatment applied using the AffeX device is non-invasive, safe, low cost, doesn't need special equipment and can potentially be self-administered by the patients, and, therefore, potentially scalable to the mass usage. Patient feedback from our PPI activities indicated that the proposed tAN-based therapy might be a preferred 'treatment of choice' by the hypertensive patients.

If the findings of the PoC study are confirmed (reduction in systolic BP by >10 mmHg, lasting for >1 month after the discontinuation of therapy), tAN will prove to be more efficacious, as compared to renal denervation procedure or taking any single anti-hypertensive medication. Just to contextualise these findings on the significance of lowering BP by 5-10 mmHg: a recent – the largest and most detailed – individual patient-level meta-analysis of data obtained in 348,854 participants from 48 randomised clinical trials (that evaluated the effects of BP -lowering treatments on the risk of major cardiovascular events and death in patients with and without cardiovascular disease) demonstrated that over an average of 4 years of follow-up, each 5 mmHg reduction in systolic BP lowered the relative risk of major cardiovascular events by ~10%. The risks of stroke, ischaemic heart disease, heart failure and death from cardiovascular disease (CVD) are reduced by 13%, 7%, 14% and 5%, respectively, with each 5 mmHg reduction in systolic BP [20].

Building on the promising results of the PoC study, SCRATCH-HTN main trial is designed as a randomised, double-blind, sham-controlled study. Results of several earlier trials that tested the efficacy of renal nerve ablation as a treatment of high BP were hampered by lack of sham treatment groups. Therefore, randomized-parallel arm, sham-controlled study design is the gold standard for clinical trials of this type. The trial will recruit 63 patients, to enable a 2:1 fractional allocation to active (n,42) - and sham (n,21) - treatment groups with 80% power at the two-sided alpha level of 0.05 (assuming SD of mean difference of 11 mmHg) to allow detection of a minimum difference of 8.9 mmHg between the treatment and sham-stimulation groups, after allowing for 20% drop-outs and compliance-related issues.

**SCRATCH-HTN Sub-Study**

tAN treatment applied using the AffeX device aims to redress the autonomic imbalance that some of the hypertensive patients exhibit. 26 participants recruited into the SCRATCH-HTN main trial will be invited to take part in the SCRATCH-HTN sub-study, where we will evaluate the autonomic function before, at the time or immediately after the randomisation and at the end of the trial period. This will provide us with the mechanistic and pathophysiological insights about which components of the autonomic functions are affected by the treatment. As part of this sub-study participants will complete a number of physiological tests, commonly known as Autonomic Target-organs Neurophysiological tests (ATONT). The results of all these tests provide quantitative indications of the statuses of the resting sympathetic and resting parasympathetic activities in an individual together with quantitative statuses of the central regulatory mechanisms and peripheral inputs together with central outputs of the controls of BP. It takes approximately 90 minutes to carry out all the procedures in ATONT. The procedure itself is simple, painless and non-invasive. It includes continuous measurements of heart rate, beat-to-beat blood pressure, tissue oxygenation, respiratory rate, and ECG monitoring. Participants will be asked to complete simple physiological manoeuvre including standing from lying position, hand-grip, sit up, Valsalva manoeuvre and carotid massage (for full procedure details see Appendix 1). The objective of ATONT is to collect information on the standardised clinical measures, which provides understanding about various aspects of autonomic function.

These include:

1. Resting supine central parasympathetic activity measured as cardiac vagal tone (CVT) and quantified using the validated clinical units of the atropine-derived Linear Vagal Scale (LVS).
2. Central respiratory modulation of CVT will be quantified to examine the central respiratory gating of parasympathetic activity. This test examines the brainstem regulatory inter-neurons controlling cardiorespiratory functions.
3. Central baroreflex function will be quantified as the overall central baroreflex gain, also known as central baroreflex responsiveness (BRR). Including the prevailing negative feedback control of the resting supine blood pressure, which is also known as Cardiac Sensitivity to Baroreflex (CSB). These are the main real-time central blood pressure restraining mechanisms at rest as well as during physical exercise.
4. Peripheral baroreflex function will be examined by quantifying the cardiodepressor and vasodepressor responses of the carotid sinus to digital massage. These are surrogate measures of peripheral inputs into the brainstem for real-time blood pressure regulation.
5. Resting supine central sympathetic activity will be measured using its best surrogate, the mean supine arterial blood pressure (MAP).
6. Regional sympathetic activities will be examined by quantifying the following:
    a. Cardioaccelerator function will be quantified by measuring the increase in heart rate during a standardised isometric exercise. This is an important surrogate of the autonomic control of the chronotropic function of the heart.
    b. Vasoconstrictor function in the skeletal muscles will be quantified by measuring the change in diastolic blood pressure during standardised isometric exercise. This is an important surrogate of the autonomic control of the windkessel vascular resistance.
    c. $a_1$-Adrenergic function in the splanchnic vascular bed will be quantified by measuring systolic blood pressure changes in the late Phase IIi of Valsalva's manoeuvres. This is an important surrogate of the autonomic control of the venous vascular capacitance.

**Cognitive Assessments**

Evidence suggests that hypertension and presence of uncontrolled blood pressures over a period of time is associated with progressive decline in cognitive function. Data also suggests that neuromodulation improves quality of life, mood and sleep [16, 21]. It is possible that improvement in autonomic imbalance will also be associated with increased cerebral blood flow, by improvement of blood pressures. Therefore, in this study cognitive assessment will be performed, as an exploratory outcome, to assess the effect on cognitive performance. Participants will be asked to complete a computer-based cognitive assessment test designed to be sensitive to minor cognitive impairment. The test battery will consist of a series of visuospatial recognition tasks administered using PEBL2 software [22]; this software has been used in over 700 peer-reviewed publications and extensively validated with human subjects. The test battery will be initiated by the input of a Participant ID number into the software and will proceed automatically with on-screen instructions until completion. Participants will interact with the test using single key-press inputs when prompted and will take no more than

10 minutes to complete. The same test sequence will be repeated during the post-treatment assessment. Test data will be stored locally in .csv format consisting of Participant ID number and test data with one file per participant. The outcome measure will be the difference in cognitive performance before the treatment course and that measured afterwards. Cognitive performance is assessed by combining test performance metrics, for example; number of correctly matched task elements and reaction time to test cues.

#### 1.3 Assessment and Management of Risk

This project aims to develop a novel, low-cost, self-administered, device-based solution for patients with high blood pressure who do not respond to medications, or are intolerant/unable to use them. The benefits of lowering BP in such patients is well-known, which will not only reduce the risk of stroke, heart failure, coronary artery disease and cardiovascular mortality, but also improve the quality of life and life-span of the patients.

**Risk and benefits related to inclusion in the trial:** The inclusion in trial is likely to be beneficial for all patients, whether they receive active or sham arm, not only due to Hawthorne effect but also because of the overall care that they will receive in being a part of the trial. Therefore, we foresee little additional risk. In addition, our device – as per pilot data and other similar device-based studies- is likely to be efficacious and that will further reduce their risk vs. if they were in usual care.

The trial participants included are those with uncontrolled BP on treatment, and are not asked to change their medications. That can be deemed as a risk. However, the trial duration is only for 12 weeks, which is a much shorter period than that in the community or secondary care after dose escalation or another appointment. Moreover, we will be carefully monitoring these patients during the trial period and if, in the unlikely case, their BPs are at a levels that can potentially damage target organs, for example, mean average office BPs : SBP > 180 mmHg or DBP> 120 mmHg, at any stage during the trial, we will allow introduction of another medication or escalation of existing antihypertensive treatment as per the current guidelines/ their physician advice. We will make a note of all such instances. We will also instruct all participants to routinely use home BP monitoring as they would normally do. However, we will advise them to always take 3 BP readings – when they do their BP monitoring- and record the lowest of the three readings. Participants will be advised to contact the study team if their. systolic BP ≥ 170 mm Hg or diastolic BP≥ 115) . Aside from that, within the first month of the randomization the study team will have at least two opportunities to monitor their office BPs (at 2 weeks and 4 weeks after randomization), as well as conduct a 24 hour ABPM (at 4 weeks). If the study team notes any significant increase in BP (although below the target for escalation) or rising BPs trends, the study team will pro-actively advise participants to take BP recordings more often, when at home. The study team, on a case-by-case basis and depending on clinical judgement of investigators may also arrange additional clinical interactions to closely monitor BPs, and act on them as and when needed. This will allow for earlier escalation in the treatment, if required. It is possible, but unlikely that participants have significant lowering of BPs such that they develop postural hypotension, or symptoms of that. This is extremely unlikely given that included participants have uncontrolled BPs, but we will be monitoring all patients carefully and if the office (or where available home) SBP<100 mmHg systolic or they have postural hypotension, then we will allow reduction in their existing medications or stopping them altogether following standard treatment guidelines together with temporarily stopping any self-stimulation until the participant is clinically assessed by investigator/physician. The physician will evaluate the patient as soon as possible within 3 working days, and depending on the BP recordings on the day of the evaluation and the home BP recordings prior to that day, will decide whether to stop/reduce the oral medications alone or stop using the device as well. If in this evaluation, BPs is high, then antihypertensive medication in reduced dosage with or without device treatment will be recommenced in gradual manner under monitoring. However, if in this clinical assessment SBP remains lower than 100 mm Hg, or person is still symptomatic with postural hypotension, then antihypertensive agents and device stimulation will remain suspended, and those participants will again be assessed within 7-14 days, and a decision will be taken on medication and device use depending on their office and home BP measurements. All such participants will be encouraged to document their home BPs during that period. If in the subsequent evaluation, BPs are high or within reasonable limits, device stimulation may continue alone with no other medication treatment and participants will be closely monitored as per their clinical need. However, if BPs remain low in those evaluation, no treatment will be offered but participants will remain under monitoring for full study period.

No serious or life-threatening risks related to the use of this device are known; there is a limited data available on this, but most available data suggest that this device is safe and with only have transient and mild/moderate adverse effects. In a study on healthy subjects (6 healthy volunteers), no adverse effects of tAN administered using AffeX-CT device on heart rate, blood pressure or ECG parameters (QT interval) were observed during 30 min of stimulation and for 60 min after the stimulation. No side effects and/or adverse events were reported by the patients recruited in the proof-of-concept study (19 participants).

Results of the systematic review and meta-analysis of all relevant studies published prior to 2018 indicate that tAN is a very safe and well-tolerated treatment (51 studies including 1,322 participants). The most common side effects were local skin irritation from electrode placement (240 participants, 18.2%), headache (47, 3.6%) and nasopharyngitis (23, 1.7%). Whilst heterogeneity in overall side-effects event rates between studies was not accounted for by the parameters of stimulation, a minority of subjects (35 participants, 2.6%) dropped out of studies due to the side-effects. It is important to note that 89 studies were not included in that analysis as these studies had not reported safety or tolerability data and when approached the authors did not respond to formal requests to provide data.

The following side effects with frequency of <1% associated with tAN applied via electrical stimulation of the tragus may be anticipated:

- Light-headedness
- Fatigue/tiredness
- Mood changes
- Neck pain
- Tooth pain
- Pain/local skin irritation due to attachment of the device ear clips
- Tingling sensation due to the use of the device
- Ventricular extrasystoles (increased frequency)

Most of these adverse side effects are transient and temporary in nature, and are likely to reduce in intensity and frequency over regular usage.

### 2.0 Trial Objectives

#### 2.1 Primary Objective

To determine whether treatment with tAN therapy can reduce daytime SBP in uncontrolled hypertensive subjects to a greater extent than treatment with a placebo sham therapy.

#### 2.2 Secondary Objective (s)

- To determine whether treatment with tAN therapy can reduce daytime DBP to a greater extent than treatment with a sham therapy.
- To determine whether treatment with tAN therapy can reduce 24-hour SBP and DBP to a greater extent than treatment with a sham therapy.
- To determine whether treatment with tAN therapy can lead to a higher proportion of patients with controlled BP than treatment with a placebo sham therapy.
- To evaluate differences in BP variability between those receiving tAN therapy compared to those receiving sham therapy.
- To evaluate differences in HR variability between those receiving tAN therapy compared to those receiving sham therapy.
- To evaluate differences in reported serious adverse events (SAEs) and major adverse cardiovascular events (MACE) between those receiving tAN therapy compared to those receiving sham therapy.
- To determine whether treatment with tAN therapy can improve quality of life and well-being to a greater extent than treatment with a placebo sham therapy.
- To determine whether treatment with tAN therapy can improve quality of sleep to a greater extent than treatment with a placebo sham therapy.
- To evaluate differences in cumulative adherence to medications between those allocated to treatment with tAN therapy and those on sham therapy.

##### 2.2.1 Exploratory Objective (s)

- To evaluate changes in left ventricular systolic and diastolic functions using echocardiography in participants receiving treatment with tAN therapy.
- To evaluate changes in anti-hypertensive medication between baseline and the end of follow-up in participants receiving treatment with tAN therapy.
- To compare the average daily number of antihypertensive medications that patients are on during the trial between those allocated to treatment with tAN therapy and those on a sham therapy.
- To evaluate changes in quality of life and well-being between baseline and the end of follow-up in participants receiving treatment with tAN therapy.
- To determine whether treatment with tAN therapy can reduce central BP to a greater extent than treatment with a placebo sham therapy.
- To evaluate whether treatment with tAN therapy compared with sham therapy is associated with improvement in cognitive functions including short-term memory and reaction/response time.

- To evaluate differences in brainstem regulatory inter-neurons controlling cardiorespiratory function between those receiving tAN therapy compared to those receiving sham therapy.
- To evaluate differences in central baroreflex function between those receiving tAN therapy compared to those receiving sham therapy.
- To evaluate differences in prevailing negative feedback control of the resting supine blood pressure (cardiac sensitivity to baroreflex) between those receiving tAN therapy compared to those receiving sham therapy.
- To evaluate differences in peripheral baroreflex function between those receiving tAN therapy compared to those receiving sham therapy.

##### 2.2.2 Feasibility Objectives (s)

- To evaluate tolerability of the AffeX-CT device by patients.
- To evaluate adherence to use of the AffeX-CT device by patients.
- To evaluate the ease of use of the AffeX-CT device by patients.
- To determine reasons for early withdrawal from the study, if any.
- To evaluate the success of the blind procedure using a sham therapy control.

#### 2.3 Endpoints

##### 2.3.1 Primary Endpoint

Change in average daytime ambulatory SBP from baseline to the end of treatment (3 months).

##### 2.3.2 Secondary Endpoint (s)

- Change in average daytime ambulatory SBP and DBP from baseline and 1 month.
- Change in average daytime ambulatory DBP from baseline to the end of treatment (3 months).
- Controlled BP at the end of treatment (3 months) defined as mean daytime ambulatory SBP<135 mmHg and mean daytime ambulatory DBP<85 mmHg.
- Change in average 24-hour ambulatory SBP and DBP from baseline to the end of treatment (3 months).
- Change in average office SBP and DBP from baseline to 1 month, and from baseline to the end of treatment (3 months).
- Change in average daytime ambulatory HR, and in average night-time ambulatory HR from baseline to the end of treatment (3 months).
- Change in BP variability defined as the coefficient of variation (SD/mean) of 24-hour ambulatory SBP, and of within-visit office SBP from baseline to the end of treatment (3 months).

- Change in HR variability defined as the coefficient of variation (SD/mean) of 24-hour ambulatory HR, and of within-visit office HR from baseline to the end of treatment (3 months).
- Occurrence of a serious adverse event (SAE), fatal or non-fatal, within 3 months.
- The occurrence of a major cardiovascular event (MACE), including myocardial infarction (MI), stroke, and cardiovascular-related mortality within 3 months.
- Change in Quality of life between baseline and the end of the treatment (3 months) using the EuroQol Visual Analogue score (0-100), and the EuroQol 5 Dimension (EQ5D) quality of life (QoL) questions.
- Change in sleep quality between baseline and the end of the treatment (3 months) using the insomnia severity index (ISI), a 7-item questionnaire with each question allowing responses on a 5-point Likert scale from 0-4. Responses to the 7 questions can be summed to give an overall score of 0 to 28.
- Adherence to trial therapy, assessed as the proportion of days out of total days in follow-up when therapy was self-administered, and the average daily duration of self-administered therapy over the 3 months of follow-up (90 days).

#### 2.4 Exploratory Endpoint (s)

- Change in left ventricular ejection fraction (LVEF), left ventricular mass (LVM), relative mass thickness (RMT), left atrial volume (LAV), left ventricular end-diastolic pressure (LVEDP), and E/e´ ratio from baseline to the end of treatment (3 months), in participants receiving treatment with tAN therapy.
- Changes in anti-hypertensive medication between baseline and the end of follow-up (4 months) in participants receiving treatment with tAN therapy.
- Average daily number of antihypertensive medications that participants are on assessed through urinary drug screening (UDS).
- Change in Quality of life between baseline and the end of follow-up (4 months) in a sub-group of participants receiving treatment with tAN therapy**,** using the EuroQol Visual Analogue score (0-100), and the EuroQol 5 Dimension (EQ5D) quality of life questions.
- Change in average central BP, measured by Sphygmocor Vx device, from baseline to the end of treatment (3 months) – only for sub study participants.
- Change in the cognitive functions from baseline to the end of treatment (3 months)
- Central respiratory modulation of cardiac vagal tone.
- Overall baroreflex gain (central baroreflex responsiveness).
- Cardiodepressor and vasodepressor responses of the carotid sinus to digital massage.

##### 2.4.1 Feasibility Endpoint (s)

- AffeX-CT device-related adverse events within 3 months.
- Adherence to use of the AffeX-CT device assessed using the Extent of Adherence (EoA) questionnaire [23]
- Ease of use of AffeX-CT device assessed using a participant feedback visual analogue scale (VAS).
- Reasons behind any early withdrawals from the study.
- Success of the blinding procedure assessed using a blinding index at 1 month and 3 months [24]

#### 2.5 Objectives, Endpoint (s) and Outcome Measure Table

##### 2.5.1 Trial Efficacy and Safety

| Primary Objective | Primary Endpoint | Outcome measure |
|---|---|---|
| **To determine whether treatment with tAN therapy can reduce daytime SBP in uncontrolled hypertensive subjects to a greater extent than treatment with a placebo sham therapy by the end of treatment (3 months).** | Change in mean ambulatory **daytime SBP** from baseline to the **end of treatment (3 months)**. Daytime will be taken based on participant reported daytime hours, but where daytime hours have not been given, daytime hours will be defined as from 7am up to 11pm (16-hour period). | The difference in mean change in daytime ambulatory SBP, from baseline to the end of treatment (3 months) between active and sham therapy trial arms. |

| Secondary Objective | Secondary Endpoint(s) | Outcome measure(s) |
|---|---|---|
| **To determine whether treatment with tAN therapy can reduce daytime SBP in uncontrolled hypertensive subjects to a greater extent than treatment with a placebo sham therapy by 1 month.** | Change in mean ambulatory **daytime SBP** from baseline to **1 month**. Daytime will be taken based on participant reported daytime hours, but where daytime hours have not been given, daytime hours will be defined as from 7am up to 11pm (16-hour period). | The difference in mean change in daytime ambulatory SBP, from baseline to 1 month between active and sham therapy trial arms. |

| | | |
|---|---|---|
| **To determine whether treatment with tAN therapy can reduce daytime DBP in uncontrolled hypertensive subjects to a greater extent than treatment with a placebo sham therapy.** | Change in mean ambulatory **daytime DBP** from baseline to the **end of treatment (3 months).** Daytime will be taken based on participant reported daytime hours, but where daytime hours have not been given, daytime hours will be defined as from 7am up to 11pm (16-hour period). | The difference in mean change in daytime ambulatory DBP, from baseline to the end of treatment (3 months) between active and sham therapy trial arms. |
| | Change in mean ambulatory **daytime ambulatory DBP** from baseline to **1 month**. Daytime will be taken based on participant reported daytime hours, but where daytime hours have not been given, daytime hours will be defined as from 7am up to 11pm (16-hour period). | The difference in mean change in daytime ambulatory DBP, from baseline to 1 month between active and sham therapy trial arms. |
| **To determine whether treatment with tAN therapy can lead to a higher proportion of participants with controlled BP than treatment with a placebo sham therapy.** | Controlled BP defined as mean daytime ambulatory SBP<135 mmHg and mean daytime ambulatory DBP<85 mmHg at the **end of treatment (3 months).** Daytime will be taken based on participant reported daytime hours, but where daytime hours have not been given, daytime hours will be defined as from 7am up to 11pm (16-hour period). | The relative difference in proportion of participants with controlled BP at the end of treatment (3 months) between active and sham therapy trial arms. |
| **To determine whether treatment with tAN therapy can reduce 24-hour BP in uncontrolled hypertensive subjects to a greater extent than treatment with a placebo sham therapy.** | Change in mean ambulatory **24-hour SBP** from baseline to the **end of treatment (3 months).** | The difference in mean change in 24-hour mean ambulatory SBP, from baseline to the end of treatment (3 months) between active and sham therapy trial arms. |
| | Change in mean ambulatory **24-hour DBP** from baseline to the **end of treatment (3 months).** | The difference in mean change in 24-hour mean ambulatory DBP, from baseline to the end of treatment (3 months) between active and sham therapy trial arms. |

| | | |
|---|---|---|
| **To determine whether treatment with tAN therapy can reduce office BP in uncontrolled hypertensive subjects to a greater extent than treatment with a placebo sham therapy.** | Change in mean **office SBP** from baseline to the **end of treatment (3 months).*** | The difference in mean change in office SBP, from baseline to the end of treatment (3 months) between active and sham therapy trial arms. |
| | Change in mean **office SBP** from baseline to **1 month.*** | The difference in mean change in office SBP, from baseline to 1 month between active and sham therapy trial arms. |
| | Change in mean **office DBP** from baseline to the **end of treatment (3 months).*** | The difference in mean change in office DBP, from baseline to the end of treatment (3 months) between active and sham therapy trial arms. |
| | Change in mean **office DBP** from baseline to **1 month.*** <br><br> * Mean office BP will be calculated as the mean of the last 2 of 3 measurements taken. If only 2 are taken then the mean of those will be used, and if only 1 is taken that single measurement will be used. | The difference in mean change in office DBP, from baseline to 1 month between active and sham therapy trial arms. |
| **To determine whether treatment with tAN therapy can reduce heart rate in uncontrolled hypertensive subjects to a greater extent than treatment with a placebo sham therapy.** | Mean change in **daytime** ambulatory heart rate from baseline to the end of treatment (3 months). Daytime will be taken based on participant reported daytime hours, but where daytime hours have not been given, daytime hours will be defined as from 7am up to 11pm (16-hour period). | The difference in mean change in daytime ambulatory heart rate from baseline to the end of treatment (3 months) between active and sham trial therapy arms. |
| | Mean change in **night-time** ambulatory heart rate from baseline to the end of treatment (3 months). Night-time will be taken based on participant reported night-time hours, but where night-time hours have not been given, night-time hours will be defined as from 11pm up to 7am (8-hour period). | The difference in mean change in night-time ambulatory heart rate from baseline to the end of treatment (3 months) between active and sham trial therapy arms. |

| | | |
|---|---|---|
| **To evaluate differences in BP variability between those receiving tAN therapy compared to those receiving sham therapy.** | Mean change in **BP variability** defined as the coefficient of variation (SD/mean) of 24-hour ambulatory SBP, and also of within-visit office SBP from baseline to the **end of treatment (3 months).** | The difference in mean change in BP variability from baseline to the end of treatment between active and sham trial therapy arms. |
| **To evaluate differences in HR variability between those receiving tAN therapy compared to those receiving sham therapy.** | Mean change in **HR variability** defined as the coefficient of variation (SD/mean) of 24-hour ambulatory HR, and also of within-visit office HR from baseline to the **end of treatment (3 months).** | The difference in mean change in HR variability from baseline to the end of treatment between active and sham trial therapy arms. |
| **To evaluate differences in reported serious adverse events (SAEs) and major adverse cardiovascular events (MACE) between those receiving tAN therapy compared to those receiving sham therapy.** | The occurrence of SAEs (both fatal and non-fatal) within the 3 months of follow-up. | The difference in proportion of subjects experiencing an SAE within the 3 months of follow-up between active and sham therapy trial arms. |
| | The occurrence of a major adverse cardiovascular event (MACE), including myocardial infarction (MI), stroke, and cardiovascular-related mortality within 3 months of follow-up. | The difference in proportion of subjects experiencing a major cardiovascular event (MACE) within 3 months of follow-up between active and sham therapy trial arms. |
| **To determine whether treatment with tAN therapy can improve quality of life and well-being in uncontrolled hypertensive subjects to a greater extent than treatment** | Mean change in EuroQol Visual Analogue score (0-100) between baseline and the end of treatment (3 months). | The difference in mean change in EuroQol Visual Analogue score from baseline to the end of treatment (3 months) between active and sham therapy trial arms. |
| | Each of the EuroQol 5 Dimension (EQ5D) quality of life questions (each 1-5) at baseline and the end of treatment (3 months). | Descriptive visualisation of mean level at baseline and the end of treatment (3 months) for each quality of life dimension |

| | | |
|---|---|---|
| **with a placebo sham therapy.** | | presented separately for active and sham therapy trial arms. |
| **To determine whether treatment with tAN therapy can improve quality of sleep in uncontrolled hypertensive subjects to a greater extent than treatment with a placebo sham therapy.** | Mean change in the overall summative score of the 7 ISI question scores (overall summative score ranging from 0-28) | The difference in mean change in ISI summative score from baseline to the end of treatment (3 months) between active and sham therapy trial arms. |
| | Each of the 7 ISI questions (each 0-4) at baseline and the end of treatment (3 months). | Descriptive visualisation of mean level at baseline and the end of treatment (3 months) for each ISI question presented separately for active and sham therapy trial arms. |
| **To compare adherence between treatment with tAN therapy in uncontrolled hypertensive subjects to treatment with a placebo sham therapy.** | Proportion of days out of total days in follow-up when therapy was self-administered. | Difference in mean number of days when therapy was self-administered between active and sham therapy trial arms. |
| | Average daily duration of self-administered therapy. | Difference in mean daily duration of self-administered therapy between active and sham therapy trial arms. |

| **Exploratory Objective** | **Exploratory Endpoint** | **Outcome measure** |
|---|---|---|
| **To evaluate changes in left ventricular systolic and diastolic functions using echocardiography in a sub-group of participants receiving treatment with tAN therapy.** | Change in left ventricular ejection fraction (LVEF), left ventricular mass (LVM), relative mass thickness (RMT), left atrial volume (LAV), left ventricular end-diastolic pressure (LVEDP), and E/e´ ratio from baseline to the end of treatment (3 months). | Difference in mean change for each measure between active and sham therapy trial arms. |
| **To evaluate changes in anti-hypertensive medication between baseline and the end of** | Anti-hypertensive medication on at baseline and at the end of follow-up (4 months). | Descriptive tabulations of changes in anti-hypertensive medication |

| | | |
|---|---|---|
| **follow-up telephone consultation (4 months) in a sub-group of participants receiving treatment with tAN therapy.** | | from baseline to the end of follow-up (4 months). |
| **To evaluate differences in the daily average number of antihypertensive medications that participants are on between those receiving tAN therapy compared to those receiving sham therapy.** | Evidence of antihypertensive medication use through urinary drug screening (UDS). | The number and classes of drugs for which there is evidence that participants are found to be taking through urinary drug screening (UDS). |
| **To evaluate changes in quality of life and well-being between baseline and the end of follow-up telephone consultation (4 months) in a sub-group of participants receiving treatment with tAN therapy.** | Mean change in EuroQol Visual Analogue score (0-100) between baseline and the end of follow-up (4 months). | Mean change in EuroQol Visual Analogue score from baseline to the end of follow-up (4 months). |
| | Each of the EuroQol 5 Dimension (EQ5D) quality of life questions (each 1-5) at baseline and the end of follow-up (4 months). | Descriptive visualisation of mean level at baseline and the end of follow-up (4 months) for each quality of life dimension. |
| **To determine whether treatment with tAN therapy can reduce central BP in uncontrolled hypertensive subjects to a greater extent than treatment with a placebo sham therapy.** | **Mean change in central BP, measured by Sphygmocor Vx device, from baseline to the end of treatment (3 months).** | **The difference in mean change in central BP from baseline to the end of treatment (3 months) between active and sham trial therapy arms.** |

##### 2.5.2 Trial Feasibility

| Feasibility Objective | Feasibility Endpoint | Outcome measure |
|---|---|---|
| **To evaluate tolerability of the AffeX-CT device by participants.** | Device-related adverse events within 3 months. | Descriptive tabulations of device-related adverse events. |
| **To evaluate adherence to use of the AffeX-CT device by participants.** | Participant responses at the end of treatment (3 months) to the Extent of Adherence (EoA) questionnaire [23]. (The EoA questionnaire consists of 3 effect indicator | The number and percentage of patients responding at each of the 5 levels for each question will be tabulated descriptively. |

| | | |
|---|---|---|
| | questions to determine the extent of non-adherence, and 18 causal indicator questions to determine the reasons for non-adherence. | |
| **To evaluate the ease of use of the AffeX-CT device by participants.** | Participant feedback at the end of treatment (3 months) on the ease of use of the device, using visual analogue scale (VAS). | Descriptive tabulations of mean (SD) VAS for each ease-of-use question (range is from 1-10), as well as the overall sum of the scores across all VAS questions. |
| **To determine reasons for early withdrawal from the study, if any.** | Early withdrawal from the study, along with reason. | Number of participants who withdrawal early from the study, along with reasons will be tabulated. |
| **To evaluate the success of the blinding procedure using a sham therapy control.** | Assessment of success of blinding procedure using a blinding index assessed at 1 month and 3 months [24]. The blinding index is scaled to an interval of -1 to 1 with 1 being complete lack of blinding, 0 being perfect blinding and -1 indicating opposite guessing. | Blinding index as assessed at each time-point (1 month and 3 months) calculated for each trial arm. |

#### 2.6 Trial Design

The main trial is a double-blind, sham-controlled study, with a block randomised on a 2:1 basis where participants will receive either tAN treatment or sham- AffeX-CT device stimulation. The trial will recruit 63 participants with systemic arterial hypertension, who are receiving between one and four oral antihypertensive medications and remain hypertensive with BP above target levels: 24-hour ambulatory BP monitoring (ABPM) at either screening visit or baseline (randomisation) visit, with mean daytime SBP of ≥135 mmHg and <170 mmHg and mean daytime DBP of ≥85 mm Hg and <115 mmHg (N.B. By default, Ambulatory Blood Pressure Monitoring [ABPM] at screening visit will be used at baseline visit. However, if there has been an addition of new medication after participant's screening visit, 24-hour ABPM must be repeated at baseline visit.

Participants will be asked to self-administer tAN therapy using AffeX-CT device for 14 days (after Day 0) each day for the duration of 30 minutes in the late evening, preferably between 18.00-22.00. After 14 days, the participants will be asked to self-administer tAN therapy once a week for 10 weeks- i.e until end of treatment period of 12 weeks. Post-trial, we will follow these patients for safety assessment for further 4 weeks.

The expected duration of participation is 16 weeks (+/- 3 days).

Participants will be required to attend five visits, the sequence is as follows: visit 1 (screening), visit 2 (baseline and randomisation - Day 0), visit 3 at Day 14, visit 4 at Day 28 and visit 5 (End of Treatment - Day 84). Participants will also receive telephone conversations; one phone call between days 1-3, one at Day 7, and one at Day 56 and Day 112 (post trial follow up). Participants will also receive a text and/or email reminders at Day 42 and Day 70, respectively.

**Figure 1: Trial flowchart**

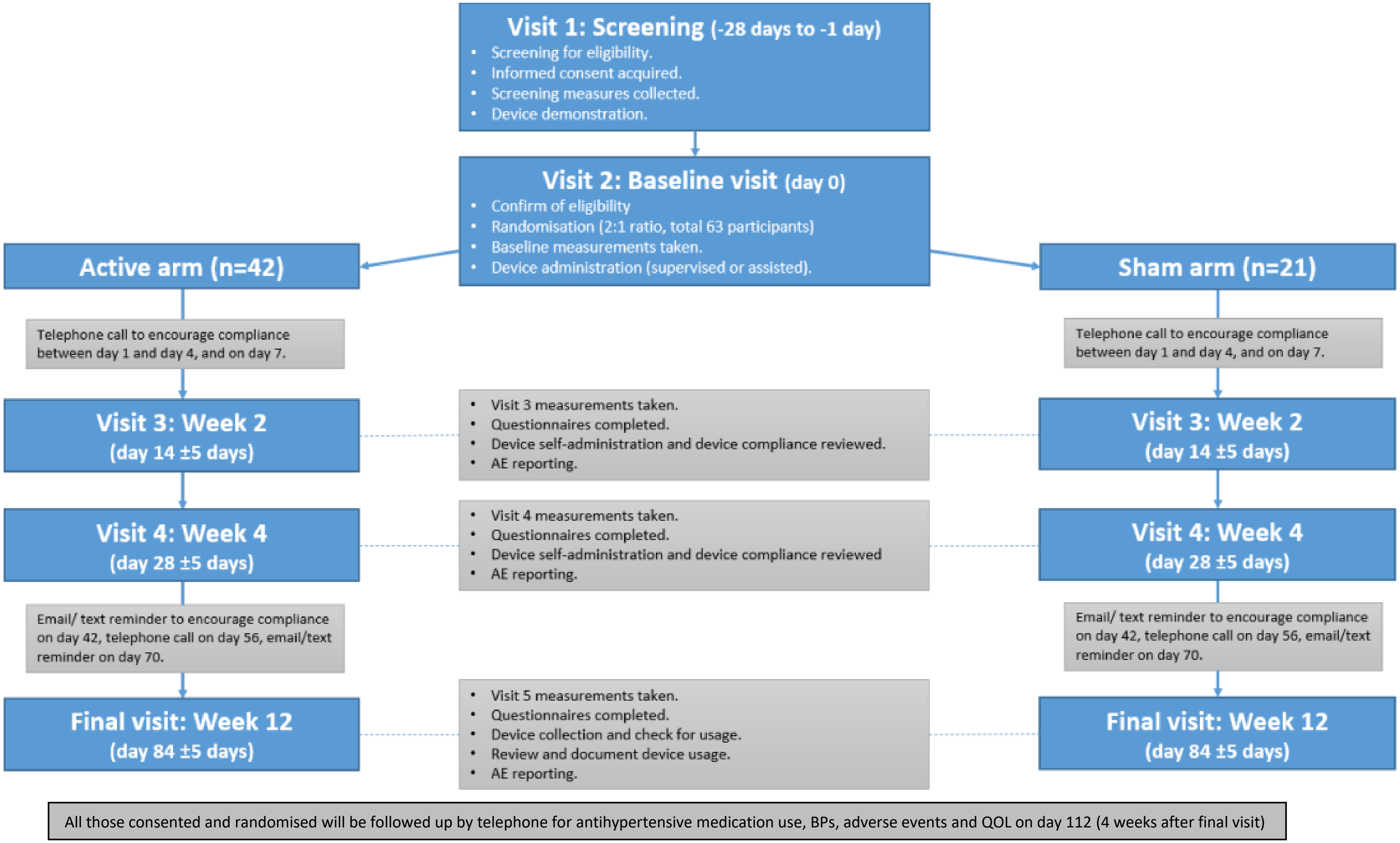

Participants eligible for the trial will receive AffeX-CT device (along with the User Guide), device logbook including training at visit 2. Participants completing device logbook (logging of the date, time and duration of each treatment session) will monitor compliance with the tAN procedure and urinary antihypertensive drug screening and Extent of Adherence (EoA) questionnaire will assess antihypertensive medications compliance.

No changes in the antihypertensive medications are permitted during the trial (between Day 0 and Day 84).

The sub-study is open label and 26 participants randomised to the main trial will be invited to take part. Participants will be asked to complete ATONT at visit 2 (baseline and randomisation - Day 0) and visit 5 (End of Treatment).

##### 2.7 Trial Setting

This is a single site trial. Participant recruitment and all trial activities will take place within an NHS setting at the William Harvey Clinical Research Centre (CRC) referred to as the Investigation site. Participants will be recruited from both primary and secondary care.

Participants will be referred for consideration of the trial from within the Barts Health NHS Trust or community in the surrounding attachment areas of Investigation site.

The following recruitment strategies may be used:

1. Specialist clinics at Barts Health NHS Trust sites
2. Internal site participant database
3. Email/ electronic information leaflet will be sent to Barts/QMUL staff to inform them about the study.
4. PIC sites in secondary and primary care (refer to section 15).
5. Social media recruitment: REC approved posters and social media campaigns will be launched. Posters will be placed in the specialists' clinics at approved sites, as a reminder to the clinicians. Social media recruitment, such as Facebook and Twitter advertisements, and newspaper and radio advertisement will be used for the public.
6. Direct referral by a treating clinician
7. Collaborating with NIHR-funded North Thames Clinical Research Network to participate in their events and research initiatives targeting more community research involvement and recruitment.
8. Involvement with the NIHR Be Part Of Research (BPOR) Volunteer service (see Appendix 2 for further details)

#### 3.0 Participants Evaluability and Replacement

##### 3.1 Target Accrual

The trial aims to recruit 63 participants with systemic arterial hypertension.

##### 3.2 Participant identification and recruitment

Participants will be identified from the hypertension care pathway by healthcare professionals at the Investigation site and/or at PICs during routine care visits, and will also be recruited with the use of publicity e.g., poster, radio advertisement, newspaper advert and/or social media platform (Facebook and/or Twitter). Participants who hear/see the trial advertisement will make themselves known to the research site via self-referral using an nhs.net email account, and they will provide their own contact details. This nhs.net email account will also be used on all publicity documents.

Recruitment will also include participants who have responded to outreach by the Investigation site team via trial recruitment strategies' including participants who are taking part in existing trials and/or previous trials and have previously consented to being contacted by the Investigation site.

The participant information received from participant self-referral and NHS referral will be maintained in an excel spreadsheet for the purposes of tracking pre-screening data, participant eligibility and study progress. This spreadsheet will be stored on a secure server, with restricted access to the CI and delegated research fellows (as authorised on the delegation log) and it will be password protected.

Following identification (and where appropriate) participants will be given a Participant Information sheet (PIS) that describes the trial and contains sufficient information for the prospective participant to make an informed decision about participating in the trial, at least 24 hours prior to signing a consent form.

Eligible participants will be offered the opportunity to participate in the trial, and discussions about the trial will take place and consent will be obtained prior to any trial related activities taking place. They will be screened and assessed by the Chief Investigator (CI)or sub-investigator, and the Investigation site team. Participant recruitment will solely take place at the Investigation site.

Participant identification and recruitment will only commence when the trial has ethical and regulatory approvals in place and confirmation of capacity and capability for local site approval has been issued.

Participants randomised into the trial will be eligible to receive up to £35 per visit for reasonable travel and/or subsistence costs. Participants will receive payments even in the event of trial withdrawal, which will be provided on a pro-rata basis for the number of attended visits. Payment will be processed as per the QMUL finance policy, provided that receipts and/or evidence of expenses are given to Investigation site.

### 4.0 Informed Consent Procedures

Informed consent will be obtained prior to the participant undergoing procedures that are specifically for the purposes of the trial and are outside standard, routine care at the Investigation site (this includes collection of patient identifiable data).

The CI has overall responsibility for the informed consent of participants at their site and will ensure that any person(s) delegated responsibility to participate in the informed consent process is duly authorised, trained, and competent to participate according to the ethically approved protocol, principles of Good Clinical Practice (GCP), and Declaration of Helsinki. If delegation of consent occurs, then details will be provided in the Investigation site delegation log.

The CI or Sub-Investigator(s) (as listed on the Investigation site delegation log) will confirm eligibility of all participants on this trial.

The right of a participant to refuse participation without giving reasons will be respected. The participant will remain free to withdraw at any time from the study without giving reasons and without prejudicing their further treatment and will be provided with a contact point where they may obtain further information about the study.

Where a participant is required to re-consent, it is the responsibility of the CI to ensure this is done in a timely manner.

The entire consenting and/or re-consenting process will be documented in the participant medical records, and the participants study file and electronic case report form (eCRF). A copy of the PIS and consent form will be given to the participant, filed in medical records and participant file.

#### 4.1 Incentives

No incentives will be offered to prospective participants. However, we will cover reasonable expenses related to travel and subsistence when attending a visit. Those will be as per QMUL expense reimbursement policies, and they will be limited to maximum of £35 per visit.

#### 4.2 Vulnerable Participant Considerations

Participants deemed vulnerable will not be included in the trial.

#### 4.3 Writing, Reading, and Translation Considerations

The trial documents will not be translated. Participants who have written, reading or require translation considerations will not be included in the trial.

#### 4.4 Participants Lacking Capacity

Participants lacking capacity will not be included in the trial.

The Investigation site team will assess capacity of the participant to consent based on the following criteria:

- Understand the purpose and nature of the research.
- Understand what the research involves, its benefits (or lack of benefits), risks and burdens.
- Understand the alternatives to taking part.
- Be able to retain the information long enough to make an effective decision.
- Be able to make a free choice.

#### 4.5 Minors

Participants below the age of 18 years old and 80 years old or above will not be included in the trial.

### 5.0 Participant Allocation

Participant allocation is based on screening visit inclusion and exclusion criteria assessments (as detailed below in section 7.0 Trial Schedule) to confirm participant eligibility for the trial. All details of the eligibility screening process, including participants who are screened and not randomised, will be documented in the participant study files, medical records and eCRF.

All participants who are consented and screened, taking into consideration the participant's willingness to continue in the trial and confirmation that they are eligible for the trial, will be randomised into the main trial.

Participants that are randomised onto the main trial and who have also consented to the SCRATCH-HTN sub study can proceed with sub-study assessments.

### 6.0 Participant Eligibility Criteria

#### 6.1 Inclusion Criteria

Participants must meet all the following inclusion criteria to be eligible for the trial:

1. Participant has given written informed consent.
2. Participant has sufficient knowledge of the English language to be able understand the participant information sheet and trial materials including outcome assessments.
3. Participant is aged ≥18 years and <80 years at the time of screening visit.
4. Participant is taking between 1 to 4 antihypertensive medications (inclusive) at time of screening and baseline (randomisation) visit and is willing to adhere to no change in medication during the trial until end of the trial visit (visit 5). (NB. Participant on only one antihypertensive medication should be taking that medication for at least six weeks prior to the screening visit).
5. Participant has confirmed diagnosis of hypertension.
6. Participant meets BP criteria:
    - 24-hour ambulatory BP monitoring (ABPM) at either screening visit or baseline (randomisation) visit, with mean daytime SBP of ≥135 mmHg and <170 mmHg and mean daytime DBP of ≥85 mm Hg and <115 mmHg (N.B. By default, Ambulatory Blood Pressure Monitoring [ABPM] at screening visit will be used at baseline visit. However, if there has been an addition of new medication after participant's screening visit, 24-hour ABPM must be repeated at baseline visit, but the screening ABPM will be used for eligibility criteria).
7. Participant has one or more of the following associated conditions:
    a) Obesity: BMI >30 or waist circumference >94 cm (men) or > 80cm (women). (NB. For participants of South-East Asian/Chinese/Japanese origin these cut-offs are >90 cm (men) or >80 cm (women)).
    b) Type 2 diabetes – controlled or sub-optimally controlled (HbA1c ≤8.5% or ≤69 mmol/mol) on diet and/ or medications except insulin.
    c) Heart rate (any one of the three recordings) ≥70 bpm at screening or baseline (randomisation) visit (measurements taken after 5 minutes of rest in a seated position and when finger probe has been placed for a minimum of 30 seconds thereafter) or a heart rate (any one of the three recordings) ≥60 bpm at screening or baseline (randomisation) visit if the patient is taking beta-blocker medication or a rate-limiting calcium channel-blocker medication.

d) HbA1c ≥42 mmol/mol or fasting blood glucose (if available) ≥5.6 mmo/L **AND** either low HDL cholesterol (≤1.03 mmol/L for men and ≤1.29 mmol/L for women) or high triglyceride (triglycerides ≥1.7 mmol/L)

e) Both low HDL cholesterol (≤1.03 mmol/L for men and ≤1.29 mmol/L for women) **AND** high triglyceride (triglycerides ≥1.7 mmol/L)

f) Diagnosed or known case of polycystic ovarian syndrome.

8. Female participants of child-bearing potential (all those below 55 years except if they are surgically sterile, meaning they have undergone a hysterectomy, bilateral tubal ligation, or bilateral oophorectomy, or formally diagnosed by their doctors to be post-menopausal) must agree to use the acceptable methods of contraception from the time of consent until last follow up visit.
9. Participant is able to communicate satisfactorily with the Investigator and Investigation Site staff, and to participate in, and comply with all clinical study requirements.
10. Participants agrees to have all trial procedures performed and is able and willing to comply with all trial visits and protocol requirements.

#### 6.2 Exclusion Criteria

If any of the following exclusion criteria is met, the participant cannot be randomised:

1. Participant is unable and unwilling to use the AffeX-CT device daily.
2. Participant has a small tragus (ie. the size or shape of the tragus is such that it doesn't allow the application of the ear-clips of the AffeX-CT device for a sustained period of time).
3. Participant has a piercing on the tragus of the ear.
4. Participant is diagnosed with atrial fibrillation or other form of cardiac arrhythmia
5. Participant has eGFR <45 ml/min/1.73 $m^2$ at screening visit.
6. Participant has type 1 diabetes mellitus.
7. Participant has type 2 diabetes mellitus on Insulin or those on oral antidiabetic medications with poor glycaemic control defined as HbA1c above 8.5% (or >69 mmol/mol).
8. Participant has a history of falls or symptoms of orthostatic hypotension in the last 3 months prior to baseline (randomisation) visit.
9. Participant is pregnant, nursing or planning to become pregnant within the next 6 months.
10. Participant suffers from chronic pain and has taken anti-inflammatory drugs for two or more days per week over the last month prior to baseline (randomisation) visit.
11. Participant has clinically significant or symptomatic hypertension-mediated target organ damage such as severe heart failure with NYHA 4, end stage renal damage, medically diagnosed/imaging proven stroke, symptomatic peripheral vascular disease, or severe retinopathy.
12. Participant has a history of stable or unstable angina or had an acute coronary event within 3 months prior to baseline (randomisation) visit or had a myocardial infarction within the last six months of enrolment prior to baseline (randomisation) visit.
13. Participant has history of renal denervation within 1 year prior to baseline (randomisation) visit.
14. Participant has a therapeutic implantable electronic/electrical device such as pacemaker, implantable cardioverter-defibrillators (ICDs), implanted vagal stimulators.

15. Participant has history of hospitalization (> 24 hour) for heart failure, or cerebrovascular accidents, or history of stroke diagnosed based on imaging or evidence of specialist diagnosis or any other indirect evidence such as discharge summary or clinical letter (at any time in the past).
16. Participant has mean daytime ABPM pulse pressure ≥ 80 mmHg at screening or baseline (randomisation) visit.
17. Participant has a heart rate <50 bpm at screening or baseline (randomisation) visit (measurement taken after 5 minutes of rest in a seated position and when finger probe has been placed for a minimum of 30 seconds thereafter).
18. Participant has auricular dermatitis.
19. Participant has postural hypotension, defined as a fall > 20mmHg in SBP on standing at 3 minutes (compared with sitting).
20. Participant has a history of hospitalisation for hypertensive emergency or urgency in the last six months of enrolment prior to baseline (randomisation) visit. ‘Hospitalisation’ is defined as admission for more than 24 hours or between 12-24 hours with an overnight stay.
21. Participant is identified as unsuitable to participate by the CI/Sub-Investigator(s) and/or Investigation site team for another reason (e.g., for other medical reasons, laboratory abnormalities, limited life expectancy, etc.).
22. Participants with history of epilepsy and are currently on anti-epileptic medication or those who are not on any anti-epileptic medication but have history of a seizure within last 10 years

### 7.0 Trial Schedule

#### 7.1 Schedule of Treatment for Each Visit

**Visit 1 (Screening Visit)**

The screening period will be a maximum of 28 days (- 1 day).

At the screening visit, all participants will sign an Informed Consent Form (ICF) and will be assigned a participant ID number. Participants will provide the following: demographic information, medical history, social history, and current concomitant medication. Vital signs, height & waist circumference, weight & BMI, ECG, 24-hour ABPM and office BP assessments will be completed. Participants will have their blood taken for pathology assessments, and serum pregnancy test for female participants. In addition female participants (where applicable) will complete a urine pregnancy test. Inclusion and exclusion criteria will be evaluated, and adverse events will be assessed.

**Re-screening**

At the CI’s judgement, the screening visit may be repeated once.

Participants will be required to re-consent into the trial and a new participant ID number will be issued.

**Visit 2 (Baseline and Randomisation Visit), Day 0**

Participants will be requested to bring their anti-hypertensive medication to this visit, so that the Investigation site team can witness administration of these medications at the visit.

At the baseline visit, the following information will be reviewed for data completeness and where required, reassessed to aid in confirming participant eligibility: medical history, demographics, social history, 24-hour ABPM (if medication changes since screening visit), concomitant medication, vital signs and inclusion and exclusion criteria. Weight & BMI, office BP, central BP, 24-hour Holter ECG, 6-minute walk test (6MWT) and echocardiogram will be completed. Participants will have their blood and urine taken for pathology assessments (plasma and serum for trial laboratory), and for pregnancy test for female participants only (serum and urine pregnancy test). Participant eligibility for the trial will be confirmed during this visit; the visit can be completed in two parts to allow time for collation and/or review of all assessment results. Participants will be randomised only after all baseline assessments have been completed and reviewed, and the participant is deemed suitable for the trial.

Following randomisation, participants will be assigned a AffeX-CT device code and will be provided with that AffeX-CT device (including a user manual), device logbook and participant ID card. The participant will be trained to use the device and complete the device logbook. After training with the stimulation setting on the training device, participants will be handed their Affex-CT device and will be asked to do self-stimulation under the direct supervision of the research staff. The participant will also complete four questionnaires (Extent of Adherence scale, Insomnia Severity Index (ISI), EQ-5D QoL, cognitive assessment) and adverse events will be assessed. Participants will have safety blood pressures recorded after 30 minutes of their first self-stimulation.

For participants recruited into the sub-study: Participants will complete ATONT procedure within 5 days of this visit.

**Phone Call, Day 1-4**

On one of the Days 1-4 participants will receive a phone call, where they will be reminded to apply treatment daily for the duration of the initial course of treatment (14 days) and complete AffeX-CT device procedure and device logbook entries. Concomitant medication and adverse events will be reviewed.

**Phone Call, Day 7 (week 1)**

On Day 7 (visit window +/- 3 days) participants will receive a phone call, where they will be reminded to apply treatment daily for the duration of the initial course of treatment (14 days) and complete AffeX-CT device procedure and device logbook entries. Concomitant medication and adverse events will be reviewed.

**Visit 3 – Day 14 (week 2)**

Participants requested to bring their anti-hypertensive medication to this visit, so Investigation site team can witness administration of these medications at the visit.

At visit 3 (visit window +/- 5 days) concomitant medication and AffeX-CT device logbook will be reviewed. The participant will complete the following assessments: vital signs, office BP, 6MWT and demonstrate the correct use of AffeX-CT device including entry in device diary. Participants will also complete three questionnaires (Extent of Adherence scale, EQ-5D QoL Questionnaire and Cognitive Assessment). Concomitant medication and adverse events will be reviewed.

**Visit 4 – Day 28 (week 28)**

Participants requested to bring their anti-hypertensive medication to this visit, so Investigation site team can witness administration of these medications at the visit.

At visit 4 (visit window +/- 5 days) concomitant medication, vital signs, 24-hour ABPM, office BP, 24-hour Holter ECG, 6MWT, use of the device including entry in AffeX-CT device logbook will be completed. Participants will have their blood and urine taken for pathology assessments and blood samples (plasma and serum) for trial laboratory analysis. Participants will also complete five questionnaires (AffeX-CT device Usability, EQ-5D QoL Questionnaire, ISI questionnaire, Blinding questionnaire and Cognitive Assessment) and adverse events will be assessed.

**Text/Email Reminder - Day 42 (week 6)**

On day 42 (visit window +/- 3 days) participants will receive a reminder to complete AffeX-CT device procedure and device logbook entry

**Phone Call, Day 56 (week 8)**

On day 56 (visit window +/- 3 days) participants will receive a phone call, where they will be reminded to complete AffeX-CT device procedure and device logbook entry. Concomitant medication and adverse events will be reviewed.

**Text/Email Reminder – Day 70 (week 10)**

On day 70 (visit window +/- 3 days) participants will receive a reminder to complete AffeX-CT device procedure and device logbook entry.

**Visit 5 (End of Treatment Visit) - Day 84 (week 12)**

Participants requested to bring their anti-hypertensive medication to this visit, so Investigation site team can witness administration of these medications at the visit.

At visit 5 (window +/- 5 days) the participant will complete the following assessments: vital signs, concomitant medication, weight & BMI, 24-hour ABPM, office BP, central BP, 24-hour Holter ECG, 6MWT and echocardiogram. Participants will have their blood and urine taken for pathology assessments and blood samples (plasma and serum) for trial laboratory analysis. The use of the device procedure and entry in AffeX-CT device logbook, will be completed for the final time and both items will be returned. AffeX-CT device logbook entries will be reviewed, and participants will also complete all trial questionnaires (Extent of Adherence scale, Insomnia Severity Index (ISI), AffeX-CT device Usability, EQ-5D QoL Questionnaire, blinding and Cognitive Assessment) and adverse events will be assessed.

For participants recruited into the sub-study: Participants will complete ATONT procedure within 5 days of this visit.

**Extra-visit/clinical assessment (anytime as per clinical need)**

These visits will be offered to the participants as per their clinical need, and particularly if BPs are either high (SBP >180 mm Hg) or low (SBP <100 mm Hg), or they are symptomatic with postural hypotension or there are any other concerns. Participants will be requested to bring their anti-hypertensive medication to this visit, and will be encouraged to bring their home BP records too. At such visit/clinical assessment participant will be clinically evaluated by the physician/investigator. Participants will complete the following assessments: vital signs, concomitant medication, weight & BMI, office BP, standing BPs (if required) and may have extra blood or urine tests as per clinical need.

**Phone Call Follow-Up – Day 112 (Week 16)**

On Day 112, participants will receive a phone call; during the phone call participants will be asked to answer the questions of the EQ-5D QoL Questionnaire, and site staff will complete the questionnaire as per the participant's responses. Concomitant medication and adverse events will also be reviewed.

#### 7.2 Schedule of Assessments

| **Schedule** | Visit 1 (Screening Visit) | Visit 2 ** (Baseline /Randomisation Visit) | Phone Call-1 | Phone Call-2 | Visit 3** | Visit 4** | Text / Email Reminder -1 | Phone Call-3 | Text / Email Reminder-2 | Visit 5** (End of Treatment Visit) | Phone Call-4 Follow-up* |
|---|---|---|---|---|---|---|---|---|---|---|---|
| **Timeline (weeks/days)** | n/a | Week 0 Day 0 | Day 1-4 | Week 1 Day 7 | Week 2 Day 14 | Week 4 Day 28 | Week 6 Day 42 | Week 8 Day 56 | Week 10 Day 70 | Week 12 Day 84 | Week 16 Day 112 |
| **Visit Window** | 28 days (-1 day) | n/a | n/a | +/- 3 days | +/- 5 days | +/-5 days | +/-3 days | +/-3 days | +/-3 days | +/-5 days | +/-3 days |
| **Informed Consent** | X | | | | | | | | | | |
| **Medical History, including demographic information and social history** | X | X[1] | | | | | | | | | |
| **Vital Signs[2]** | X | X | | | X | X | | | | X | |
| **Height & Waist Circumference** | X | | | | | | | | | | |
| **Weight & BMI** | X | X | | | | | | | | X | |
| **Concomitant Medication** | X | X | X | X | X | X | | X | | X | X |
| **24-hour ABPM** | X | X[3] | | | | X | | | | X | |
| **Office BP[4]** | X | X | | | X | X | | | | X | |
| **Central BP[†]** | | X | | | | | | | | X | |
| **24-hour Holter ECG** | | X*** | | | | X | | | | X**** | |
| **6-minute walk test (6MWT)** | | X | | | X | X | | | | X | |
| **Echocardiogram** | | X*** | | | | | | | | X**** | |

| | | | | | | | | | | | |
|---|---|---|---|---|---|---|---|---|---|---|---|
| **Electrocardiogram (ECG)** | X | | | | | | | | | | |
| **Blood Test[5]** | X | X | | | | X | | | | X | |
| **Blood Samples for storage and later evaluations (Plasma & Serum)** | | X | | | | X | | | | X | |
| **Urine Pregnancy** | X | X | | | | | | | | | |
| **Urine Sample[6]** | | X | | | | X | | | | X | |
| **ATONT Assessment†** | | X[7] | | | | | | | | X[8] | |
| **Inclusion and Exclusion** | X | X[9] | | | | | | | | | |
| **Randomisation** | | X | | | | | | | | | |
| **Device procedure and logbook** | | X[10] | X[11] | X[11] | X[12] | X[12] | X | X[11] | X[11] | X[12, 13] | |
| **Extent of Adherence scale (Voils[23]) Questionnaire** | | X | | | X | | | | | X | |
| **Insomnia Severity Index (ISI) Questionnaire** | | X | | | | X | | | | X | |
| **Blinding Questionnaire** | | | | | | X | | | | X | |
| **AffeX-CT Device Usability Questionnaire** | | | | | | X | | | | X | |
| **EQ-5D QoL Questionnaire** | | X | | | X | X | | | | X | X |
| **Cognitive Assessment** | | X | | | X | X | | | | X | |
| **AE Reporting** | X | X | X | X | X | X | | X | | X | X |

| | |
|---|---|
| * | All participants will be offered a safety reporting follow-up and permitted to change antihypertensive medication (if needed) after tAN procedure has ceased/ participants are not using the device. Participants will be encouraged to keep a record of their HR & BPs at home during this period however this is not mandatory. This follow-up is to assess and collate AffeX-CT device safety and efficacy prolonged data. |
| ** | All participants will be asked to **not** take their morning medications prior to the visits; they will be asked to bring their medication with them and administer their medication(s) in the Investigation Site, after their office BP measurements have been taken. If the participant has taken the medication prior to the visit, site staff will make a note on the medical files and eCRF of the participant. If the participant has not taken their medication to the site and did not receive their medication prior to the visit – and site staff cannot arrange to obtain the medication locally – the visit will be re-scheduled. |
| *** | Baseline Holter and/or Echocardiogram that is conducted within a period of -28 to +3 days from the randomization/baseline visit, will be allowed if there are no changes in the blood pressure treatment between the time of that investigation and randomization |
| **** | Visit 5 Holter and/or Echocardiogram that is conducted within a period of -5 to +14 days of the Visit 5 clinic date |
| † | To be complete for sub-study only. |
| X[1] | Any incomplete medical history sections to be reviewed and completed. |
| Vital signs[2] | Pulse rate, respiratory rate, temperature and oxygen saturation assessments. |
| X[3] | Screening 24-hour ABPM will be used at baseline, only if within screening period <u>and</u> no subsequent treatment changes have been made. Otherwise, 24-hour ABPM must be repeated at baseline (randomisation) visit. |
| Office BP[4] | 3 readings will be performed and average mean (after excluding first of 3 BP readings) calculated. |
| Blood Test[5] | 1 SST, 2 EDTA and 1 Oxalate; Full blood count (FBC), lipid profile, glucose (fasting), HbA1c, fructosamine, U&Es, Serum pregnancy for female participants on screening and randomisation visits. |
| Urine Sample[6] | Urinary albumin creatinine ratio and urinary antihypertensive drug screen. |
| Blood samples for storage and later evaluations (plasma and serum) | 2 SSTs and 1 EDTA; These will be collected and stored for future analysis, if need (i.e., in case new information arises or if there is a safety signal, and or further investigation is required. The stored samples will be used for the sub-study analysis in a batch basis, and will be used if there are any signals that will indicate that a more mechanistic or pathophysiological insight is required. |
| X[7] | For subgroup only - ATONT assessment on both arms will completed with participants' consent at baseline visit and/or within 5 days after baseline & randomisation visit; Investigation site team will endeavour to complete the assessment at the baseline & randomisation visit. |

| | |
|---|---|
| X[8] | For subgroup only – ATONT assessment on both arms will completed at end of treatment visit and/or within +/-5 days of the visit window. |
| X[9] | Inclusion and Exclusion criteria will be reviewed and any incomplete sections subsequent to screening visit will be completed. |
| X[10] | Device and training will be provided to the participant. Following training participants will be observed using the device during the visit to ensure they are competent with AffeX-CT device handling. Participants will be observed for 30 minutes after self-stimulation within the research facility, and their sitting blood pressure (X3 times) will be recorded before they are allowed to go home. |
| X[11] | Participants will be reminded to complete AffeX-CT device procedure and logbook entry. |
| X[12] | AffeX-CT device logbook will be reviewed. |
| X[13] | Participants will stop tAN Procedure and will return the device. |

Notes:

- Participants are required to complete device procedure daily from Day 0 to Day 14 and once weekly after Day 14 to Day 82.
- Participants will receive introduction to device and overview of how the device works with opportunity to demo and ask questions at screening visit.

#### 7.3 Randomisation Method

Eligible participants will be randomised at the baseline visit to either the active (tAN) or sham (sham-tAN) device arm of the trial at a ratio 2:1 ratio. Double the number of participants will be randomised to active therapy in order to collect more data, particularly safety data, for the active intervention.

The dynamic minimisation approach will be used for randomisation with balancing factors of age at baseline (categories: <65; 65+ years), sex, BMI at baseline (categories: <30; 30+ kg/$m^2$), and baseline mean daytime SBP (categories: <160 mmHg; 160+ mmHg) to help achieve a balance in these prognostic risk factors between the randomised trial groups.

A trial specific Randomisation, Unblinding and Maintaining the Blind SOP will detail all randomisation, blinding and unblinding trial procedures.

#### 7.4 Randomisation, Blinding and Unblinding Procedure

Sealed Envelope, an online EDC randomisation tool will be used and recorded as part of the eCRF for this trial. The randomisation tool will be available to CI, Sub-Investigator (s), Investigation site, and participant details (Participant ID, subject initials, date of birth, sex, mean daytime ABPM, BMI and Age (<65 years or ≥ 65 years)) and eligibility criteria will need to be entered in Sealed Envelope for the randomisation function to be enabled. Once randomised, the participant will be allocated a specific device code, and the corresponding device (coloured-coded) will be identified by the research team at WHRI, and given to the participant for use during the trial. The randomisation confirmation from Sealed Envelope will be printed for the participant study file.

Sponsor and CVCTU Coordinating team will have access to Sealed Envelope for monitoring purposes. A master randomisation/allocation list will also be held by Afferent Medical Solutions Ltd. The Investigation site team will complete a corresponding enrolment log to track recruitment.

The CI is responsible for the medical care of each individual trial participant (Declaration of Helsinki section 3, and GCP section 4.3) and the coding system in blinded studies should include a mechanism that permits rapid un-blinding (ICH GCP 5.13.4).

As per Sponsor requirements, a 24-hour emergency unblinding procedure will be available in this trial. Participants will also be given participant trial card with emergency CI and Investigation site contact details.

The CI and Sub-investigators are responsible for unblinding on the trial, and unblinding of a participant allocation will be via Sealed Envelope.

If the person requiring the unblinding is not associated with the CI/Sub-Investigator (s) and/or the Investigation site, the requesting health care professional will notify the Investigator site team that an unblinding is required for a trial participant, and an assessment to unblind the participant will be made in consultation with the Investigation site and CI.

The CI will be notified in writing as soon as possible the necessity of the code break.

Once the participant allocation is revealed, and the requester is notified, the CI/Sub-Investigator (s) or Investigation site team will detail the reason for the unblinding on the eCRF, participant trial file and electronic participant records/clinical notes.

The sponsor's and manufacturer's safety reporting teams will be unblinded to the trial intervention. In the event that a safety event must be reported to the MHRA unblinded, a member of these teams will check the participant's allocation via Sealed Envelope and submit an unblinded report, without the need to unblind the study team.

All trial unblinding will be disseminated to the respective Trial Committee for review in accordance with the charter, where relevant. Participant unblinding will be documented in the End of Trial and statistical reports.

A trial specific Randomisation, Unblinding and Maintaining the Blind SOP will detail all randomisation, blinding and unblinding trial procedures.

#### 7.5 Trial Assessments

The following trial assessments will be recorded as per Section 7.2 schedule of Assessments:

**Demographics**
The following demographic information will be collected as part of the screening: gender, initials, date of birth, age, marital status and ethnicity (self-identified).

**Social History**
The following social history information will be collected as part of the screening: education level (GCSE, A Levels, Graduate), smoking status, alcohol status, diet (self-identified), salt intake (mild/moderate/high) and level of activity.

**Medical History**
The following medical history will be collected as part of the screening: all past and current diagnosed medical conditions including incidence of diabetes, hypertension (duration), stroke, heart attack, chronic kidney disease, any cancer and any organ transplant.

**Concomitant Medication**
The following concomitant medication will be collected: all current medications including pain killers/ NSAIDS, vitamins and minerals and anti-hypertensive BP medications.

**Vital signs**
The following medical history will be collected: pulse rate, respiratory rate, temperature and oxygen saturation assessments.

**Height, waist circumference and weight**

Height, waist circumference will be measured at screening visit only. Weight will be measured at each study visit.

**24 hour ambulatory BP monitoring (ABPM)**
Ambulatory blood pressure will be monitored on visits 1 or 2,and visit 4 and visit 5. The ABPM will be conducted only after the participant has confirmed that they have taken their morning medication. Refer to schedule of assessment for actions to be taken if the participant has not taken their morning medication. Only key summary results will be entered into the eCRF. The detailed results will be kept in the participant's medical file. Participants will be asked to make a note while doing ABPM of their bedtime and wake up time. This information will be entered in the eCRF, if available.

The Investigator will decide the appropriate course of action, if mean daytime falls outside the ranges; SBP ≥170 mmHg or <110 mmHg, mean daytime DBP ≥115 mm Hg and <55mmHg.

**24-hour Holter ECG**
24-h Holter monitoring will be done on visits 2, 4 and 5. Only key summary results will be entered into the eCRF. The detailed results will be kept in the participant's medical file.
Patients will be consulted after 26 patients have completed visit 4 to determine whether the number of assessments is too burdensome..This will be done by asking the patient in person during their scheduled visit 4. This will not effect the primary, secondary or exploratory objectives of the study.

**Office BP**
Office BPs will be taken ideally before the subject has taken their morning anti-hypertensive medication on the non-dominant arm (one set of three measurements separate by 1-2 minutes using the same cuff). Three office BP measurements will be obtained. The mean of the last two BP readings will be calculated. The Investigator will decide on the appropriate course of action if mean office BP (which is mean of last two readings from 3 BP recordings) falls outside the ranges; SBP ≥180 mmHg and <110 mmHg and DBP ≥120 mmHg and <60 mmHg.

**Central BP**
Central BP will only be taken for sub-study patients at visits 2 and 5. A Sphygmocor device will be used to capture brachial waveforms after fitting a standard brachial cuff (which also takes systolic and diastolic BPs). These waveforms are interpreted by the software and converted to the central aortic waveforms. The platform also provides with a print-out of central aortic SBP, central PP and central DBP. Those values will be transcribed in the eCRF. In case the solitary equipment for assessing these central BPs, i.e., Sphygmocor device/software is not working, a Vicorder device will be used as a back-up, or the central recordings will not be taken and a relevant note will be made on the eCRF. Since, this is an exploratory endpoint, any extra-visit solely for this purpose will not be scheduled.

**Echocardiogram**

Echocardiogram will be obtained at baseline and the end of the trial visit (visit 5). Left ventricular ejection fraction (LVEF), left ventricular mass (LVM), relative mass thickness (RMT), left atrial volume (LAV), left ventricular end-diastolic pressure (LVEDP), and E/e´ ratio will be captured in the eCRF, and the detailed report will be kept in the participant's medical files.

**AffeX-CT device logbook & Review**

AffeX-CT device logbook will be completed by the participant's through Days 1 to 14 and once a week during weeks 3 to 12. Participants will be shown the AffeX-CT device logbook during their screening visit, and will be trained on how to complete it on the baseline visit.

**Questionnaires**
Participants will be asked to complete the following questionnaires as per schedule of assessments:

- Adherence to current antihypertensive medications will be assessed using the Extent of Adherence (EoA) questionnaire [23]
- tAN procedure compliance/accountability questionnaire, including ease of use of Affex-CT device assessed using a participant feedback visual analogue scale (VAS).

- Nature, severity and impact of insomnia will be assessed using the Insomnia severity Index (ISI) questionnaire [25]
- Success of the blinding procedure assessed using a blinding index at 1 month and 3 months. We will do this for both participants and the trial team in direct contact with the participants.[24]
- Quality of Life will be assessed using the EuroQOL five dimensions questionnaire (EQ-5D)

**Cognitive/memory assessment**

Cognitive/ memory assessment using PEBL2 software will be done on visit 2, 3, 4, and 5. [22]

**Safety Blood and Urine Assessments**

| | |
|---|---|
| **Blood samples** | |
| Biochemistry | Lipid profile, glucose (fasting, preferable)<br>HbA1c<br>Fructosamine<br>U&Es<br>Serum pregnancy for female participants on screening and randomisation visits |
| Haematology | FBC |
| **Urine Samples** | |
| Urinalysis | Urine albumin<br>Albumin creatine ratio<br>Pregnancy test (if applicable – at screening and randomisation) |
| Urine antihypertensive drug screen | All common Calcium-Channel Blockers (CCBs)<br>All common Angiotensin-Converting Enzyme Inhibitors (ACEIs)/ **Angiotensin** II receptor antagonists<br>All common Diuretics<br>**Mineralocorticoid receptor** antagonists (MRA) – spironolactone, amiloride<br>Alpha blocker –Doxazosin<br>All common beta blockers<br>Centrally acting – clonidine, minoxidil, moxonidine<br>All common anti-diabetic drugs |

#### 7.6 Follow up Procedures

Participates will be followed-up at 4 months (phone call follow-up), 4 weeks after the end of treatment visit within a window of +/- 3 days.

The following will be performed and recorded during the call:

- Concomitant Medication
- EQ-5D QoL Questionnaire
- AE Reporting

### 8.0 Participant, Study, and Site Discontinuation

It is always within the remit of the responsible physician to withdraw the participant from the study for appropriate medical reasons. This can be (but is not limited to) individual adverse events, new information gained about a treatment, or if it is felt to be in the participant's best interest.

Early termination of the participant from the trial could happen for the following reasons:

1. Withdrawal of consent

2. Development of physical or mental health condition that in the opinion of the participant or the investigator will not allow self-administration of the device stimulation

3. For other medical conditions that are temporary, and are not related to the device therapy, we will allow temporary disruption in the schedule at the discretion of the principle investigator and in consultation with the participant and his/her healthcare providers.

### 9.0 Laboratories and Samples

#### 9.1 Local Laboratories

All research samples will be collected, stored and processed at WHHC and other WHRI facilities. Blood samples for routine safety tests will be transferred to Barts Laboratory:

- William Harvey Research Institute – Heart Centre, Barts and The London School of Medicine and Dentistry, Queen Mary University of London, Charterhouse Square, London, EC1M 6BQ
- Barts Health NHS Trust Pathology Department, St Bartholomew's Hospital, West Smithfield, London, EC1A 7BE

#### 9.2 Sample Collection, Labelling, and Logging

**Screening and safety samples**
Samples to assess eligibility and safety samples to monitor participants safety will be collected by the research nurse or trained phlebotomist at screening (visit 1), baseline (visit 2), visit 4 and visit 5, labelled as per Trust policy and sent to the Pathology department at Barts Health NHS Trust, where they will be processed as per Trust policies. Additional samples may be required to monitor participants' safety.

**Additional samples for future analysis**
Additional samples will be taken for future analysis, if study findings suggest that further analysis is needed. In addition, bloods will be stored in case any additional analysis or unexpected safety analysis and other assessments are needed, during the course of the trial such as: measuring markers of inflammation, liver function tests or auto-immune profiles or

other cardiovascular biomarkers such as acetyl choline. We will be seeking participants consent to do these additional tests during the trial based on the findings and availability of newer technology.
At the end of the trial, all the samples that contain cellular material s will be moved to Barts Bio resource facility (BBRC, www.bartsbioresource.org.uk), or will be destroyed. Participant's consent will be obtained to allow their unused samples to be shifted to the BBRC.

**Amount of blood to be collected during the study**
During the trial period, bloods samples will be obtained at screening (visit 1), baseline (visit 2), 1 month (visit 4) and 3 months visit (visit 5). During the screening visit up to 15 ml of blood will be obtained (no additional blood will be obtained for storage for future analysis). For baseline (visit 2), 1 month (visit 4) and 3 months (visit 5) visits additional blood samples will be obtained; up to 29ml of blood will be obtained.

Baseline visit, 1 month and 3 month visit: total volume up to 29ml. Up to 15 ml for routine tests, and up to14 ml will be collected (in 2 EDTA and 1 SST bottle) that will allow us to separate plasma and serum and store them for future usage. Where possible samples will be aliquoted into 1.0- 1.5 ml of plasma and serum, with at least two aliquots of each.

**Biomarkers samples**
Participants who are eligible for the study will have the option to give informed consent to donate additional blood samples for future biomarkers analysis. Serum and plasma aliquots will be sent to the Barts Bioresource where it will be stored for future research.

**Labelling of blood samples**
Labelling of the samples will include study ID, participant number, date and time when sample was collected and the number of the visit.

#### 9.3 Sample Transfer, Chain of Custody, and Accountability

Blood samples will be collected by research nurse or technician, and will be immediately processed in the WHRI. The safety samples will be labelled as per the Barts NHS Trust standard procedures, and will be under custody of a member of research team until they are handed over to the member of the Barts Pathology laboratory services. Other samples for storage will be stored immediately in the WHRI facilities (see section 9.6). At the end of trial, all stored samples will be transferred to Barts Bioresource (BBR) facility using appropriate dry ice containers physically by a member of the research team and handed over to the member of Barts Bioresource.

#### 9.4 Sample Analysis Procedures

All research samples will be collected, stored and processed at WHHC and other WHRI facilities. Blood samples for routine safety tests will be transferred to Barts Laboratory and will not be transported outside the trust. Those samples will be processed, analysed, stored and destroyed as per the practices and policies at Barts Health Trust.

It is possible that the results of these routine clinical safety analysis could produce findings of clinical significance for participants. If so, we will arrange to notify the individuals concerned, and make the necessary follow up required to manage the findings.

The blood samples for storage will be curated at WHHC facilities, and centrifuged as per the WHHC local SOPs. Plasma and serum aliquoted samples will be stored within WHRI research facilities during the period of the trial at -80ºC.

We will use these samples as per section 9.3. After the end of the study, we will follow the procedures described in section 9.3.

#### 9.5 Sample Storage Procedures

Safety blood samples will be processed, analysed, stored and destroyed as per the practices and policies at Barts Health Trust.

The blood samples for storage for future analysis or research will be curated at WHHC facilities, and centrifuged as per the WHHC local SOPs and study lab manual. Plasma and serum aliquoted samples will be stored within WHRI research facilities during the period of the trial at -80ºC. After the end of the trial, all cellular samples, will be transferred to BBR facility as per the patient's consent, and if there are sufficient quantity left for meaningful research. If such samples have insufficient quantities, those will be destroyed. All stored samples will be labelled with the study identifiers as described in section 9.3. Barts Bioresource is an approved bio-resource by REC (REC ref 14/EE/0007).

- Information about this resource is publicly available, and we will share the link with the trial participants in the PIS – http://www.bartsbioresource.org.uk/about

Samples that are not sent to the BBR and contain cellular information will be destroyed after the trial has ended.

#### 9.6 Sample and Result Recording and Reporting

Screening and safety results will be reported on the participant's medical records, reviewed by one of the Investigators, and recorded on the CRF. Any results deemed by the Investigator to be Clinically Significant will be recorded as an Adverse Event, and will be followed up until resolution or the end of the study participation.

#### 9.7 Sample Management at End of study

Safety samples being sent to Barts Pathology services will be labelled with patient-identifiable information to allow processing, as per Trust guidelines.
All other samples (i.e. Samples that will be stored for future analysis) will be pseudo-anonymised with only the participants study number and stored securely at facilities in WHRI during the period of the study.  At the end of study those samples containing cellular material will be either transferred to BBR or destroyed. Participants will give informed consent for their samples to be stored in BBR for use in future research (see section 9.6).

### 10.0 Investigational Device

#### 10.1 Name and Description of Investigational Device

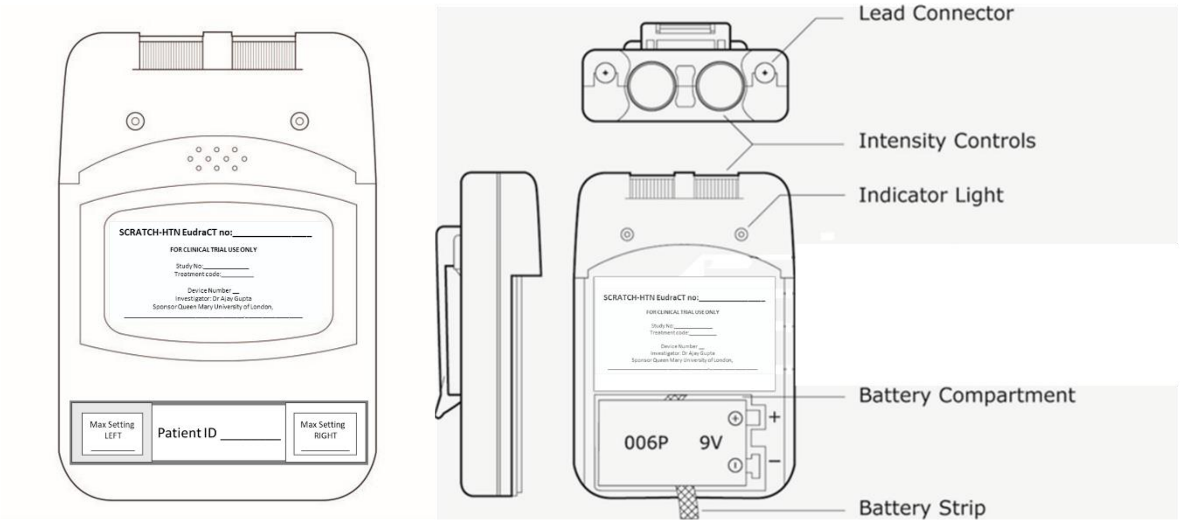

AffeX-CT device comprises of a battery operated control unit, two electrode pairs arranged on ear clips and (connected to the control unit via electrical leads) generates an electrical signal that is used to stimulate the sensory nerves that supply the auricle. The output is comprised of two signals (current pulse waveform) with defined parameters (pulse shape, duration, and frequency). The amplitude of the output signal (strength of the stimulation) is adjustable by the user using control dials. The duration of stimulation is set to deliver the treatment for a fixed period of 30 min.

The AffeX-CT device is based on a Totally TENS unit (model WL-2103A / TT-21AL) manufactured by Well-Life Healthcare Ltd (Taiwan) and distributed by Euromedics GmbH (Germany). The waveform and timing controls are pre-set to the required settings (section 4.8, Intended Performance). Plastic panel is attached to cover the dials and block the user access to the controls.

### 10.2 Intended Performance

AffeX-CT has been set to deliver a biphasic pulse with the following parameters: The performance of each device has been validated to meet the Intended Performance specification.

**Waveform parameters:**

Channel: Dual, isolated between channels

Mode of operation: continuous,

Pulse intensity: recommended range for the investigation is 0.1-8mA (device range is 0-80mA)

Pulse Rate: 30 Hz

Pulse width: 200 µsec

Duration /Time: Continuous for 30min

Wave Form: Bi-Phasic Asymmetrical square pulse

**Electrical specification**:

Battery 9v (6F22)

**Mechanical specification**:

Dimensions: 95(H) x 60(W) x 23(T) mm

Weight: 115 grams (inc. battery)

### 11.0 Legal Status of Investigational Device

AffeX-CT device has not been approved by the MHRA for use as "a non-implanted, non-invasive transcutaneous autonomic neuromodulation device for the functional management of the arterial blood pressure" (Intended Use).

It has been supplied for 'research only' use as "a non-implanted, non-invasive transcutaneous autonomic neuromodulation device for the functional management of the arterial blood pressure" (Intended Use).

The AffeX-CT device is a TENS device (Totally TENS, model WL-2103A/TT-21AL) on which it is based has been marketed as an Over-The-Counter (OTC) TENS device (Intended Use: (with a clinical prescription) for the symptomatic relief and management of chronic (long term) pain. No adverse events have been reported.

The trial device falls within the European Medical Device Regulations (MDR) as a Class IIa device under the MDR (in line with other TENS devices) and Class 2 device by the FDA. The AffeX-CT device uses the Totally TENS, model WL-2103A/TT-21AL product which has been designed and manufactured to ISO13485 Quality Management System.

The device is manufactured using standard electronic components, pcb assembly processes and ABS (Acrylonitrile butadiene styrene) for the casing.

Ear-clips use moulded plastic parts with conductive silicone rubber electrodes.

Each device is designed for use by one participant only and must not be re-used by other participants. There will be in total 75 individual investigational devices supplied for the trial, one for each participant and 12 devices as replacements devices should devices issued to participant fail or malfunction. Each device is marked with a unique identification number. The electrodes supplied with the device will also be marked with the device's unique identification number. Additional devices will be supplied for training purposes and in reserve, if any participants encounter issues or cases of malfunction.

The device is compliant with the following electrical, mechanical and safety standards:

Mechanical:

Robustness EN 60601-1 (Classification IP22)

Water resistance EN60529 (Classification IP22)

Ingres protection EN60529 (Classification IP22)

Electrical:

EMC susceptibility / immunity IEC 60601-1-2/60601-1-2

Low voltage safety IEC 60601-1-2/60601-1-2

Quality Management System used: ISO 13485: 2016

#### 11.1 Device Manufacturer (s) and supply arrangement

For the purpose of the SCRATCH-HTN study, the AffeX-CT device is being used 'off-label' and Afferent Medical Solutions Ltd will assume responsibilities of 'the manufacturer' and must therefore fulfil all the requirements of a manufacturer as set out in the UK Medical Devices Regulations 2002, including notification of a clinical investigation to the MHRA.

Named supplier (Afferent Medical Solutions) (''Afferent'') will assume responsibility for:

- manufacturing
- labelling in accordance with the medical device's regulations 2002,ISO14155 and GCP
- separately testing and calibrating each device prior to the delivery to the trial team
- delivering the investigational units to the research facility
- assuming management of and providing maintenance services
- each device will have a unique identification number which will appear on the device labels. Their associated ear-clip & leads and carry case will also have an identification label with the same identification number as the device.
- disposal of devices.

The Sponsor will be responsible for the management of collecting all units upon trial completion and their return to Afferent Medical Solutions (see CIP section 11.10 destruction, return, and recall devices)

**Supplier information**

| | | | |
|---|---|---|---|
| **Company** | **Afferent Medical Solution Ltd** | | |
| **Register Office** | C/O Clockwise , Brunel House, Fitzalan Road, Cardiff, Wales, CF24 0EB | **Company No** | **12126455** |
| **Contact Person** | Dr Everard Mascarenhas | **Tel** | 07815 444305 |
| **Address** | 6 Almond Av, Ickenham, England UB10 8NA | **email** | |
| **Technical Consultant/ Scientific advisor** | Prof Alexander Gourine | **email** | |
| **Regulatory Consultant** | Dr George Zajicek | **email** | |
| **Medical Advisor** | Dr Andrey Gourine | **email** | |

#### 11.2 Investigational Device Software

The device is a manual unit and has no software installed on it.

#### 11.3 Investigational Device Accessories

AffeX-CT is a standalone device and supplied as following:

- Control Unit
- 4x Ear-clips and connecting leads
- Battery (6F22)
- User Guide
- Instructions for Use (IFU)(see IB Appendix B)
- Storage case (soft bag)

#### 11.4 Packaging and labelling of Investigational Devices

Each device will have the following labels on the front cover:

| | |
|---|---|
| Front | 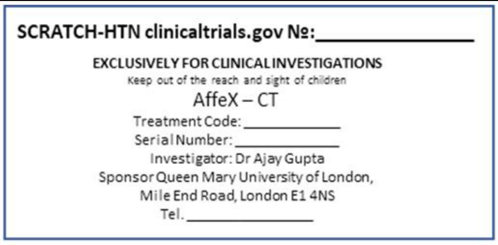 |
| Bottom Front | 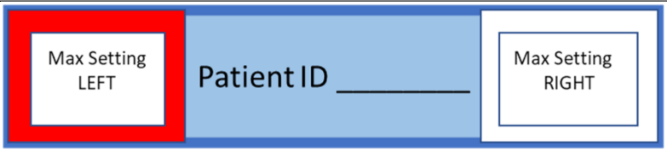 |
| Back cover label | 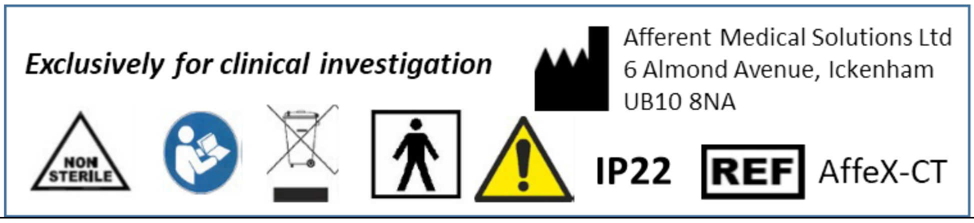 |

Refer to IB Appendix A for more information on Symbols and Nomenclature Description

The carry case will have the information appearing on the front cover label printed on it.

AffeX-CT is a non-sterile product.

There are no special requirements for packaging.

Device will be labelled in accordance with medical device regulations 2002 and ISO 14155 GCP (see IB 10.5; appendix E)

Front labels are to be colour coded; range of colours for participants and white labelled for demonstration/training devices.
The ear-clips will be colour coded, white and black and red and black to allow users to identify which ear-clip should be attached to the left and right ear.

#### 11.5 Accountability and Traceability

Each investigational device will be assigned a unique identification number.

The Investigation site team must maintain records (accountability logs), recording:

- Date at which device (device serial number) was given to the participant (participant number)
- Validation of the output signal parameters
- Device usage log
- Dates of when devices were returned by the trial participants
- Cases of malfunctioning
- Replacements, checks, maintenance
- Unused devices

#### 11.6 Assessment of compliance

Participants will be asked to self-administer and follow the treatment protocol. Compliance with the procedure will be monitored by participants being asked to maintain a device logbook: a log of the date, time and duration of each treatment session, with regular phone-call / text / email reminders:

- Phone Call; one phone call between days Day 1-4
- Phone Call - Day 7/Week 1 (visit window +/- 3 days)
- Visit 3 – Day 14/Week 2 (visit window +/- 5 days)
- Visit 4 – Day 28/Week 4/Month 1 (visit window +/- 5 days)
- Text/Email Reminder - Day 42/Week 6 (visit window +/- 3 days)
- Phone Call – Day 56/Week 8/Month 1 (visit window +/- 3 days)
- Text/Email Reminder – Day 70/Week 10 (visit window +/- 3 days)
- Visit 5 (End of Treatment Visit) - Day 84/Week 12/Month 3 (visit window +/- 5 days)
- Phone Call Follow-Up – Day 112/Week 16

In addition, the device logbooks will be examined and the compliance with the device use will be assessed during the 3 visits (visit 3, 4 and 5) by the study team.

Compliance to the existing antihypertensive medications will be assessed by the Extent of Adherence (EoA) questionnaire [23] and urinary antihypertensive drug screening.

Compliance with the device usage is one of the acceptability criteria that is being evaluated. We will use 80% or more days of device usage out of the maximum possible to define as good usage, and less than 33% days of device usage as poor usage. For per-protocol analysis, we will use data for those who have good device usage.

#### 11.7 Device storage

Trial participants will be urged to store units at room temperature away from moisture and away from other electrical devices and strong magnetic fields:

- Range: 0°C to 38°C (32°F to 100°F)
- Humidity: 10% to 90%
- Barometric Pressure: 80 to101 kPa

After use, participants will be urged to unplug the ear clip leads from the unit, and place all components back into the sachet/casing, in such a way, as to minimise the chance of accidental removal or damage (e.g., drawer or shelf)

Devices held at the research site must be stored in a secure location (e.g., locked cupboard), with access limited strictly to members of the investigation team.

#### 11.8 Device training and experience requirements

Prior to the trial start, a training session will be conducted by an Afferent trainer for members of the Investigation site team. In case of any additional inquiries, contact details to a delegated Afferent representative, as well as the trainer, will be provided, to respond to any requests and for provision of any additional information.

The Investigation site team will be responsible for training participants in the use of the device. They will also be required to follow a set training routine, which will include an assessment of the participant's ability to satisfactorily use the device.

#### 11.9 Administration of investigational device

Target Participants: Participants with uncontrolled and drug-resistant arterial hypertension. (see section 6.0 of the CIP for more information on participant eligibility criteria)

tAN treatment can be started by attaching an electrode clip to the tragus of each ear and turning 'ON' the stimulation signal by turning the dial clockwise from the off position (marked as 0 on the dial to position 4 or 5 on the dial.  Once the participant experiences a tingling sensation, the signal amplitude is reduced by turning the dial anti-clockwise for 1 division below the tingling sensation threshold. The AffeX-CT device will then continue to apply the stimulation for a fixed duration. (The electrode must remain attached to the tragus of the ear for the entire treatment session.)  When this period has ended, the device will automatically stop the treatment session.

The jack-plug of each electrode is inserted into the socket of the AffeX-CT unit and clipped onto the tragus. It is important that both electrodes are connected, one for each ear. Each stimulation is applied for 30 min daily, preferably in the evening time (18.00-22.00, but preferably around 20.00) for 14 consecutive days. After that the stimulation repeated once per week for 10 additional weeks.

Participants will set up the stimulation one level below the threshold throughout their participation in the trial, provided that they do not feel any tingling sensations subsequently. However, they will be advised to dial the voltage to one degree down and try again, if they do feel any tingling at any point during the trial. Participants will be asked to note this in their log-book. Participants will be asked to refer to the User Guide and Instructions for Use (IFU), see Appendix B of the IB for further information.

#### 11.10 Destruction, return, and recall devices

All units are to be returned to Afferent Medical Solutions upon trial completion. Afferent will arrange for the disposal of all the units in an appropriate and controlled manner, upon the request from the investigator. The Sponsor should return the units to Afferent Medical Solutions Ltd, 6 Almond Avenue, Ickenham UB10 8NA, UK.

Separate ear-clips will be used to train participants and are to be disposed of by the Research Nurse in accordance with the clinical unit's standard operating procedures.

Regulations require that disposal of electrical and electronic equipment, including used and unused medical devices, is handled in a controlled manner. Devices that may be contaminated after use or that may contain chemicals or elements that may be hazardous to people or the environment will be disposed of in accordance with the applicable government regulations.

Afferent will be responsible for the disposal of all devices and will ensure their disposal complies with RoHS and in accordance with the applicable government regulations.

In case of a device recall, Afferent will be responsible for coordinating with the investigator and managing the process in a controlled manner.

#### 11.11 Usage schedules

Each stimulation is applied for 30 min daily in the evening time (between 18.00-22.00) for 14 consecutive days. After the 14 days, participants will be required to stimulate once per week for 10 additional weeks. Participants will be encouraged by investigation site staff to apply this stimulation when they are resting/relaxing/or in sitting or lying position, for example, when watching television, listening to music (but not when wearing the headphones or airpods) or reading a book, or at the end of the day retiring to the bed. During device usage, participants will be advised to abstain from any moderate or heavy physical activity (such as exercise, running, continuous walking or lifting weights etc.).

AffeX-CT will be pre-set to deliver a stimulation signal for 30 minutes. (Other parameters of the stimulation signal will also be pre-set, the control for adjusting these parameters will not be accessible by users/participants.)

#### 11.12 Usage modifications and delays

We do not envisage any modification in device usage or device itself during the conduct of the trial, except if a person develops local irritation/dermatitis which makes it difficult to use the clips to the ear. In that case, we will withhold treatment until the person can use the device or try to consider alternative options.

There are no stopping rules for device, except for the inability to use the device because of ear/tragus inflammation or allergy. If there is significant blood pressure lowering to make a person symptomatic, we will first reduce their anti-hypertensive medications as per the standards of clinical care and only then consider stopping the device treatment. Based on the results of our PoC study we do not anticipate this extremely rare event.

As mentioned above, we can consider modification of the tragus access in special circumstances and will explore the alternatives as per participants' wishes/request. In case we cannot come up with any viable and practical alternatives, we will stop the device usage, but continue to follow the patient as per the protocol.

Most likely scenario we anticipate is the interruption of a treatment session. It is possible that patient stops the device or removes the electrode clips from the ear(s) before full 30 minutes. In that case, we advise the participant to use the device again for the remaining period on the same day and record the interruption. If they are not able to use the device for full 30 minutes on the same day, then they have the following option depending on the stage they are in the trial.

a. If they are in the first 14 days (initial course of treatment), with daily use protocol. They can continue using the device as prescribed on the next and following days. We deem more than 11 or more stimulation out of the total 14 in the first stage as being compliant with the first phase.
b. If they are on weekly stimulation phase, and unable to take full 30 minute stimulation on the prescribed day. They can do this next day or day after (up to 5 days after the prescribed stimulation day), and thereafter continue with weekly stimulation, as before.

#### 11.13 Management of adverse events(including device-specific events)

Any adverse events (AEs), including device-specific AEs, will be recorded in the patient record file. SAEs will be reported to the sponsor and CVCTU in pseudoanonymised format as specified in section 13.8.

Any non-serious device related AEs will be reported to the device manufacturer on an ongoing basis at regular monthly intervals.
All device related SAEs will be informed to the device manufacturer (Afferent Medical Solution Ltd) as soon as possible after the investigation site staff has become aware of the event. Details of the device-specific adverse events will be sent to the Afferent team on the following, and if necessary, a member of the CVCTU team will contact the Afferent team via phone at 07724578883.
We anticipate following side effects associated with tAN applied via electrical stimulation for the tragus:

- Light-headedness
- Fatigue/tiredness
- Mood changes
- Neck pain
- Tooth pain
- Pain/local skin irritation due to attachment of the device ear clips
- Ventricular extrasystoles (increased frequency)

Some participants may experience a tingling sensation when using the device. This will be documented on the participant diary card but does not need to be documented as an AE. This is an expected sensation when stimulating nerves using this device in the majority of participants.

Participants will also be requested to record, if experienced, any of the anticipated side effects in the device logbook. AEs will be recorded on the appropriate AE pages of the CRF corresponding to the visit or interaction with the study team.

Serious adverse events (SAEs) require expedited reporting to the Sponsor or designee regardless of relationship to study device or study procedure. (refer for more information to IB 9.3). All those will be informed as soon as possible and within 24 hours of the Investigator/ Investigation site staff become aware of the event.

Any Device related SAEs are not anticipated. However, if they occur, the PI and Investigation site will follow Sponsor's SOPs for reporting and managing them.

See CIP section 13 for further information.

#### 11.12 Potential interactions with other therapies

We do not anticipate any adverse interactions between existing medications and tAN treatment, except it is possible that those on beta-blockers may have an increased BP lowering effect with the use of device. This is because tAN is likely to augment vagal tone, whereas beta-blockers inhibit sympathetic effects, and both of these could be additive for the patient. Similarly, drugs that increase the parasympathetic stimulation, such as clonidine may also increase the BP efficacy of the device. However, clonidine is rarely used nowadays, and certainly not amongst first line medications for the treatment of high blood pressure.

##### 11.13 Recommended concurrent treatment

Not applicable. We do not recommend any concurrent treatment.

##### 11.14 Prohibited therapies

Nil, except the participant should not have any renal denervation therapy in the previous 12 months.

##### 11.15 Study restrictions

No changes in the antihypertensive medications are allowed during the trial. Participants will be offered a safety reporting follow-up and permitted to change antihypertensive medication (if needed) after tAN treatment has ceased/ participant is not using the device.

##### 11.16 Management of incorrect usage

Participants will be urged to keep a device logbook/ to track stimulations and keep to a strict protocol to prevent having more than one treatment session on a single day.

Participants will be instructed to keep stimulation below the threshold at a comfortable level.

##### 11.17 Precautions regarding contraception

Participants pregnant, nursing or planning to become pregnant within the next 6 months are to be excluded from the trial (see CIP section 6.2 for exclusion criteria; see CIP section 13.12 Pregnancy for more information)

##### 11.18 Arrangements for post-study access to the investigational device

We will follow-up all participants for a period of four weeks after the completion of trial. During this period changes to blood pressure treatments, if required, are acceptable. We will not provide any device to the participants, as this is investigational device. The devices are returned to Afferent, we can store these after they have been returned to us. We should stipulate a time period say 12months i.e.. Afferent will retain all the devices used in the trial for a period of 12 months after the completion date of the trial.

#### 12.0 Equipment and other Devices

Not applicable

#### 13.0 Safety Reporting

#### 13.1 General Definitions

| Term | Definition |
|---|---|
| Adverse Device Effect (ADE) | An adverse event related to the use of a medical device. This includes any adverse event resulting from insufficiencies or inadequacies in the instructions for use, the deployment, the implantation, the installation, the operation, or any malfunction of the medical device. This also includes any event that is a result of a use error or intentional misuse. |
| Adverse Event (AE) | Any untoward medical occurrence, unintended disease or injury, or untoward clinical signs (including abnormal laboratory findings) in participants users or other persons, whether or not related to the *investigational medical device* and whether anticipated or not. |
| Adverse Reaction (AR) | An untoward and unintended response in a participant to an investigational medicinal product which is related to any dose administered to that participant.<br>The phrase "*response to an investigational medicinal product*" means that a causal relationship between a study medication and an AE is at least a reasonable possibility, i.e., the relationship cannot be ruled out.<br>All cases judged by either the reporting medically qualified professional or the sponsor as having a reasonable suspected causal relationship to the study medication qualify as adverse reactions. |
| Device Deficiency | Inadequacy of a medical device with respect to its identity, quality, durability, reliability, usability, safety, or performance. |
| Serious Adverse Device Effect (SADE) | Adverse Device Effect that has resulted in any of the consequences characteristic of a serious adverse event. |
| Serious Adverse Event (SAE) | A serious adverse event that led to any of the following:<br><br>a) death<br>b) serious deterioration in the health of the subject, users, or other persons as defined by one or more of the following:<br>a. a life-threatening illness or injury, or<br>b. a permanent impairment of a body structure or a body function including chronic disease, or<br>c. in-patient or prolonged hospitalisation<br>d. medical or surgical intervention to prevent life-threatening illness or injury, or permanent impairment to a body structure or a body function<br>c) foetal distress, foetal death, a congenital abnormality, or a birth defect including physical or mental impairment.<br><br>NOTE: The term "life-threatening" in the definition of "serious" refers to an event in which the participant was at risk of death at the time of the event; it does not refer to an event which hypothetically might have caused death if it were more severe. |
| Serious Adverse Reaction (SAR) | An adverse event that is both serious and, in the opinion of the reporting Investigator or medical assessor, believed with reasonable probability to be due to one of the study treatments, based on the information provided. |
| Unanticipated Serious Adverse | Serious Adverse Device Effect which by its nature, incidence, outcome has not been identified in the risk assessment |

| |
|---|
| Device Effect (USADE) |

#### 13.2 Site Investigator Assessment

The CI is responsible for the care of the participant, or in his absence an authorised medical practitioner (as listed on the delegation log) is responsible for assessment of any event for:

- **Seriousness**
  Assessing whether the event is serious according to the definitions given in section 13.1.
- **Causality**
  Assessing the causality of all serious adverse events in relation to the study treatment according to the definition given. If the SAE is assessed as having a reasonable causal relationship, then it is defined as a SADE.
- **Expectedness**
  Assessing the expectedness of all SADEs according to the definition given. If the SADE is unexpected, then it is defined as a USADE.
- **Severity**
  Assessing the severity of the event according to the following terms and assessments. The intensity of an event should not be confused with the term "serious" which is a regulatory definition based on participant/event endpoint criteria.
    - **Mild**: Some discomfort noted but without disruption of daily life
    - **Moderate**: Discomfort enough to affect/reduce normal activity
    - **Severe**: Complete inability to perform daily activities and lead a normal life

#### 13.3 Reference Safety Information (RSI)

Reference Safety Information (RSI) is the information used for assessing whether an adverse reaction is expected.

Investigator Brochure (IB) is the information used for assessing whether an adverse reaction is expected in this trial.

#### 13.4 Notification and recording of Adverse Events (AEs), Reactions (ARs), Adverse Device Effects (ADEs) or Device Deficiencies

All AE, ARs, ADEs and/or device deficiencies are to be documented in the participants' medical notes or other source data documents and the CRF. Once assessed, if the AE is not defined as Serious, the AE is recorded in the trial file and the participant is followed up by the Investigation site team until the participant's last follow up visit.

#### 13.5 Notification of AEs of Special Interest (AESIs)

Not applicable.

#### 13.6 Adverse Events That Do Not Require Reporting

All AEs will be captured on the eCRF and the participant's study files. The period for AE reporting will be from visit 1 (screening visit) until the post-study follow-up call (Day 112). Refer to section 11.13 for more details on anticipated side effects and their reporting.

Patients may feel a slight tingling sensation when using the device. This is an expected sensation and not an adverse event. Any tingling while using the device will be recorded in the patient diary.

#### 13.7 Device Adverse Reporting Failures, Malfunctions and Re-use

All device deficiencies will be recorded on the clinical investigation device deficiency log and where appropriate in the participant's medical records.

Inadequacy of a medical device with respect to its identity, quality, durability, reliability, usability, safety or performance must be reported as a device deficiency. This could include malfunctions, user errors and inadequacy in the information supplied by the manufacturer, including labelling.

The PI must confirm assessment on the Device Deficiency Log as to whether the device deficiency could have caused a SADE:

- If suitable action had not been taken or
- If intervention had not been made or
- If circumstances had been less fortunate

Device deficiencies which could have caused a SADE must be reported to the Sponsor and device manufacturer within 24 hours of becoming aware of the event by submitting a Device Reporting Form to the following:
Sponsor:
Afferent:
 Device deficiencies must be recorded and reported throughout the Clinical Investigation.

AffeX-CT has been specifically designed to be used by only one patient and must not be re-used by other patients.

#### 13.8 Notification and reporting of Serious Adverse Events (SAEs) and Unexpected Serious Adverse Device Events (USADEs)

Refer to IB sections 9.2 and 9.3.

All Serious Adverse Event (SAEs) will be recorded in the participants' notes, the eCRF. The Sponsor SAE form and reported to the Sponsor (Joint Research Management Office via mailbox at), Afferent Medical Solution Ltd. ( and to the CVCTU (via mailbox at) within 24 hours of the CI or Sub-Investigator (s) becoming aware of the event.

Nominated Sub-Investigators (as listed on the Investigation site delegation log) will be authorised to sign the SAE forms in the absence of the CI at the participating sites.

SAEs and reportable device deficiencies must be reported from consent until the participant's last follow-up visit.

#### 13.9 Sponsor Medical Assessment

Sponsor has delegated the responsibility for oversight of investigational device safety profile and medical assessment of safety events to the CI as medical assessor. The CI must review all SAEs and reported device deficiencies within 72 hours of receipt. This review should encompass seriousness, relatedness, and expectedness. Day 0 for all reported events is when the event is received by the CI and/or CVCTU coordinating team and/or Sponsor (whichever is first).

The CI must also maintain oversite of non-serious AEs reported on the eCRFs and review them periodically to confirm agreement.

It is expected that the CI will achieve oversight of AffeX-CT device safety profile through the trial committees as per section 25.0 of the CIP.

#### 13.10 Procedures for Reporting Blinded safety events

The CI, as Sponsor medical assessor, will assess the event blinded for all possible active and placebo AffeX-CT device procedures. The sponsor's and device manufacturer's safety teams will be unblind and can submit unblind safety reports to the MHRA as required.

#### 13.11 Urgent Safety Measures

The CI may take urgent safety measures to ensure the safety and protection of the clinical study participants from any immediate hazard to their health and safety. The measures should be taken immediately. In this instance, the approval of the Competent Authority prior to implementing these safety measures is not required. However, it is the responsibility of the CI to attempt, where possible, to discuss the proposed change with the sponsor and Medical Advisor at the MHRA (via telephone) prior to implementing the change if possible.

The CI has an obligation to inform both the MHRA and Research Ethics Committee in writing **within 3 days** of implementing the Urgent Safety Measure. They must also submit a substantial amendment documenting the changes with 14 days of implementing the urgent safety measure. The Sponsor must be sent a copy of the correspondence with regards to this matter as soon as it is sent.

#### 13.12 Pregnancy

If a participant becomes pregnant whilst involved in this trial, it is not considered to be an SAE or an AE. However, it is an event that requires reporting, monitoring and follow up. If a participant or participant's partner becomes pregnant whilst or within 4 months from the end of their participation in the study, the sponsor should be notified immediately (within 24 hours of site becoming aware of the pregnancy) using the sponsor pregnancy form. The pregnancy reporting procedure will be the same as the SAE reporting route.

The CI (in conjunction with the site PI) should determine if the foetus has been exposed to AffeX-CT device. The CI has the responsibility to ensure that the pregnancy form is completed and sent to the sponsor within the agreed timelines. The initial report should be sent within 24 hours of the CI or Sub-Investigator(s) becoming aware of the event and follow up information submitted as and when it becomes available up to agreed follow up time after birth.

The Sponsor will arrange for a review of the pregnancy report by an appropriate expert medic (usually a consultant obstetrician). The study team must follow all instructions provided by the Sponsor's expert.

### 14.0 Annual Reporting

#### 14.1 Annual Progress Report (APR)

N/A.

### 15.0 Statistical Considerations

#### 15.1 Sample Size Calculation

This is a pilot study designed to collect data required to develop a larger efficacy trial.

The trial is powered in relation to the primary endpoint of change in daytime ambulatory SBP between baseline and the end of the treatment at 3 months in the active treatment arm. This was done using a paired t-test approach.

Using a conservative assumption of a mean change in SBP of 5.5 mmHg with a standard deviation of 11 mmHg, based on existing data for hypertensive patients, 34 participants would give 80% power to detect such a change at the two-sided alpha level of 0.05. After inflation for a potential 10% drop-out and a further 10% non-compliance level, we require 42 subjects in the intervention arm.

The study will also recruit participants to be randomised to a comparator, sham treatment arm, in order to compare changes in SBP to a control and take into account potential Hawthorne and placebo effects. In order to collect more data, particularly safety data, on the active treatment than the sham treatment, the trial will recruit double the number of subjects for the active treatment than for the sham treatment, a randomisation ratio of 2:1. Hence the sample size for the sham treatment will be 21, giving a total sample size across both arms of 63 subjects.

With the sample size of 63 subjects and 2:1 randomisation ratio, we would need to observe a mean difference in SBP between the groups of 8.4 mmHg with no drop-out or non-compliance, or 9.3 mmHg if 20% of subjects either drop-out or are non-compliant, to be able to detect the difference at the two-sided alpha level of 0.05, with 80% power to do so. These power calculations were made using a two-sample t-test approach, and an assumed standard deviation for the difference of 11 mmHg.

#### 15.2 Learning Curve Justification

There is no learning curve for the research team. It is expected that there will be a learning curve for the participants self-administering the device. Participants will be provided with information about the device, will be given a dummy demonstration of the device use at the time of the screening as part of their device usage training, and will be supervised on the device usage after the training during the randomization/baseline visit. If participants have any device related queries after the screening/baseline visit they will be advised to contact the Investigation site staff to address those and help them use the device.

#### 15.3 Planned Recruitment Rate

Participants will be recruited over a period of 14 months through the Investigation site. The PIC for this study will be the following:

a. Barts Health NHS Trusts (Primary site/PIC).
b. GP practices and centres that are part of North East London Health and Primary care partnership
c. University College London Hospitals NHS Foundation Trust
d. Homerton University Hospital Foundation Trust.
e. Imperial College Healthcare NHS trust and associated hospitals.
f. St George's University Hospitals NHS Foundation Trusts.
g. King's College London NHS Foundation Trust
h. Royal Free Hospitals NHS Foundation Trust.
i. Broomfield Hospital Mid and South Essex Hospitals Foundation NHS Trust

An estimated recruitment rate of 1 participant per week is planned.

### 16. End of Trial (EOT) Definition

End of trial definition: The date of the last visit of the last participant recruited into the trial; last patient last visit (LPLV).

The CI is delegated the responsibility of submitting the End of Trial (EOT) notification to REC and MHRA once reviewed by the sponsor. The EOT notification must be received by the REC and MHRA within 90 days of the end of the study. If the study is ended prematurely, the CI will notify the Sponsor, REC, and MHRA within 15 days, including the reasons for the premature termination.

#### 16.1 Statistical Analysis

The Statistical Analysis Plan (SAP) will be used in the analysis of trial data and will be finalised prior to any review and/or analysis of data.

#### 16.2 Summary of Baseline Data and Flow of Participants

Baseline data will be presented for the following factors, separately by randomised trial group:

- Age
- Sex
- BMI
- Smoking status
- Alcohol consumption
- Mean 24-hour, daytime, and night-time ABPM SBP and DBP
- Office SBP and DBP
- Vital signs
- Holter ECG
- Blood test measures
- Urine test measures
- Echocardiogram measures
- 6MWT distance

#### 16.3 Analysis of Participant Populations

The main analyses of the primary and secondary endpoints will be conducted on an intention-to-treat population, consisting of all participants with available data based on the trial group to which they were randomized to, irrespective of their compliance to their prescribed treatment as specified in the protocol.

Analyses will also be conducted on a per protocol population consisting of those subjects who complied with treatment as is specified in the protocol, i.e. who have good compliance defined as device usage on 80% or more days during the trial (see Section 11.6 on assessment of compliance).

#### 16.4 Primary Endpoint Analysis

The primary endpoint is the change in daytime ABPM SBP from baseline to the end of the treatment period (3 months). Daytime ABPM SBP will be calculated both at baseline and the end of the study based on the mean daytime SBP between the hours of 7am and 11pm from ABPM recordings.

The change in daytime ABPM SBP will be calculated for each participant by subtracting the end of treatment measurement at 3 months from the baseline measurement.

The crude difference in the mean change in daytime ABPM SBP will be calculated between trial groups, and a two-sample t-test will be used to test the null hypothesis of no difference in the change in SBP between groups.

Adjusted analysis will be conducted using linear regression comparing change in SBP between treatment groups while adjusting for baseline daytime ABPM SBP (ANCOVA), as well as adjusting for age, sex, and BMI. The estimated adjusted difference in change in SBP will be presented, along with 95% confidence intervals (CIs) and p-value.

The analysis will primarily be conducted on the intention-to-treat population and will be repeated secondarily for the per protocol population.

The assumptions of the regression analysis are that the model residuals, i.e. the error terms (difference between predicted and observed values) have a normal distribution with mean zero and equal variance in both treatment groups. It is widely accepted that both SBP and change in SBP are normally distributed, hence that be our underlying assumption. However, assessment of the distribution will be undertaken, and alternative non-parametric approach will be used as an alternative if the assumption of normally is considered to have been violated or if there are very extreme outliers observed. However, an accurate assessment of underlying distributional assumptions is limited in a small sample size such as this.

#### 16.5 Secondary & Exploratory Endpoint Analysis

All secondary endpoints that are continuous will be analysed in the same way as for the primary endpoint, detailed above.

For continuous endpoints, mean, and SD, and number of participants with available data will be presented along with estimated crude and adjusted mean differences between trial arms.

Binary endpoints will be shown as the number with each endpoint and total number in each group along with the percentage. For binary endpoints, the crude and adjusted odds ratios (ORs) will be estimated.

Some secondary and exploratory endpoints will be presented descriptively.

#### 16.5 Feasibility Endpoint Analysis

All feasibility endpoints for this pilot study will be presented descriptively or analysed qualitatively.

#### 16.6 Safety Analysis

Adverse events (AEs) will be summarised using counts and percentages. The number of subjects having at least one AE will be presented overall and tabulated by treatment. The number of subjects with AEs of mild/moderate/severe intensity will be shown overall and by treatment using the maximum severity experienced for each participant. The total number of AEs for each treatment, allowing multiple events per participant, will also be presented.

Serious AEs (SAEs), both non-fatal and fatal will be listed separately along with details of the treatment and whether the event is unexpected and whether it is thought to be related to the treatment.

#### 16.7 Subgroup Analyses

The primary endpoint will be analysed according to the following subgroups:

- BMI at baseline (<30 kg/m$^2$ vs. ≥30 kg/m$^2$)
- Diabetes status at baseline
- Age at baseline (<65 years vs. ≥65 years)
- Mean ABPM daytime SBP at baseline (<160 mmHg vs. ≥160 mmHg).

The primary endpoint will be assessed within each subgroup, and a test for interaction will be conducted between groups for each of the 4 risk factors listed.

#### 16.8 Adjusted Analysis

For the adjusted analysis of the primary endpoint and relevant secondary endpoints, analyses will be adjusted for the four baseline risk factors that were used as minimisation balance factors for randomisation: age, sex, BMI and mean daytime SBP.

#### 16.9 Interim Analysis and Criteria for the Premature Termination of the Study

The are no formal interim analyses planned during this pilot study, and hence no opportunity for the early termination of the study based on trial results from accumulating data.

The DSMC will meet to review accumulating pooled data only. The DSMC may be presented with summary baseline data. They will also be presented with safety event data and can request to be unblinded as to treatment allocation on an event-by-event basis if they find cause for concern.

#### 16.10 Procedure(s) to Account for Missing or Spurious Data

This pilot study will assess data completeness to inform decisions about missing data for a future main trial. Analysis will use all participants with available endpoint data. The level of missing data will be tabulated for each outcome, presenting the number of subjects with a data record and number with missing data, separately by trial arm.

Each endpoint will be analysed on a complete case basis.

For continuous variables, distributional assumptions will be hard to assess with the small sample size for this pilot study. However, if any continuous endpoint variables appear to seriously violate distributional assumptions or very extreme outliers observed, then non-parametric alternative approaches to analysis will be undertaken in such cases.

#### 16.11 Economic Evaluation

.
The Oxford AHSN will undertake an early health economics study to determine the cost implications required for integration of tAN device-based antihypertension therapy within the existing clinical pathways. The Oxford AHSN will also develop an effective strategy to facilitate adoption of the technology by the NHS.

#### 16.12 Other Statistical Considerations

Any changes to the original analysis plan will be recorded in the Statistical Analysis plan document (SAP revision history) along with dates, updated SAP version number and description of and reasons for the changes.

If patients are withdrawn from the trial or are lost to follow-up early, they will not be included in analyses of endpoints where their data are missing.

### 17.0 Data Handling and Record Keeping

#### 17.1 Source Data and Source Documents

ISO14155 GCP section 3.47, defines source data as "all information in original records, certified copies of original records of clinical findings, observations, or other activities in a clinical investigation, necessary for the reconstruction and evaluation of the clinical investigation."

ISO14155 GCP section 3.48, defines source document as "original or certified copy of printed, optical or electronic document containing source data."

To enable review, monitoring, audit and/or inspection of study source data, the CI agrees to keep records of all participating participants. Example of these original documents, and data records include (but are not limited to) electronic participant records/clinical notes, original signed consent form, crib sheets (s), participant trial file, participant diary, participant logbook, recorded data from automated instruments, laboratory notes, questionnaires (paper and/or electronic), evaluation checklists, accountability logs, copies or transcriptions (certified after verification as being accurate and complete) and records kept at the laboratories involved in the trial.

A Source Data Agreement (SDA) will be established by CVCTU Coordinating team detailing a full list of all trial source data documents including what comprises source data, corresponding source data documents and its location. The SDA will be held in the ISF and TMF.

All source data will be collated by the Investigation site team. The Investigation site team will create a separate trial file for each participant which will comprise of information given by the participant from screening visit and throughout the duration on the trial; some trial related documentation deemed as source will be collated in this file.

JRMO Clinical Trial Monitor or delegate will have access to source data and source documents for source data verification aspects of monitoring. Direct access will also be granted to authorised representatives from the Sponsor and the regulatory authorities to permit trial-related monitoring, audits, and inspections.

#### 17.2 Case Report Forms (CRFs)

Pseudonymised trial data will be captured electronically via electronic case report form (eCRF). The CRF will be designed by CVCTU Coordinating team with input from the CI, according to Sponsor requirements and will only collect required trial data, including safety events, that will be used for statistical analysis.

CI, Sub-Investigator(s) and Investigation site team are permitted to record trial data on CRF (s).

The eCRF will be built and managed by Castor EDC personnel and will be held on a secure web application, accessible via HTTPS/SSL.

Direct access to eCRF will be restricted, and only CI delegated and authorised users will be issued with access to the eCRF. Each user will be assigned specific user roles and rights, and this will be reflected on the respective delegation log and maintained in a CASTOR User access log. If a staff member leaves, their access to the eCRF will be revoked.

#### 17.3 Data Capture

Source data will be collected by the Investigator or the research nurses and captured in an electronic case report form (eCRF) using the Castor EDC database (21 CFR Part 11 compliant) with electronic signatures and an audit trail. A source data agreement will be written to specify where the source will be located, and as much data as possible will be collected directly in the eCRF. Direct access to the database will be restricted to relevant and specific users only. The eCRFs will be completed by the Investigator or suitably trained research staff, as designated in the site delegation log, as accurately and completely as possible throughout the study. Patient Identifiable Data (PID) will be encrypted in the database and kept separately from the clinical data.

All source data will be kept securely in the participant study files or ISF within locked cabinets in restricted access rooms. Source data will be reviewed as part of the source data verification during site monitoring.

#### 17.4 Transferring and Transporting Data

All data must be handled in accordance with the Data Protection Act (2018) and no data will be transferred outside of the EEA.

Identifiable information will not be stored, or transported on any portable device (e.g. laptops, memory sticks, CD / DVDs) unless it is encrypted and will not be sent electronically if it is not subject to end-to-end encryption. Barts Health Participant Identifiable Data (PID) will not be taken out of Barts Health without participant consent.

#### 17.5 Data Management

A trial specific Data Management Plan will be developed by CVCTU Coordinating team detailing all key methods of data management for collecting, recording and handling of trial data.

### 18.0 Confidentiality

The CI will be the data custodian for all data generated during the study.

The CI, Investigation trial team and the CVCTU Coordinating team will ensure that all participants' identities are protected at every stage of the trial. To ensure this, at time of consent, each participant will be allocated a unique participant ID number via Castor EDC by Investigation trial team before undergoing any screening procedures. Following participant randomisation and during the study, all participant trial records and samples will be linked by this unique participant ID number.

The CI is responsible for protecting the identity of participants at their site. Participants will be referred to only by their unique trial identifier, whenever data is transferred outside of the site, and in all correspondence between the site and the CVCTU Coordinating team, Sub-Investigator (s), Sponsor, or anyone associated with the study.

No participants will be individually identifiable from any publications resulting from the study.

Information regarding trial participants will be kept confidential and managed in accordance with the Data Protection Act (2018), the Research Governance Framework for Health and Social Care and Research Ethics Committee approval. All trial data will be stored in line with the Medicines for Human Use (Clinical Trials) Regulations 2004 and subsequent amendments and the Data Protection Act. Trial data will be archived in line with the Medicines for Human Use (Clinical Trials) Regulations 2004 and all subsequent amendments, and as defined in the JRMO SOP 20, Archiving.

#### 18.1 De-identification of Participants

Personal identification information (full name, Initial, date of birth and/or NHS number) will only be used on participant consent form (s) and a corresponding screening log which will be maintained by the Investigation site team, throughout the trial. Participant consent form (s) and trial screening log will be held within the ISF, which will be kept within a lockable filing cupboard within a locked room; that only Investigation site team members will have access to.

Only the Investigation site team will have access to participants' full personal identifiable information and electronic participant records/clinical notes, for the purposes of identifying potential participants. Sponsor and CVCTU Clinical Trial Monitor or delegate will be permitted access to participant personal identification information and electronic participant records/clinical notes for monitoring, pharmacovigilance and/or audit purposes.

Participant trial file will be kept within cupboards in a locked room; that only investigation site team members will have access to.

De-identification of participant screening number is via trial screening log maintained by the Investigation site team.

The unblinding code (active or sham device) will be held electronically on Sealed Envelope and by a designated member of Afferent Medical Solution Ltd. Access is limited to maintaining the blind on the trial.

The process to unblind trial participants is detailed in trial specific Randomisation, Unblinding and Maintaining the Blind SOP.

### 19.0 Monitoring, Audit, and Inspection

#### 19.1 Monitoring

The JRMO Clinical Trial Monitor has the responsibility of monitoring the trial.

A trial specific Monitoring Plan will be developed by the JRMO and the CVCTU Coordinating team detailing all monitoring procedures including on-site visits based on the Sponsor risk assessment; the Sponsor and CI will agree on the monitoring plan.

The Investigation site team will be initiated and monitored with in line with JRMO SOP 28 – Monitoring.

#### 19.2 Auditing

The Sponsor retains the right to audit any aspect of the trial, investigation site or central facilities. In addition, regulatory authority and Funder (where applicable) may inspect any part of the trial.

All sites and vendors must inform the Sponsor if notified of any Audit or inspection affecting this trial.

### 20.0 Compliance

The CI will ensure that the trial conduct complies with the principles outlined in the Medical Devices Regulations 2002, current UK Policy Framework for Social and Health Care Research (2017), ISO 14155 GCP guidelines, the World Medical Association Declaration of Helsinki (1996), the Sponsor's SOPs, and other regulatory requirements.

The trial will not commence until Sponsor gives permission to activate the investigation site.

#### 20.1 Non-Compliance

Prospective, planned deviations or waivers to the protocol are not allowed under the UK regulations on Clinical Trials and will not be used (i.e. it is not acceptable to enroll a participant if they do not meet the eligibility criteria or restrictions specified in the study protocol).

A variety of different sources including monitoring visits, corrective and preventative actions (CAPAs), CRFs, communications and updates will capture non-compliances.

Investigation site will record all (site level) protocol deviations on site deviation log. CVCTU will record all (coordinating delegated duty) protocol deviations on CVCTU deviation log. Respective committees (where deemed necessary) will receive a summary of all protocol deviations.

The CI and CVCTU Coordinating team will assess non-compliances and will action a timeframe to deal with them (dependent on the severity), including whether there is a need to escalate to Sponsor, and where required the CI will report to Sponsor as per their guidelines (JRMO SOP31 – Non-Compliance and Serious Breach reporting).

Any event with the potential to affect participant safety or data integrity will be reported to the Sponsor within 24 hours of the CI or the CVCTU Coordinating team becoming aware.

The Sponsor will maintain a log of non-compliances to ascertain if there are any trends developing which need to be escalated.

#### 20.2 Notification of Serious Breaches to GCP and/or the Protocol

A 'serious breach' is a breach, which is likely to affect to a significant degree:

- The safety or physical or mental integrity of the participants of the study; or
- The scientific value of the study.

The CI is responsible for reporting any potential serious breaches to the Sponsor and to the CVCTU Coordinating team within **24 hours** of becoming aware of the event.

The Sponsor is responsible for determining whether a potential serious breach constitutes a serious breach, and will work with the CI to investigate, and notify and report to the MHRA and REC (as applicable) within 7 working days of becoming aware of the serious breach.

#### 20.3 Amendments to the Clinical Investigation Plan

Should the CI or Sponsor deem it necessary to make an amendment to the CIP or to the documents submitted with the Clinical Investigation application, these will be implemented as amendments. The full amendment process is located in JRMO SOP 17, Amendments.

##### 20.4 Suspension or Early Termination of the Clinical Investigation

If the clinical investigation is temporarily suspended, this will be notified to the Ethics committee and competent authority via a substantial amendment. A further substantial amendment will be implemented to resume the clinical investigation.

If a decision is made to terminate the clinical investigation early the ethics committee and competent authority will be notified within 15 days through the submission of an End of Study notification form.

##### 20.5 Contractual Agreements

SCRATCH-HTN Trial is funded by NIHR. Contractual agreements between Sponsor and Afferent Medical Solutions Ltd have been executed, and necessary vendor agreements will be in place before the start of the trial.

#### 21.0 Declaration of Interests

The CI, Sub-investigators, PI (s) at each PIC (s), and committee members for the overall study management, will provide:

- All competing interests (Sponsor requirement).
- Personal or professional relationships with the investigational device manufacturer.
- Ownership interests that may relate to products, services, or interventions considered for use in the study or that may be significantly affecting the study.
- Commercial ties (e.g. pharmaceutical, behavior modification, and/or technology companies).
- Non-commercial potential conflicts (e.g. professional collaborations that may affect academic promotion).

Trial Master Files (TMF) will hold all completed declaration of interests. All enquiries are to be sent to.

#### 22.0 Peer Review

The trial design and methodology has undergone independent, expert and scientific peer reviews by the Scientific Committee of the CVCTU and also extensive review by the Funder. This CIP has also been reviewed by the CIs Institute (QMUL) prior to trial sponsorship in principle given by the Sponsor requirements.

#### 23.0 Public and Patient Involvement (PPI)

Aspect of trial design includes review by Public and Patient Involvement (PPI) and Barts NIHR Biomedical Research Centre Patient and Public Advisory Group (PPAG) in the initial design of all trial participant facing literature, usability of the AffeX-CT device prototype and the design of the trial; to ensure that trial proposal is understandable to intended participant population. PPI representatives will also be invited to be members of the Trial Steering Committee.

### 24.0 Indemnity/Insurance

The Insurance policy that Queen Mary University of London has in place provides cover for the design and management of the study as well as "No Fault Compensation" for participants, which provides an indemnity to participants for negligent and non-negligent harm. A copy of this document will be filled in the TMF and ISF.

### 25.0 Trial Committee (s)

The Trial Committees outlined (below) will be established and run in accordance with the Barts CTU Trial Committees SOP CTU GEN TM 14, Sponsor and Funder requirements.

#### 25.1 Trial Management Group (TMG)

The Trial Management Group (TMG) is mandate for all MHRA regulated studies; members will meet regularly to discuss all aspects of the trial progression.

The terms and conditions of this committee including frequency of meetings is detailed in the TMG charter.

#### 25.2 Trial Steering Committee (TSC)

The Trial Steering Committee (TSC) is mandate for all MHRA regulated studies and the role is to provide the overall supervision of the trial.

A TSC will review and monitor the progress of the trial, ensuring protocol adherence, appropriate action to safeguard participants is taken and the quality of the trial is maintained.

Primarily independent members will form the TSC composition; meetings will occur either face to face or via teleconference and will include an independent chair with at least two other independent members, Participant representative (where deemed necessary), CI/Sub-Investigator (s), Trial Statistician and Trial Manager, Trial Coordinator and/or Afferent Medical Solution Ltd.

The terms and conditions of this Committee including the authority and frequency of meetings is in detail in the TSC charter.

#### 25.3 Data Safety Monitoring Committee (DSMC)

The Data Safety Monitoring Committee (DSMC) will monitor participant safety and treatment and review unblinded trial data whilst the trial is ongoing.

Members of DSMC will be independent of the CI/Sub-Investigator (s), Funders and Sponsors; meetings will occur either face to face or via teleconference and will include 3 to 4 members with at least one Clinician experienced in the clinical area and at least one expert Statistician.

CI/PI/Sub-Investigator(s), Trial Statistician, Trial Manager, Trial Coordinator and/or Afferent Medical Solution Ltd where deemed necessary may receive an invite to the meetings.

The terms and conditions of this Committee including the authority and frequency of meetings is in detail in the DSMC charter.

##### 25.4 Research Steering Group (RSG)

The Research Steering Group (RSG) is a recommendation for the Funder to monitor the performance and technical content of the trial against the protocol. The RSG will critically assess ongoing results of the trial including identify weaknesses/delays, what has been learnt and agree future research. The RSG will operate as the key forum where the Funder, CI and Afferent Medical Solution Ltd shall discuss trial progress, milestones and/or targets.

Members of the RSG will comprise of the CI (RSG chair), at least one independent expert adviser, and one representative of Afferent Medical Solution Ltd and up to two representatives or nominees from the Funder.

The terms and conditions of this Committee is as per ‘Section 7 of the Research Contract between NIHR and QM’.

#### 26.0 Publication and Dissemination Policy

##### 26.1 Publication

Data attained from the entire trial including the sub study or from subsets of the trial will be submitted to peer review journals and published on clinicaltrials.gov website within one year of the End of Trial Definition being met.

The CI will adhere to the requirements laid out by The International Committee of Medical Journal Editors and is responsible for how the manuscript will be written and edited, including the number and order of authors, the journal to which it will be submitted and all other issues.

All publications will be sent to the Sponsor prior to publication and should acknowledge the Sponsor with correct designation “Queen Mary University of London”.

##### 26.2 Dissemination Policy

The data generated from the entire trial including sub-study will be solely owned by Queen Mary University of London and upon completion of the trial, the data will be analysed, tabulated and a Final Trial Report prepared.

The role of Data Controller will be solely Queen Mary University of London. The CI will be Data Custodian for the entirety of the trial, and CVCTU team, Oxford AHSN Afferent Medical Solution Ltd is Data Processor.

On completion of the trial, the CIP, Final Trial Report, anonymised participant level dataset and/or statistical code for generating the results will be accessible on the clinicaltrials.gov website, and release of this information will require permission from the CI and Afferent

Medical Solution Ltd. The participating Investigators and employees of Queen Mary University of London and trial Partners will have rights to publish any of the trial data.

There are no time limits or review requirements on the publications, but the funder of the trial is to be acknowledged.

Participants will not be notified of the outcome of the trial, but may request access to their own trial data from the CI after the Final Trial Report has been compiled and results have been published.

If professional medical writers are required, their engagement will be acknowledged.

#### 26.3 Access to The Final Trial Dataset

The CI, Investigation site team, delegated CVCTU Coordinating Team, Health Innovation Oxford and Thames Valley (Health Economics analysis) and Afferent Medical Solution Ltd will have access to the Final Trial Dataset via the eCRF data extracts for analysis and reporting.

A full,detailed list of whom will have access to the Final Trial Dataset will be as per the Data Management Plan and will outline the access, data entry, upload and/or transfer.

The RSG, TSC and DSMC will have interim access as per Committee Charter requirements.

The datasets generated during the study and to be analysed at study closure for SCRATCH-HTN will be available upon reasonable request after embargo period of 12 months from the date of publication of main findings from the CI. The anonymised, processed data will be available after one year from the publication of main findings for a further period of 5 years after, and all study participants have consented with this data sharing. Other requests for access to SCRATCH-HTN data will need to specify the reason for the request and how the data will be processed, and data will be shared using secure data transfer methods.

### 27.0 Archiving

During the trial the CI is responsible for oversight of all trial documents ensuring secure conditions, safe locations and retention of all records.

Archiving of all trial documentation including the eCRF at the End of Trial will be authorised by the Sponsor, following the submission of the End of Trial Report.

All essential documents maintained in TMF, ISF, participant files along with trial dataset downloaded and printed from the eCRF will be archived for 25 years after the completion of the trial as per Sponsors requirements and Barts Health Policy.

Any destruction of essential documents will require authorisation from the Sponsor.

### 28.0 References

1. Markle W, M. Fisher, and R. Smego. Understanding Global Health. Economics and Global Health. 2007.
2. Williams B, Mancia G, Spiering W, Agabiti Rosei E, Azizi M, Burnier M, Clement DL, Coca A, de Simone G, Dominiczak A, Kahan T, Mahfoud F, Redon J, Ruilope L, Zanchetti A, Kerins M, Kjeldsen SE, Kreutz R, Laurent S, Lip GYH, McManus R, Narkiewicz K, Ruschitzka F, Schmieder RE, Shlyakhto E, Tsioufis C, Aboyans V, Desormais I and Group ESCSD. 2018 ESC/ESH Guidelines for the management of arterial hypertension. *Eur Heart J*. 2018;39:3021-3104.
3. England PH. Health matters: preventing cardiovascular disease 2019.
4. Foundation BH. BHF Coronavirus and Heart & Circulatory Disease Statistics. 2020.
5. Egan BM, Kjeldsen SE, Grassi G, Esler M and Mancia G. The global burden of hypertension exceeds 1.4 billion people: should a systolic blood pressure target below 130 become the universal standard? *J Hypertens*. 2019;37:1148-1153.
6. England PH. Hypertension prevalence estimates in England. 2017;2020.
7. England PH. Tackling high blood pressure From evidence into action. 2014.
8. Achelrod D, Wenzel U and Frey S. Systematic review and meta-analysis of the prevalence of resistant hypertension in treated hypertensive populations. *Am J Hypertens*. 2015;28:355-61.
9. Abegaz TM, Shehab A, Gebreyohannes EA, Bhagavathula AS and Elnour AA. Nonadherence to antihypertensive drugs: A systematic review and meta-analysis. *Medicine (Baltimore)*. 2017;96:e5641.
10. Carey RM, Calhoun DA, Bakris GL, Brook RD, Daugherty SL, Dennison-Himmelfarb CR, Egan BM, Flack JM, Gidding SS, Judd E, Lackland DT, Laffer CL, Newton-Cheh C, Smith SM, Taler SJ, Textor SC, Turan TN, White WB, American Heart Association Professional/Public E, Publications Committee of the Council on H, Council on C, Stroke N, Council on Clinical C, Council on G, Precision M, Council on Peripheral Vascular D, Council on Quality of C, Outcomes R and Stroke C. Resistant Hypertension: Detection, Evaluation, and Management: A Scientific Statement From the American Heart Association. *Hypertension*. 2018;72:e53-e90.
11. Dahal K, Khan M, Siddiqui N, Mina G, Katikaneni P, Modi K, Azrin M and Lee J. Renal Denervation in the Management of Hypertension: A Meta-Analysis of Sham-Controlled Trials. *Cardiovasc Revasc Med*. 2020;21:532-537.
12. Gupta A, Prince M, Bob-Manuel T and Jenkins JS. Renal denervation: Alternative treatment options for hypertension? *Prog Cardiovasc Dis*. 2020;63:51-57.
13. Azizi M, Schmieder RE, Mahfoud F, Weber MA, Daemen J, Lobo MD, Sharp ASP, Bloch MJ, Basile J, Wang Y, Saxena M, Lurz P, Rader F, Sayer J, Fisher NDL, Fouassier D, Barman NC, Reeve-Stoffer H, McClure C, Kirtane AJ and Investigators R-H. Six-Month Results of Treatment-Blinded Medication Titration for Hypertension Control Following Randomization to Endovascular Ultrasound Renal Denervation or a Sham Procedure in the RADIANCE-HTN SOLO Trial. *Circulation*. 2019.
14. Kandzari DE, Bohm M, Mahfoud F, Townsend RR, Weber MA, Pocock S, Tsioufis K, Tousoulis D, Choi JW, East C, Brar S, Cohen SA, Fahy M, Pilcher G, Kario K and Investigators SH-OMT. Effect of renal denervation on blood pressure in the presence of antihypertensive drugs: 6-month efficacy and safety results from the SPYRAL HTN-ON MED proof-of-concept randomised trial. *Lancet*. 2018;391:2346-2355.
15. Clancy JA, Mary DA, Witte KK, Greenwood JP, Deuchars SA and Deuchars J. Non-invasive vagus nerve stimulation in healthy humans reduces sympathetic nerve activity. *Brain Stimul*. 2014;7:871-7.
16. Bretherton B, Atkinson L, Murray A, Clancy J, Deuchars S and Deuchars J. Effects of transcutaneous vagus nerve stimulation in individuals aged 55 years or above: potential benefits of daily stimulation. *Aging (Albany NY)*. 2019;11:4836-4857.

17. Redgrave J, Day D, Leung H, Laud PJ, Ali A, Lindert R and Majid A. Safety and tolerability of Transcutaneous Vagus Nerve stimulation in humans; a systematic review. *Brain Stimul*. 2018;11:1225-1238.
18. Ettehad D, Emdin CA, Kiran A, Anderson SG, Callender T, Emberson J, Chalmers J, Rodgers A and Rahimi K. Blood pressure lowering for prevention of cardiovascular disease and death: a systematic review and meta-analysis. *Lancet*. 2016;387:957-967.
19. van Bilsen M, Patel HC, Bauersachs J, Bohm M, Borggrefe M, Brutsaert D, Coats AJS, de Boer RA, de Keulenaer GW, Filippatos GS, Floras J, Grassi G, Jankowska EA, Kornet L, Lunde IG, Maack C, Mahfoud F, Pollesello P, Ponikowski P, Ruschitzka F, Sabbah HN, Schultz HD, Seferovic P, Slart R, Taggart P, Tocchetti CG, Van Laake LW, Zannad F, Heymans S and Lyon AR. The autonomic nervous system as a therapeutic target in heart failure: a scientific position statement from the Translational Research Committee of the Heart Failure Association of the European Society of Cardiology. *Eur J Heart Fail*. 2017;19:1361-1378.
20. Rahimi K. Blood pressure reduction – lower really is better. *European Society of Cardiology*. 2020.
21. Veldsman M, Tai, XY., Nichols, T. et al. Cerebrovascular risk factors impact frontoparietal network integrity and executive function in healthy ageing. *Nat Commun 11, 4340 (2020)*. 2020.
22. Mueller ST and Piper BJ. The Psychology Experiment Building Language (PEBL) and PEBL Test Battery. *J Neurosci Methods*. 2014;222:250-9.
23. Voils CI, Maciejewski ML, Hoyle RH, Reeve BB, Gallagher P, Bryson CL and Yancy WS, Jr. Initial validation of a self-report measure of the extent of and reasons for medication nonadherence. *Med Care*. 2012;50:1013-9.
24. Bang H, Ni L and Davis CE. Assessment of blinding in clinical trials. *Control Clin Trials*. 2004;25:143-56.
25. Bastien CH, Vallieres A and Morin CM. Validation of the Insomnia Severity Index as an outcome measure for insomnia research. *Sleep Med*. 2001;2:297-307.

### Appendix 1: Autonomic Function Test procedure

(1) Reassure the participant so he/she can feel comfortable.

a) Explain the test from the safety point of view and give reassurances and mention the painless procedure.
b) Give a brief summary to the participant and to what is involved.

(2) Prepare the participant for the procedure.

a) Participant should be rested comfortably in a reclining position with the neck being well supported.
    i. Explain to the participant what you are going to do – explain briefly and give sufficient reassurances.
b) Prepare the electrodes.
    i) ECG electrodes sites must be cleaned and scarified.
    ii) Blood gas (transcutaneous blood gas): skin must be cleaned without scarification but strongly preferably until red to encourage the increase in blood flow. This sight is at subcostal region on the mid-clavicle line of the body next to the liver where it is warm.
c) Attach the pre-gel ECG electrodes onto the participant.
d) Apply the breathing belt. This should be placed at the xiphisternal level. The slackness must first be removed by pulling and tightening. The hook should be tightened such that the eye of the belt is at xphisternal level. Next the belt should be pulled and dragged so that the eye from the xiphisternum to the mid-clavicular is lined up to create tension on the belt. This is usually sufficient tension for all body types.
e) Connect the ECG leads in such a way that the red goes to the right shoulder and black goes to the left shoulder and the yellow goes to the sixth intercostal space in the mid-clavicular line or below the breast.
f) Apply the contact liquid in the form of drops into the fixation cups and then screw the transcutaneous gas probes tightly into the fixation cups, then secure them with either millipores adhesive tape or similar.
g) Enquire from the participant whether they are left or right (L/R) handed so that the dominant side can be determined. Apply the photo-plethysmograph finger cup to the participant's middle finger of the non-dominant hand (because the dominant hand will be used later for isometric exercise). Then connect the set up to the 'FINA Press' control box and secure the control box at the back of the hand using the available velcro.

h) Apply a sling around the participant neck and hand bearing the control box on the sling to alleviate the weight of the box and make sure that the finger with the photo-plethysmograph is at the level of heart.

i) Start the 'FINA Press' and make sure that the blood pressure waveform is normal and regular.

j) Switch the 'NeuroScope' ON and enter the participant details in the Vagos software.

(3) Data login and check in – Start data login.

a) Make sure that the breathing trace is central on screen and adjust the gain to show reasonable visual breathing amplitude (if not then it may be due to incorrect tension of the belt). It may need to be adjusted accordingly.
b) Check and make sure that the ECG signal is of good amplitude and is noise free. You may have to swap electrode positions to obtain optimum R-waves.
c) Check and make sure that the scale of CVT is optimally displayed (if it is too high CVT then needs adjustment so that it can be analyzed).
d) Check and make sure that the BP waveform is continuous and without jitter. You may have to change the photo-plethysmograph finger cuff to the correct size more appropriate to the size of the participant's finger.
e) Check and make sure that the $O_2$ and $CO_2$ traces have been properly recorded.
f) Check and make sure that the contractility and pre-ejection period have also been recorded properly.
g) Place a marker for the start for the recline position, then turn off the re-calibration server in the Finometer.

### Appendix 2: Be Part of Research Volunteer Service

The purpose of the Be Part of Research Volunteer Service (BPORVS) is to allow members of the public to become volunteers by creating an account, specifying the areas of research that they are interested in and give consent to be contacted by the Be Part of Research team. Those who consent will receive information about BPORVS, in particular to alert them to specific BPORVS registered studies that they may be interested in, based on their volunteered details and study specific eligibility criteria, using an online self-registration service. The register is open to those that live in the UK, are over 18 and have an email address.

At the time of registration, volunteers are made aware that they are not signing up to take part in a specific health study when they join this register and that they will only be signposted to studies that have NIHR funding or are listed on the NIHR CRN Portfolio. If the volunteer is interested in the study there will be a link in the email to take them to the study team (e.g. website, pre-screener) where they will move into the study teams screening process and consenting process if they take part in the study.

The Be Part of Research Volunteer Service is funded by the Department of Health and Social Care and delivered by the National Institute for Health and Care Research (NIHR) in conjunction with Public Health Agency, Research & Development, Northern Ireland, NHS Scotland and Health and Care Research Wales.

Further information on the Be Part of Research Volunteer Service is available here: https://bepartofresearch.nihr.ac.uk/volunteer-service/researchers

**This protocol is based on JRMO Protocol template for MHRA Regulated Studies; Version 6.0, February 2021.**

### 4b. SCRATCH HTN CIP V10.0 (final) - 29OCT24

Final Audit Report 2024-10-29

| | |
|---|---|
| Created: | 2024-10-29 (Greenwich Mean Time) |
| By: | Jane Field |
| Status: | Signed |
| Transaction ID: | CBJCHBCAABAAqRm3s3FDqGZ3-C6LdWQZsW6P_-DFu1EM |

#### "4b. SCRATCH HTN CIP V10.0 (final) - 29OCT24" History

Document created by Jane Field
2024-10-29 - 10:23:13 AM GMT

2024-10-29 - 10:24:19 AM GMT

2024-10-29 - 1:30:52 PM GMT

Document e-signed by Ajay Gupta
Signature Date: 2024-10-29 - 1:32:34 PM GMT - Time Source: server

2024-10-29 - 1:32:37 PM GMT

2024-10-29 - 1:55:57 PM GMT

Document e-signed by Annastazia Learoyd
Signature Date: 2024-10-29 - 1:56:36 PM GMT - Time Source: server

Agreement completed.
2024-10-29 - 1:56:36 PM GMT

Adobe Acrobat Sign
